# Imputation of fluid intelligence scores reduces ascertainment bias and increases power for analyses of common and rare variants

**DOI:** 10.1101/2025.06.18.25329418

**Authors:** David M. van den Berg, Wei Huang, Daniel S. Malawsky, Petr Danecek, Klaudia Walter, Ewan Birney, Karin J. H. Verweij, Dirk J. A. Smit, Sarah J. Lindsay, Matthew E. Hurles, Abdel Abdellaoui, Hilary C. Martin

## Abstract

Studying the genetics of measures of intelligence can help us understand the neurobiology of cognitive function and the aetiology of rare neurodevelopmental conditions. The largest previous genetic studies of measures of intelligence have used ∼270k individuals who completed the fluid intelligence (FI) test in UK Biobank. Here, we integrate additional FI measures in this cohort and leverage eighty-two correlated variables to impute FI values for unmeasured individuals, increasing the sample size to >450k. Through population-based and within-family genome-wide association studies and downstream analyses, we show that this imputation produces a phenotype that genetically resembles measured FI and reduces ascertainment bias within the cohort. We further show that combining measured and imputed FI scores increases the number of independent SNP associations (p<5×10^-8^) from 385 to 608 and increases polygenic score accuracy in external cohorts by 15% on average. Additionally, incorporating imputed FI scores increases the number of gene-level associations with rare variants from five to twenty-six (FDR<1%). These include fourteen well-established developmental disorder-associated genes, a four-fold enrichment (p=8×10^-8^); for several of these, our results suggest that loss-of-function variants in the gene impact neurodevelopment, in addition to the previously documented altered-function variants. We also implicate twelve genes without strong prior evidence of association developmental disorders, of which eight have not been previously linked to intelligence (*ROBO2, RB1CC1, ANK3, CHD9, TLK1, PCLO, DPP8, IPO9)*. These twelve genes were significantly enriched for *de novo* loss-of-function mutations in a set of >31k patients with developmental disorders (p=6.8×10^-4^). We further identify three genes showing significant rare variant associations with educational attainment but not with FI, including *CADPS2* in which, unusually, protein-truncating variants show a positive association. Our results demonstrate the power of phenotype imputation for genetic studies and suggest that incorporating genetic association results for cognitive phenotypes in the general population could help discover new developmental disorder genes.

## Introduction

Various measures of intelligence are significantly associated with many life outcomes, including educational attainment^1^, occupational success^2^, income^3^, and mental^4,5^ and physical health^6,7^. Understanding the genetic architecture of intelligence can help uncover the biological basis of cognitive functioning and the aetiology of rare neurodevelopmental conditions, which are negatively genetically correlated with measures of intelligence^8^. It would also facilitate better-powered studies into the role of intelligence in socio-economic and health-related disparities.

Pedigree-based studies have unequivocally shown that genetics makes a substantial contribution to variation in measures of intelligence^9–11^, with the heritability of “general intelligence” estimated at ∼0.6-0.8 from twin and sibling-regression studies^12,13^. The largest GWAS of intelligence measures to date^14^ (N=269,867) was a meta-analysis of 14 cohorts including UK Biobank (UKB; ∼73% of the total sample) and COGENT consortium^15^ (∼13%) conducted by Savage et al.. They reported 205 associated genomic loci and a SNP heritability of 0.19^16^. A recent within-family GWAS meta-analysis conducted by Tan et al.^17^ estimated direct genetic effects on measures of intelligence (i.e., the influence of an individual’s genotype on their own phenotype, removing the effects of genetic nurture mediated by relatives), and quantified the confounding due to residual population stratification and assortative mating. The within-family SNP heritability was estimated at 0.188, highly similar to the population GWAS-based estimate. Moreover, the genetic correlation between direct effects and the population effects was not significantly different from 1 when estimated with linkage disequilibrium score (LDSC) regression^18^ which removes uncorrected population stratification. However, over half of the variance in population-based GWAS effects was uncorrelated with the direct genetic effects and likely reflects residual population stratification. These findings suggest that common genetic variants contribute substantially to variation in measures of intelligence both within and between families, but that the population-based GWAS results may be biased by residual confounding.

Recent studies have also investigated the contribution of rare variants to measures of intelligence in UKB^19,20^. These studies have shown that rare protein-truncating variants (PTVs) in genes intolerant of loss-of-function^21^ are associated with lower scores on intelligence tests^19,20^; such variants are also implicated in rare developmental disorders (DDs)^22,23^. Chen et al. identified seven genes in which rare PTVs are negatively associated with measures of intelligence^20^, of which three are established DD-associated genes^20^. The rare variants in these genes have substantially larger effect sizes on the measures of intelligence used (∼0.25-1 standard deviations, SDs) than common variants implicated through GWAS (typically <0.1 SD)^24^. Increasing sample sizes in rare variant analyses should enable the discovery of additional genes associated with measures of intelligence.

The intelligence measure in UKB that these studies used is a measure of verbal-numerical reasoning commonly referred to as “fluid intelligence” (FI), the ability to solve novel problems independent of prior knowledge^25^. It was measured using a thirteen- or fourteen-item survey in the baseline in-person visit and in subsequent online questionnaires completed at different times (Supplementary Note 1, Supplementary Table 1, Supplementary Figure 1). This FI test has a test-retest correlation of ∼0.6 (Supplementary Table 2) and correlates ∼0.55 with a measure of “general cognitive ability” estimated from a set of gold-standard cognitive tests^26^. To date, ∼62% of UKB participants have completed the FI test at least once, but previous studies have analysed only a subset of these. Combining the different FI measures is complicated by the fact that the distribution of FI scores differs significantly between surveys (Supplementary Figure 2A). This could be due to several factors, including differences between the online and in-person tests, the fact that participants were experiencing cognitive decline over time, and selection biases in the characteristics of participants taking the different tests. UKB is also known to be biased towards more educated people, who tend to score higher on intelligence tests^27,28^. Participants who completed FI tests tended to have higher educational attainment and experience lower socio-economic deprivation within UKB, particularly for the non-baseline surveys (Supplementary Figure 2B). These selection biases have been shown to distort genetic associations, affecting downstream estimates of heritability and genetic correlations, and Mendelian randomization analyses^29^.

Inspired by recent work in major depressive disorder^30^, we sought to integrate FI values across the various FI tests in UKB and impute missing values for the 38% of individuals of European ancestries who did not complete any of them. We predicted this would ameliorate ascertainment bias and boost power for downstream genetic analyses. We extensively tested multiple approaches to integrate all FI tests and impute missing FI scores. We evaluated these approaches by examining imputation accuracy, SNP heritability, genetic correlations with other traits, and summary statistics obtained from within-family GWAS. Then we used the integrated and imputed FI scores to obtain the largest common-variant GWAS for intelligence to date (N=490,934) and additionally boosted power for rare variant-based analyses (N=438,285). For a less technical description of the paper and ethical considerations, see the General Lay Summary and Frequently Asked Questions (FAQ) section in the Supplementary Information.

## Results

**Box 1:** Glossary of Abbreviations and Terms.

| Box 1: Glossary of Abbreviations and Terms |  |
| --- | --- |
| Abbreviation | Description |
| **Savage et al. Intelligence** | GWAS meta-analysis of measures of intelligence in 14 cohorts, including UKB, from Savage et al.<sup>14</sup> |
| **COGENT** | GWAS meta-analysis of measures of intelligence across 24 cohorts in the COGENT consortium from Trampush et al.<sup>15</sup>. No UKB participants included. |
| **NonCogEA** | GWAS of the “non-cognitive component” of educational attainment, derived using GWAS-by-subtraction by Demange et al.<sup>31</sup> |
| **FIS** | Fluid Intelligence Score in UKB |
| **FIS1** | FIS measured at baseline (20016-0.0) |
| **Earliest FIS** | Earliest measured FIS across five measured tests (Supplementary Table 1) |
| **Average FIS** | Average measured FIS across five measured tests (Supplementary Table 1) (named ImpFIS-average in the supplement) |
| **Observed FIS** | Term used in the main text to describe “Average FIS” in downstream analyses |
| **Imputed FIS** | Term used in the main text to describe imputed FIS produced by inputting Average FIS into the imputation along with 82 other variables |
| **Combined FIS** | Mega-analysis of Observed and Imputed FIS (named MegaFIS-average in Supplement) |

### Integration of measured fluid intelligence phenotypes

We first set out to integrate FI scores (FIS) measured at five different time points (Supplementary Note 1; Supplementary Table 1; Supplementary Figure 1). After adjusting for age and sex (see Methods), we evaluated two approaches for integration of the transformed FIS values (Extended Data Figure 1): (i) taking the average of available scores (average FIS), and (ii) taking the first available score (earliest FIS). We compared these approaches to using only the FI test taken at the initial assessment visit (FIS1). To assess the characteristics of these three phenotypes, we conducted population-based GWAS (see Methods) and then considered the SNP heritability (h^2^ ) and the genetic correlations (r_g_) with a range of traits (Supplementary Figure 3; Supplementary Table 3). In particular, we examined genetic correlations with two previous GWAS of measures of intelligence^14,15^, and, as negative controls, GWAS for height^32^ and the non-cognitive component of educational attainment (NonCogEA)^31^. The NonCogEA GWAS was derived by ‘subtracting’ the association signal from a cognition GWAS (a meta-analysis of the COGENT GWAS with a GWAS of FIS in UKB) from the GWAS of educational attainment (EA) by Lee et al.^33^. NonCogEA therefore reflects factors other than cognitive ability that contribute to EA, which we want to minimize in a GWAS for either observed or imputed fluid intelligence; however, we note that NonCogEA may also capture aspects of cognitive ability which are not measured by the UKB FI test or the COGENT cognition tests.

Integrating the phenotypes using the average and earliest FIS approaches increased the sample size from 146,625 (FIS1) to 283,587 individuals (Figure 1A). Accordingly, the number of FIS-associated lead SNPs identified using FUMA^34^ increased from 92 for FIS1 to 385 and 333 for the average and earliest FIS approaches, respectively (Figure 1D). The estimated h^2^ for FIS1 was 0.23 (SE = 0.01), which was marginally higher than that estimated for average FIS (0.21, SE = .007) and earliest FIS (0.20, SE = .006) (Figure 1B). There were no significant differences between the integrated FI scores and FIS1 in terms of their patterns of genetic correlation with measured intelligence, height or NonCogEA (Figure 1C), suggesting GWAS of these different summary measures captured similar signals to FIS1.

**Figure 1.**
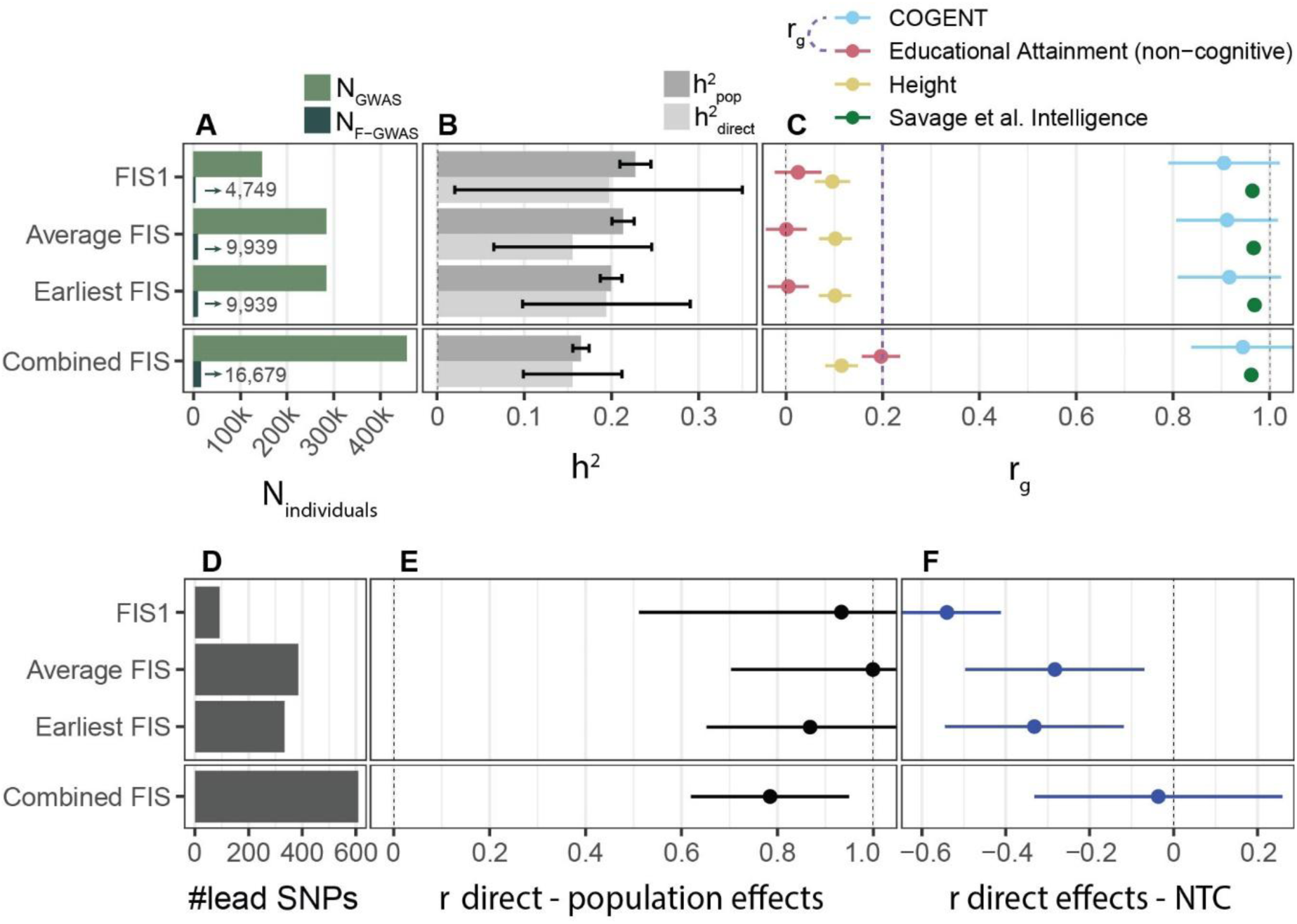
Evaluation of different data integration and imputation approaches. All panels give various summary statistics from either population-based or within-family GWAS of measured FI phenotypes (top three results) and via mega-analysis of average measured and imputed FIS (“Combined FIS”; bottom result). **A)** Sample sizes of population GWAS (N_GWAS_) and within-family GWAS (N_F-GWAS_). **B)** Population and direct effect heritabilities estimated using LDSC^18^. **C)** Population effect genetic correlations estimated using LDSC^35^ with traits from external studies of measures of intelligence (blue and green), the non-cognitive component of educational attainment (NonCogEA) (red), and height (yellow). The purple dashed line indicates the genetic correlation between the COGENT GWAS and NonCogEA. **D)** Number of significant (p<5×10^-8^) independent lead SNPs from the population-based GWAS computed using FUMA^34^. **E)** Genome-wide correlations estimated using LDSC between the population effects and the direct effects from a within-family GWAS conducted on the indicated trait. **F)** Correlations between direct effects and non-transmitted coefficients (NTC) estimated using snipar^36^ for the indicated trait. Error bars indicate 95% confidence intervals.

We first considered the heritability attributed to direct genetic effects (Figure 1B). For FIS1, this was not significantly different from the population GWAS heritability estimate (h^2^_direct_ = 0.20, SE = 0.090; h^2^_pop_ = 0.23, SE = 0.009), and is concordant with prior estimates (h^2^_direct_ = 0.19, SE = 0.027)^17^. Earliest FIS had a similarly high h^2^_direct_, whereas average FIS had a lower, but not significantly different, point estimate. Furthermore, we found that the genetic correlation between the direct effects and the corresponding population effects were not significantly different from 1 in any of these GWAS for FI, suggesting the genetic signals captured in the population GWAS largely resemble those of the direct genetic effects after accounting for residual population stratification accounted for by LDSC regression^18^ (Figure 1E).

Next, we considered the correlation between the direct effects and NTCs (r_direct-NTC_). A negative correlation has previously been observed for a range of phenotypes including FI in UKB, potentially due to ascertainment bias^36^. Indeed, we replicated the previously observed negative r_direct-NTC_ for FIS1 (Figure 1F). We hypothesized that including a larger fraction of UKB by integrating the various FI tests would ameliorate the ascertainment bias and hence attenuate this negative correlation. In line with this, when using average or earliest FIS, the r_direct-NTC_ estimates were closer to 0 (r_g_=-0.28, r_g_=-0.33 respectively) than they were for FIS1 (r_g_=-0.54), though still significantly negative (Figure 1F). Collectively, these results suggest that, despite the slight differences between the various FIS measurements (Supplementary Note 1), phenotype integration yielded a trait that is highly genetically similar to FIS1 while reducing ascertainment biases.

### Imputation of fluid intelligence scores

Although integration of FIS measurements substantially increased the sample size compared to using FIS1 alone, there were over 170,000 UKB individuals with no FI test taken at any time point. We set out to impute missing FI scores by leveraging correlated variables, in order to expand the sample size for genetic analyses and reduce artefacts in downstream analysis that might arise from residual biased ascertainment. We evaluated several imputation approaches (Supplementary Figure 5), including different sets of variables in the imputation and different methods for combining measured and imputed FI scores in a single GWAS. After extensive evaluation (Supplementary Figures 4 and 6; Supplementary Notes 2-3), we selected an approach based on imputation of average FIS to take forward for downstream analyses. In this approach, an average FIS measure was imputed for individuals without any FIS measure using 82 correlated variables. Imputed measures were then combined with the observed average FIS (henceforth “observed FIS”) via a mega-analysis that we will henceforth call “combined FIS”. The accuracy of imputation was assessed in a separate analysis based on the correlation between observed and imputed FIS for a set of randomly chosen participants (Supplementary Table 5), whose measured values were removed prior to imputation. Our chosen strategy achieved an accuracy of r=0.46 (SE = .004), which is slightly lower than the test-retest correlation of FI we found in UKB (Supplementary Table 2, r ≈ 0.6). The accuracy was highest amongst individuals whose observed FIS measure was in the lowest tercile (r = 0.33), versus those in the middle or highest tercile (r = 0.16 and r = 0.20 respectively) (Supplementary Figure 9).

Combining observed and imputed average FIS via mega-analysis (“combined FIS”) increased the sample size to 455,636 individuals while maintaining a heritability estimate (h^2^ = 0.16, SE = 0.005) comparable to that of measured FIS, and with similarly minimal attenuation of h^2^ relative to h^2^ (Figure 1B). Notably, it also resulted in 608 lead SNPs, which is substantially more than analyses using only observed FI measures (e.g. 385 for average observed FIS; Figure 1D; Extended Data Figure 2). The genetic correlation profile of combined FIS was highly similar to that of the COGENT GWAS (Extended Data Figure 3; Supplementary Figure 3); importantly, genetic correlations with height and NonCogEA were consistent with those found with the COGENT GWAS (Figure 1C). Additionally, the point estimate for r_direct-NTC_ was closer to zero than was observed using measured FIS alone, and was not significantly different from zero (Figure 1F), suggesting that imputation further ameliorated the ascertainment bias observed when using only the measured FI values.

### Improved power for common variant analyses of intelligence

To boost power even further, we performed a meta-analysis of combined FIS with the COGENT GWAS (combined FIS+COGENT), increasing the sample size to 490,934. The resulting meta-analysis revealed 662 lead SNPs while the SNP heritability remained 0.16 (SE = 0.004). We compared results from the GWAS of FIS1 (N=146,625), observed FIS (N=283,587), combined FIS (N=455,636) and combined FIS+COGENT (N=490,934) by examining gene prioritization with MAGMA, tissue enrichment and polygenic index (PGI) performance in external cohorts.

The number of prioritized genes (FDR<1%; Supplementary Table 8) increased substantially and monotonically with the size of the GWAS (Figure 2A), indicating that the integration, imputation, and cross-cohort meta-analytic approaches each increased power. In particular, the number of prioritized genes increased by a factor of 3.6 between the FIS1 (smallest) and cross-cohort (largest) analyses. The vast majority of genes were prioritized by at least two different GWAS. MAGMA gene-based p-values were then used to assess enrichment for expression in 54 GTEx tissues (Figure 2B; Supplementary Table 10). Across each GWAS, prioritized genes were only significantly (p<0.05/54) enriched in brain tissues, and the degree of enrichment became more significant as the GWAS sample size increased, suggesting that the integration of missing FI measures was producing a biologically-relevant phenotype.

**Figure 2.**
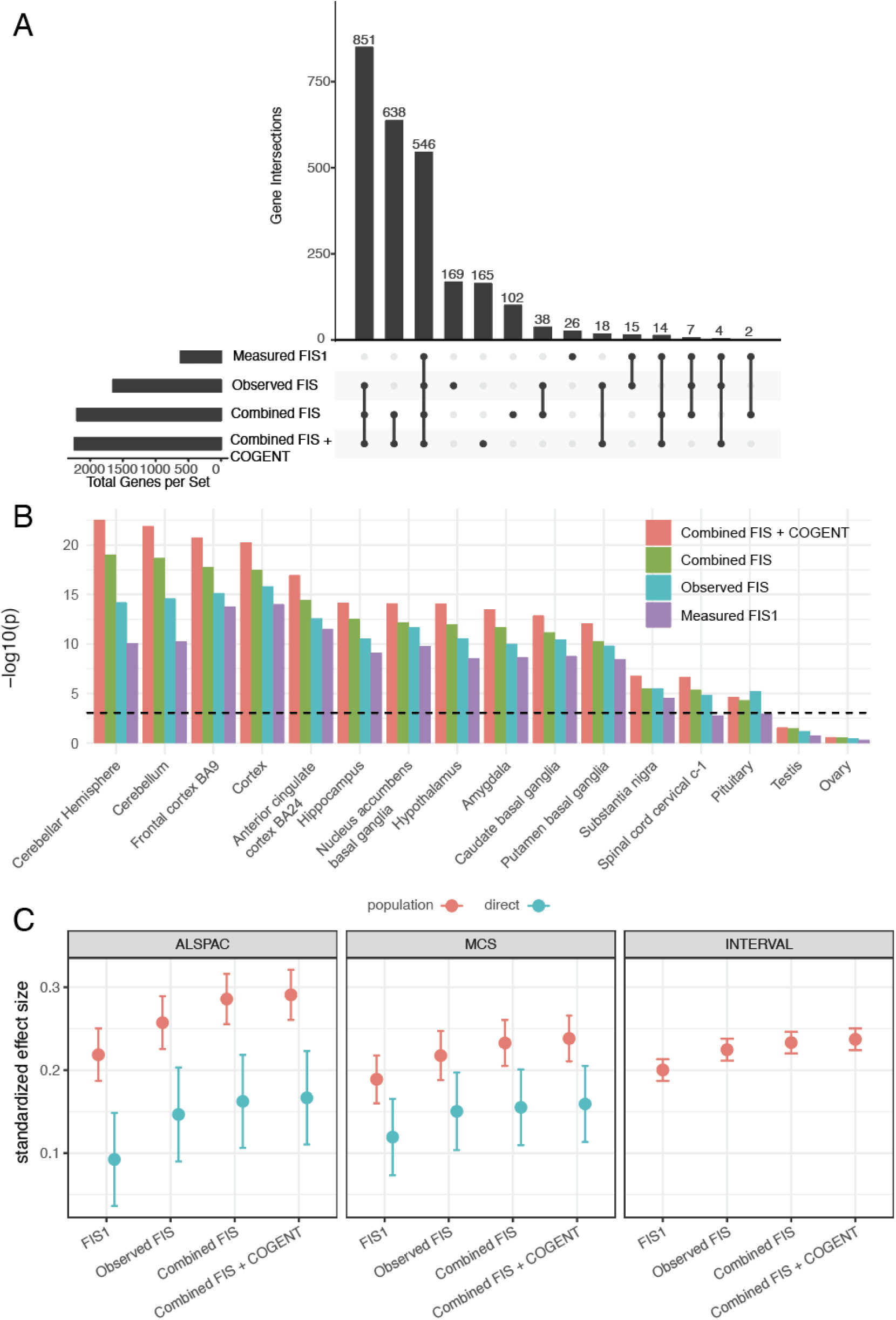
Integration and imputation increases power for downstream analyses of GWAS results. **A)** An upset plot showing the overlap of protein-coding genes prioritized by each GWAS using MAGMA (FDR<1%). **B)** - log10(p) values for enrichment analyses of MAGMA-prioritized genes amongst 54 GTEx v8 tissues using FUMA. The dashed line indicates a Bonferroni-corrected -10log(p-value) threshold of 0.05/(54 tissues tested). The figure shows the tissues that passed Bonferroni correction plus two others (testis and ovary) for comparison (Full results in Supplementary Table 10). **C)** Associations between PGIs constructed with summary statistics from the various GWAS and cognitive measures in three independent cohorts. Points show standardized effect sizes with lines indicating 95% CIs. In ALSPAC we used full-scale IQ measured at age 8. In MCS we used a cognitive performance measure derived from a factor analysis using a combination of various cognitive tests administered across childhood (age 3-7). In INTERVAL we used the average of the two fluid intelligence tests administered 24 and 48 months after recruitment. The population effects in red indicate results from a regression of the PGIs on the cognitive ability measure, controlling for sex and 20 PCs. The direct effects were estimated in the two birth cohorts by additionally controlling for parental PGIs in the regressions.

We derived PGIs based on the various GWAS and tested them in three independent UK-based cohorts with genetically-inferred European ancestries (Supplementary Methods). Specifically, we examined IQ at age 8 in the Avon Longitudinal Study of Parents and Children (ALSPAC; N=5,283)^37,38^, a combined measure of cognitive performance in early childhood (ages 3-7) in the Millennium Cohort Study^39^ (MCS; N=5,621), and the same FI measure as UKB in the INTERVAL study of blood donors (N=20,328)^40^. We observed significant associations between all PGIs and the intelligence measure in all cohorts, with the greatest variance explained in ALSPAC, likely because the measure of intelligence used there (the Wechsler Intelligence Scale Test) has lower measurement error than the measures used in MCS and INTERVAL. In all three cohorts, the variance explained by the PGIs increased monotonically with sample size, regardless of whether the increase was due to integration, imputation, or meta-analysis with external cohorts. (Figure 2C, Supplementary Table 9). The same trend was observed for direct effect estimates obtained by controlling for parental PGIs in ALSPAC and MCS, suggesting the increased variance explained was at least partly due to improved within-family prediction. This association between discovery GWAS sample size and predictive power of the PGI was significant in a multilevel mixed effects model when considering both the population and direct effect estimates (beta = 0.036, p = 2.3×10^-10^) or the direct effect estimates alone (beta = 0.043, p = 0.022). On average across cohorts, the population effect variance explained by the combined FIS *versus* the observed FIS PGI increased by 15.0%, and the variance explained within families increased by 14.7%. These results suggest that the combined FIS GWAS resulted in increased power for genetic prediction of measured cognitive ability in external cohorts, providing additional validation for the use of imputed FI in genetic studies.

### Improved power for rare variant analyses of fluid intelligence

Fluid intelligence is known to be negatively associated with the burden of rare protein-truncating variants (PTVs) and damaging missense variants, particularly in loss-of-function-intolerant genes^20^. As expected, we also found that the burden of rare (MAF<0.001%) PTVs and damaging missense variants was negatively associated with imputed FIS (Supplementary Note 5; Supplementary Figures 10-11). Using combined FIS reduced the size of the confidence intervals compared to using observed FIS alone (red versus blue lines in Supplementary Figures 10-11), while not significantly changing the effect size for these two classes of variants. This motivated us to carry out gene-based rare variant association analyses on combined FIS to take advantage of the larger sample size.

We performed gene-based rare variant association tests with both observed FIS (N=273,091) and combined FIS (N=438,285) using Regenie^41^. We used both burden tests (which assume all included variants in the gene have the same effect size) and SKAT-O (which allows them to have different effect sizes including different directions of effect), and applied these to three combinations of rare (MAF<0.001%) variants: PTVs alone, damaging missense alone, and PTV+damaging missense combined (see Methods; Supplementary Table 11). We thus had twelve tests per gene (two phenotypes, two methods, three consequence classes), many of which were correlated, so we calculated the false discovery rate (FDR) across all conducted tests to account for multiple testing.

As expected, given the larger sample size, most genes were more significant using combined FIS (Figure 3A). Amongst the top genes (FDR<1%), effect sizes were highly correlated between observed and combined FIS, suggesting minimal biases being introduced by the imputation, with only one gene showing a notable difference in effect size (attenuation) after adding in imputed FIS (*ATP1A1*, a gene causing a dominant intellectual disability syndrome^42^) (Figure 3B). SKAT-O tended to improve power compared to burden tests (Supplementary Figure 12). Twenty-six genes passed FDR<1% (corresponding to p-value<5.03×10^-6^) (Table 1; Supplementary Table 12), of which all had negative effects on FIS. Fourteen are known DD-associated genes (defined as in the DDG2P knowledgebase), a highly significant enrichment given that only 2,047 out of 15,806 protein-coding genes we tested are in DDG2P (fold-enrichment = 4.18; Fisher’s exact test p=8×10^-8^). This fold-enrichment was highly significant across several FDR thresholds (Extended Data Figure 4A). It was also more pronounced than that observed for MAGMA-prioritized genes from the Combined FIS + COGENT GWAS, which were enriched ∼2-2.5-fold amongst the genes with rare variant associations (Extended Data Figure 4B). Among the twenty-six genes identified through rare variant testing, seven (*IGF1R, KCNMA1, GIGYF1, IPO9, CACNA1E, DPP8, NF1*) were also prioritized by gene-level enrichment for common variants in the Combined FIS + COGENT GWAS, but this overlap was not significant (Fisher’s exact test p=0.09).

**Figure 3.**
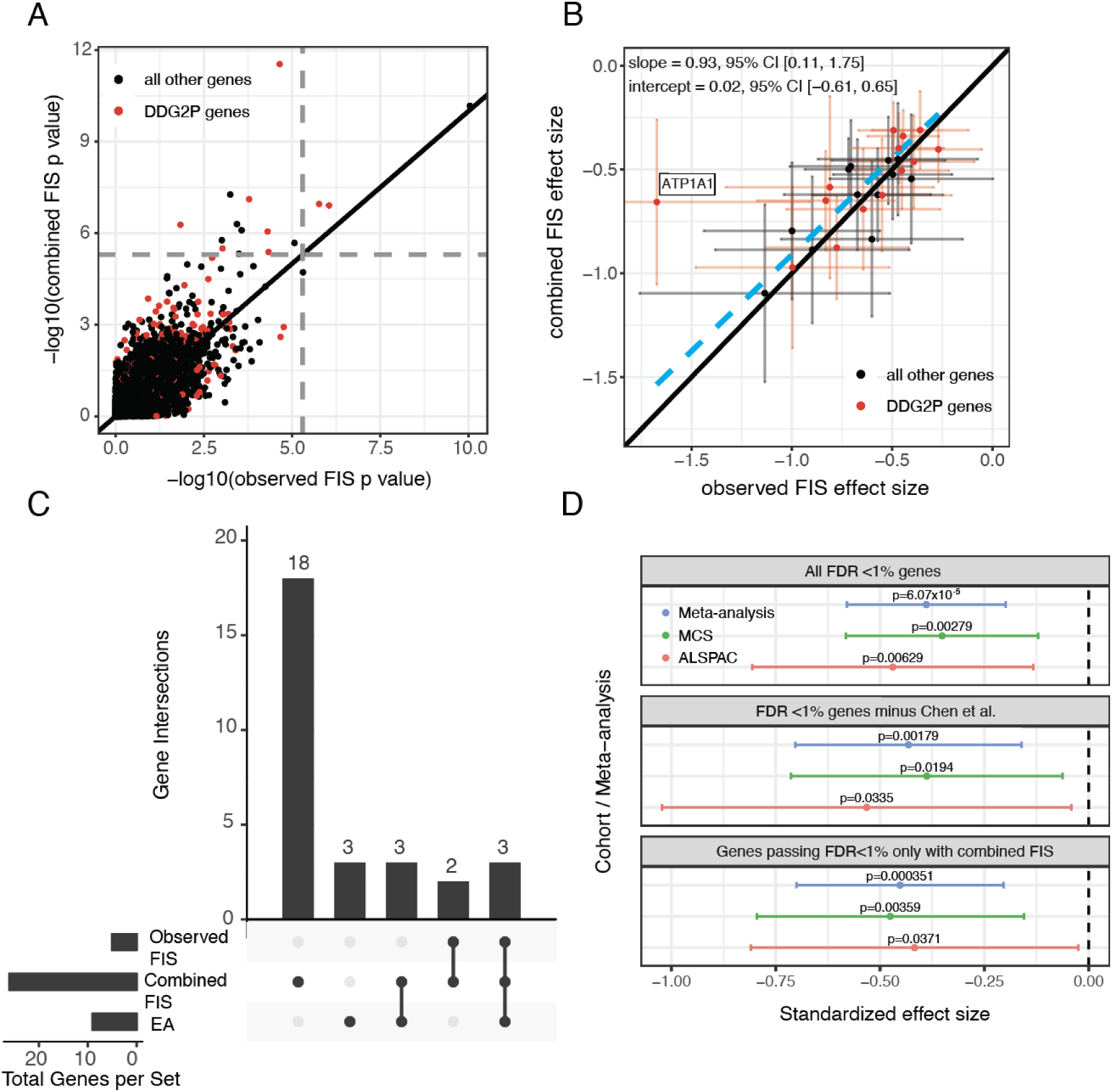
Results from gene-based rare variant association tests. **A)** -log10(p-value) obtained with observed FIS versus combined FIS from the burden test on PTVs. **B)** Effect sizes and 95% CIs obtained with observed FIS versus combined FIS from the burden test on PTVs, for genes passing FDR<1%. The dashed blue line shows the line of best fit from a Deming regression which takes into account the standard error of the effect size estimates. **C)** An upset plot showing the overlap of protein-coding genes prioritized by each phenotype analyzed by rare variants using FDR 1%. **D)** Standardized effect sizes and 95% confidence intervals for the association between PTV burden in various genesets in ALPSPAC, MCS, and an inverse variance weighted meta-analysis of both cohorts. “All FDR <1% genes” are the 26 genes that were significantly associated with observed and combined FIS as detailed in table 1. “FDR <1% genes minus Chen et al.” are the genes in the All FDR <1% genes, excluding those that were also found as associated at FDR <5% with any cognitive measure assessed in Chen et al. (18 genes). “Genes passing FDR <1% only with combined FIS” are the genes in the All FDR <1% genes that were only significant after additionally considering combined FIS rare variant analyses (21 genes). Gray dashed lines in (A) indicate the empirical p-value cutoff for an FDR <1% in our gene association tests (p<5.03×10^-6^).

**Table 1.** Results of gene-based rare variant tests from Regenie, for genes passing FDR<1%. Multiple tests were conducted per gene and the full results are given in Supplementary Table 12. This table gives the results of the most significant test for each gene. The “test type” column indicates whether the test was on observed or combined FIS, whether it considered PTVs alone or PTV+missense variants, and whether it was a SKATO or burden test. Note that SKATO does not produce an effect size. The “N. carriers” column indicates the number of individuals with rare variant carriers in the most significant test; for genes for which the most significant test was using combined FI, the number in parentheses is the number of carriers with an observed FI measure. NDC: neurodevelopmental condition.

| Gene | details of most significant test |  |  |  | Note |
| --- | --- | --- | --- | --- | --- |
|  | test type | N. carriers | beta | p-value |  |
| *IGF1R* | combined; PTV; SKATO | 57 (27) | NA | 6.9e-13 | DDG2P monoallelic and biallelic LoF<sup>43</sup> |
| *ANKRD12* | combined; PTV; burden | 154 (74) | -0.499 | 6.8e-11 | Associated with EDU (FDR<5%) and VNR (Bonferroni significant)<sup>20</sup> |
| *KCNMA1* | observed; PTV+missense; burden | 108 | -0.523 | 9.6e-11 | DDG2P monoallelic altered function<sup>44</sup> & biallelic LoF<sup>45</sup> |
| *CACNA1A* | combined; PTV; SKATO | 47 (29) | NA | 1.9e-9 | DDG2P monoallelic altered function<sup>46,47</sup>; associated with VNR (Bonferroni significant)<sup>20</sup> |
| *TNRC6B* | combined; PTV; SKATO | 61 (30) | NA | 5.1e-8 | DDG2P monoallelic LoF<sup>48</sup> |
| *ADGRB2* | combined; PTV; burden | 71 (35) | -0.622 | 5.4e-8 | Associated with EDU (Bonferroni significant)<sup>20</sup> |
| *KDM5B* | combined; PTV; SKATO | 219 (111) | NA | 6.1e-8 | DDG2P monoallelic and biallelic LoF<sup>49</sup>; associated with EDU, RT and VNR (Bonferroni significant)<sup>20</sup> |
| *GIGYF1* | combined; PTV; burden | 103 (64) | -0.505 | 7.7e-8 | DDG2P monoallelic LoF<sup>50</sup>; Associated with EDU (Bonferroni significant)<sup>20</sup> |
| *ATP1A1* | combined; PTV+missense; SKATO | 122 (71) | NA | 2.6e-7 | DDG2P monoallelic altered function<sup>51</sup> |
| *TLK1* | combined; PTV; SKATO | 28 (15) | NA | 2.8e-7 | *De novo* mutations reported in two NDC patients<sup>52</sup>; same protein family as *TLK2*, a DDG2P gene |
| *SLC8A1* | combined; PTV; burden | 19 (9) | -1.096 | 5.1e-7 | Associated with EDU (Bonferroni significant)<sup>20</sup> |
| *SPTBN2* | combined; PTV; burden | 139 (75) | -0.403 | 5.3e-7 | DDG2P monoallelic altered function<sup>53</sup> & biallelic LoF<sup>54</sup> |
| *PPFIA3* | combined; PTV; burden | 23 (15) | -0.972 | 8.8e-7 | DDG2P monoallelic altered function<sup>55</sup> |
| *MYCBP2* | combined; PTV+missense; burden | 169 (107) | -0.302 | 1.2e-6 | DDG2P monoallelic LoF (limited evidence)<sup>56</sup> |
| *IPO9* | combined; PTV; SKATO | 25 (17) | NA | 1.3e-6 | no clear link to NDCs |
| *CACNA1E* | combined; PTV; SKATO | 41 (24) | NA | 1.4e-6 | DDG2P monoallelic altered function<sup>46,47</sup> |
| *ANK3* | combined; PTV; burden | 76 (40) | -0.524 | 1.7e-6 | implicated in ID through both recessive and *de novo* variants<sup>57,58</sup>; GWAS hit for bipolar disorder<sup>59</sup> |
| *RB1CC1* | combined; PTV; SKATO | 47 (22) | NA | 2.0e-6 | Duplications associated with schizophrenia<sup>60</sup> |
| *RC3H2* | combined; PTV; burden | 32 (18) | -0.796 | 2.1e-6 | Associated with VNR (Bonferroni significant)<sup>20</sup> |
| *DPP8* | combined; PTV+missense; burden | 62 (33) | -0.507 | 2.3e-6 | no clear link to NDCs |
| *CHD9* | combined; PTV; SKATO | 78 (44) | NA | 3.6e-6 | *De novo* missense mutations reported in autism<sup>61-63</sup>; in the same family of chromatin remodelers as *CHD7*<sup>64</sup> and *CHD8*<sup>65</sup>, well-known NDC genes |
| *BCAS3* | combined; PTV; SKATO | 69 (38) | NA | 3.8e-6 | DDG2P biallelic LoF<sup>66</sup>; associated with EDU (Bonferroni significant)<sup>20</sup> |
| *NF1* | combined; PTV; SKATO | 121 (69) | NA | 4.01e-6 | DDG2P monoallelic LoF<sup>67</sup> |
| *ROBO2* | combined; PTV; burden | 49 (26) | -0.621 | 4.7e-6 | recurrent deletions reported in autism<sup>68</sup>; GWAS hit for expressive vocabulary in infancy<sup>69</sup> |
| *GABRA2* | combined; PTV+missense; SKATO | 80 (49) | NA | 4.8e-6 | DDG2P monoallelic altered function<sup>70</sup> |
| *PCLO* | observed; PTV; burden | 45 | -0.706 | 4.9e-6 | Involved in synaptic transmission; GWAS hit for depression<sup>71</sup> and post-traumatic stress disorder<sup>72</sup> |

Of the twenty-six significant genes passing FDR<1%, twenty-one achieved their lowest p-value using PTVs alone and five using PTVs plus damaging missense variants (Supplementary Table 12). Five genes passed FDR<1% using observed FIS alone (Figure 3C): *ANKRD12, PCLO, KDM5B, TNRC6B* and *KCNMA1*, of which the latter three are DDG2P genes. *ANKRD12* and *KDM5B* had been previously identified in a similar study by Chen et al. also using measured cognitive phenotypes in UKB^20^. (See Supplementary Note 5 for a more thorough comparison of our results with this paper.) An additional twenty-one genes passed FDR<1% only when analysing combined FIS, of which eleven are DDG2P genes and two additional genes (*RC3H2* and *SLC8A1*) were also reported by Chen et al.^20^. Amongst the eight genes passing FDR<1% which are neither DDG2P genes nor already reported by Chen *et al.*^20^, there are varying levels of prior evidence for their relevance to neurodevelopment (Supplementary Note 6), with two (*DPP8* and *IPO9*) having no prior evidence in humans.

The fourteen DDG2P genes passing FDR<1% have all been reported to cause syndromes with neurodevelopmental features, although in the case of *IGF1R* and *NF1*, these features are not always present^43,73^. Of these fourteen genes, four have previously only been reported to show dominant inheritance with a LoF mechanism (*TNRC68*^48^, *GIGYF1*^50^, *NF1*^67^ and *MYCBP2*^56^), one only to show recessive inheritance with a LoF mechanism (*BCAS3*^66^), and two to show both (*IGF1R*^43^ and *KDM5B*^49^). Seven have been reported to show dominant inheritance with an altered-function mechanism (*CACNA1A*^46,47^, *CACNA1E*^46,47^, *ATP1A1*^51^, *PPFIA3*^55^, *GABRA2*^70^, *KCNMA1*^44^ and *SPTBN2*^53^), of which the latter two also show recessive inheritance with a LoF mechanism^45,54^.

Given many of the significant genes are known to cause neurodevelopmental conditions, we wondered whether their associations with FI in UKB were driven by individuals recorded to have these conditions. Of the 2,045 individuals with rare PTV or damaging missense variants driving the signals for the significant genes in Table 1, 303 (∼15%) had healthcare records indicating intellectual disability, epilepsy, or autism. When we removed individuals with these codes and reran the rare variant tests, the p-values and effect sizes for most genes changed minimally (Extended Data Figure 5; Supplementary Table 13). All genes still passed multiple-testing correction except *CACNA1E* and *KCNMA1*, for which the p-values dropped from 1×10^-6^ to 6×10^-4^ and from 1×10^-10^ to 4×10^-3^ respectively. This suggests that rare damaging variants in most of these genes are associated with reduced FI even in individuals who do not have recorded diagnoses of neurodevelopmental conditions.

We sought to replicate these FIS-associated genes using recently-generated exome sequence data from two birth cohorts: ALSPAC (N=5283) and MCS (N=5621)^74^. Since we did not expect to have power to investigate any individual gene given the much smaller sample sizes of the replication cohorts, we collapsed the genes into three composite gene sets: the twenty-six genes passing FDR<1%, the eighteen of those that were not found by Chen et al. at FDR<5% with any of their cognitive phenotypes, and the twenty-one genes that passed FDR<1% in our analyses only after adding in the imputed FIS. Rare PTVs in all three of these gene sets showed significant negative effects on childhood cognition in both cohorts that became more significant after meta-analysis (Figure 3D). Thus, rare damaging variants in the novel genes discovered after incorporating imputed FI also associate with direct measures of cognitive ability.

Given the strong enrichment for DDG2P genes amongst our FIS-associated genes, we asked whether the twelve genes passing FDR<1% that were not DDG2P genes also showed direct evidence for association with NDCs. Specifically, we tested whether they were enriched for *de novo* mutations amongst ∼31,000 probands with NDCs from three published cohorts^22^. Across the twelve genes, we observed a significant enrichment of damaging *de novo* PTVs compared to the expectation under a null mutation model^75^ (observed/expected = 2.59, p=6.8×10^-4^, one-tailed Poisson test) but, reassuringly, no enrichment of *de novo* synonymous mutations (observed/expected = 1.08, p=0.40). This finding strongly suggests that damaging *de novo* variants in these FIS-prioritized genes elevate risk of NDCs.

Finally, we explored whether analysing combined FIS gave us more power for rare variant analyses than analysing educational attainment (EA) on essentially the same sample (N=434,146; slightly smaller N than for combined FIS due to missing EA values). We ran the same set of rare variant tests as described above for FIS (Supplementary Table 14) and then calculated the FDR across all tests. We found nine EA-associated genes passing FDR<1%, in contrast to the twenty-six that were found using observed or combined FIS (Figure 3C). Of these, six were also significant in our analysis of observed or combined FIS (*ANKRD12, KDM5B, ADGRB2, BCAS3, TNC6B* and *GIGYF1*), but three were not (*FOXK2, CHD6* and *CADPS2;* all with p<1×10^-3^ for all FIS tests) (Extended Data Figure 6). None of these three novel genes are known to be associated with NDCs. However, *CHD6* is in the same family of chromatin remodelers as the aforementioned NDC-associated genes *CHD7* and *CHD8*, and *FOXK2* encodes a transcription factor with roles in neural cell proliferation and neural precursor differentiation^76,77^. Notably, PTVs in *CADPS2* (calcium-dependent secretion activator 2) showed a *positive* association with EA (burden test; beta = 0.31, p-value = 2.5×10^-6^), in contrast to the other twenty-eight FDR-significant genes reported here, in which PTVs all showed *negative* effects on EA and/or FIS. PTVs in *CADPS2* showed no significant association with FIS (p>0.05 on all tests; for observed FIS, PTV burden beta=0.076; p=0.48). This gene also shows common variant associations with EA but not FIS, and colocalization with expression quantitative trait loci (eQTLs) in the brain suggests that decreased gene expression is associated with higher EA (Supplementary Note 7), concordant with our rare variant findings. In summary, incorporating imputed FIS boosted our power for rare variant-based gene discovery analyses over what could be achieved with EA alone. However, we did find several genes showing significant associations with EA but not FIS which may potentially affect EA by non-cognitive mechanisms or by influencing other aspects of cognition not captured by the UKB FI test.

## Discussion

In this study we evaluated and applied strategies for integration and imputation of FI phenotypes in UKB. We demonstrated that our approach yields a phenotype that is highly genetically similar to measured intelligence (Figure 1, Supplementary Figure 3). Through within-family GWAS, we showed that it also reduces ascertainment bias within the cohort (Figure 1F). Compared to previous common and rare variant-based studies of FI, we increased the sample size by ∼1.7-fold and ∼3.4-fold respectively. Adding imputed FIS to observed FIS increased power across multiple analyses: the number of lead SNPs in the GWAS increased by 1.6-fold (Figure 1D), MAGMA-prioritized genes by 1.3-fold (Figure 2A), polygenic score prediction accuracy in both population-based and within-family analyses in external cohorts by 1.15-fold (Figure 2C), and gene-level rare variant associations by 5.2-fold (Figure 3C).

Our study highlights the importance of evaluating the properties of an imputed phenotype using both phenotypic and genetic evidence. While our initial attempt at imputation produced a phenotype with reasonable accuracy (similar correlation with measured phenotypes [r=0.52] to the test-retest reliability of the measured phenotype [r=0.6]), it showed an excessively high genetic correlation with the “non-cognitive component” of educational attainment and with height (Supplementary Note 2; MegaFIS-vA in Supplementary Figure 4). We were able to use GWAS-by-subtraction to refine the list of phenotypic variables used in the imputation from 154 to 82 variables (Supplementary Note 2, Supplementary Figures 7 and 8), resulting in a phenotype that showed more appropriate genetic correlations with these variables (Supplementary Figure 4), with only a small decrease in accuracy (r=0.46 with measured phenotype). Thus, our results illustrate how including more correlated phenotypes in the imputation can sometimes increase accuracy at the cost of specificity.

Whereas observed FIS had a SNP h^2^ of approximately 0.20, this was somewhat lower (0.16) for our combined FIS GWAS (Figure 1). This reduction is likely due to the measurement error introduced by imputation, combined with our decision to standardise the observed and imputed scores separately before combining them (limitations discussed in Supplementary Note 3). These both decrease the relative variance explained by genetic associations, although the attenuation of the estimated heritability was modest. Reassuringly, the genetic correlation profiles of the observed and combined FIS are highly concordant. Critically, the downstream analyses, such as the external cohort PGI and tissue enrichment (Figure 2), suggested that the incorporation of imputed individuals did increase power despite the slightly lower SNP heritability.

We showed that incorporating imputed FI scores into rare variant analyses enhanced power for gene discovery, allowing us to expand the number of significant gene-level associations from five to twenty-six (Table 1), without changing effect size estimates (Figure 3AB). Fourteen of these significant genes are DDG2P genes (a four-fold enrichment; p=8×10^-8^) and well-established causes of neurodevelopmental conditions. However, we show that, with the exception of two genes, these associations are not substantially changed by excluding individuals with evidence of such conditions in their healthcare records. This suggests our findings are not simply recapitulating the known effects of variants in these DD-associated genes, and that rare variants in these genes can affect a key endophenotype for neurodevelopmental conditions (cognition) without necessarily pushing individuals over the diagnostic threshold. Given our estimated effect sizes for rare variants in these genes on FI (∼0.25-1 SDs), we would not expect all carriers of such variants to have clinically recognised intellectual disability. It is likely that polygenic background^78^ and potentially environmental factors modify the penetrance of rare damaging variants in these genes for clinical phenotypes. Such modifying factors likely also explain the observation that loss-of-function variants in two of the genes we found (*KDM5B* and *IGF1R*) can show both dominant and recessive inheritance patterns^43,49^. This is an unusual property observed for only 31 of 2447 (1.2%) DDG2P genes to date; observing this in 2 of 14 DDG2P genes that passed FDR<1% here represents a significant enrichment (Fisher’s exact test p=0.01; odds ratio=12.9). These findings support those of Fridman et al. who found that heterozygous carriers of pathogenic variants in recessive intellectual disability genes had, on average, lower educational attainment in UKB^79^. They also support the notion that there are probably more DD-associated genes than currently appreciated in which the “dominant/recessive” dichotomy does not accurately reflect a gradient of heterozygous effects^80^.

Our rare variant association findings also expand our knowledge of the mechanism of action for several of these DD-associated genes. Specifically, for five genes (*CACNA1A*^46,47^, *CACNA1E*^46,47^, *ATP1A1*^51^, *PPFIA3*^55^ and *GABRA2*^70^), only dominant gain-of-function variants were previously known to disrupt neurodevelopment, and these typically lead to epilepsy. Our findings show that loss-of-function variants in these same genes also reduce cognitive ability. Furthermore, we find twelve genes significantly associated with FI that are not well-established NDC genes (i.e. not in the DDG2P knowledgebase). Several of these are paralogs of DDG2P genes (e.g. *ANKRD12* is a paralog of *ANKRD11*, and *TLK1* of *TLK2*; *CHD9* is a paralog of both *CHD7* and *CHD8*). Others have been previously linked to autism (*ROBO2*^68^) and schizophrenia (*RB1CC1*^60^) through copy number variants, and/or are GWAS hits for brain-related phenotypes, such as *ROBO2* for infant vocabulary^69^ and *PCLO* for depression^71^ and PTSD^72^ (Supplementary Note 6). Finally, to our knowledge, two of the novel genes we find (*DPP8* and *IPO9*) have no prior evidence for involvement in cognition or brain phenotypes in humans, although the homozygous mouse knockouts do show relevant phenotypes (behavioural anomalies for *Dpp8*^81^, brain structural anomalies for *Ipo9*^82^).

Importantly, we showed that bringing in imputed FI increased the power for rare variant-based gene discovery beyond what we could achieve using EA alone (Extended Data Figure 6). This may be due to several reasons, critically that EA may have a lower rare variant heritability than combined FIS; certainly the twin-heritability (52% versus 67%)^9^ and the common SNP heritability (14.3% versus 18.6%)^17^ have been reported to be slightly lower for EA than cognitive performance. Notably, the genetic correlation estimated from direct effects between EA and cognitive ability is only 0.47^17^, implying that these two phenotypes have considerably distinct genetic contributions. This supports our findings that the genes that show rare variant associations with EA only partially overlap with those for FI, including *CADPS2*, in which PTVs were associated with increased EA but showed no association with FI. This gene encodes a calcium-binding protein involved in the release of neurotrophins^83^. Prior findings involving common variants near this gene support the notion that loss-of-function/reduced expression is associated with higher EA (see Supplementary Note 7 for details and further discussion).

This work has several limitations. Although we show that adding in imputed FIS increases the power for common variant analyses compared to using measured FIS alone, prior results suggests that the published GWAS for the cognitive component of EA produced by Genomic Structural Equation Modelling^31^ may still be a better-powered GWAS for cognitive ability. Specifically, a PGI based on the “CogEA” GWAS explained slightly more variance in ALSPAC than one based on our combined FIS GWAS (9.6% versus 8.2% for population effect; 2.9% versus 2.6% within-family; Supplementary Table 9). Nonetheless, we have demonstrated the substantial value of using combined FIS to boost power to find genes in which rare variants are associated with FI, with a few caveats. We cannot rule out that the associations between rare variants in the known DD-associated genes and FI are driven by people with NDC diagnoses, since these may be incompletely ascertained in the health records. Also, although we showed that these genes replicate in aggregate in ALSPAC and MCS, we have not attempted replication of individual genes because no cohort exists with sufficient power. Another limitation is that, due to the computational burden of applying multiple imputation, we analyzed only a single imputed dataset. This approach ignores imputation uncertainty, potentially biasing effect size estimates and standard errors. However, the imputed phenotype correlated approximately 0.50 with the masked observed measure, below the test–retest reliability (∼0.60), and preserved expected patterns of downstream genetic associations with other traits (e.g. Figure 1C), suggesting that the imputation introduced predominantly classical measurement error. Such measurement error likely biases effect size estimates toward zero and reduces statistical power, but is unlikely to inflate false positives or alter the direction of observed associations. Reassuringly, we find that effect size estimates are highly correlated in various analyses using observed versus combined FIS (e.g. Figure 3B), and we replicate both the common and rare variant signals using direct measures of intelligence in external cohorts through a variety of analyses. Finally, another limitation is that this work was limited to individuals with genetically-inferred European ancestries.

In summary, we have demonstrated how integrating and imputing fluid intelligence measures in UKB improves power for genetic discovery while reducing ascertainment bias. Since studies of this kind are often of interest to broader audiences, we have provided a lay summary and FAQ document in the Supplement to explain our motivation and how our results can and cannot be interpreted. Our findings reinforce the idea that rare variants with moderate effects on cognition are more pervasive than previously appreciated, and that integrating carefully imputed phenotypes can provide a powerful avenue for identifying novel contributors to neurodevelopmental processes, even among individuals without diagnosed conditions. Future studies should seek to integrate these findings with those from clinical cohorts to increase power to find new genes associated with neurodevelopmental conditions. Furthermore, studies in populations of non-European ancestries will be critical to explore the generalisability of these findings. Finally, this work motivates the broader use of phenotype imputation for genetic studies in large biobanks.

## Acknowledgements

This research was conducted using the UK Biobank Resource under application numbers 40310 and 44165.

1. This work used the Dutch national e-infrastructure with the support of the SURF Cooperative using grant no. EINF-7294
2. This research was funded in part by Wellcome (grant no. 220540/Z/20/A, “Wellcome Sanger Institute Quinquennial Review 2021–2026”). For the purpose of open access, the authors have applied a CC-BY public copyright license to any author accepted manuscript version arising from this submission.
3. K.J.H.V. is supported by the Foundation Volksbond Rotterdam.
4. A.A. is supported by the Amsterdam UMC Fellowship.
5. ALSPAC: We are extremely grateful to all the families who took part in ALSPAC, the midwives for their help in recruiting them, and the whole ALSPAC team, which includes interviewers, computer and laboratory technicians, clerical workers, research scientists, volunteers, managers, receptionists and nurses. The UK Medical Research Council and Wellcome (Grant ref: 217065/Z/19/Z) and the University of Bristol provide core support for ALSPAC. This publication is the work of the authors and Hilary Martin will serve as a guarantor for the contents of this paper. Genome-wide genotyping data was generated by Sample Logistics and Genotyping Facilities at the Wellcome Sanger Institute and LabCorp (Laboratory Corporation of America) using support from 23andMe. This research was specifically funded by the UK Medical Research Council and Wellcome (Grant ref: 076467/Z/05/Z) and the Department for Education and Skills (Grant ref: EOR/SBU/2002/121). A comprehensive list of grants funding is available on the ALSPAC website: http://www.bristol.ac.uk/alspac/external/documents/grant-acknowledgements.pdf
6. MCS: We are grateful to the Centre for Longitudinal Studies (CLS), UCL Social Research Institute, for the use of these data and to the UK Data Service for making them available. However, neither CLS nor the UK Data Service bear any responsibility for the analysis or interpretation of these data.

## Author contributions

DMvdB, WH, DSM analysed the data and drafted the paper. SJL carried out the *de novo* analysis in the DD cohorts and helped supervise the rare variant work. PD and KW contributed to quality control. KV and DJAS contributed to supervision. EB contributed to the FAQ and the writing. MEH, AA and HCM jointly directed the work and helped write the paper. All authors read and approved the draft.

## Data availability statement

UK Biobank data are available to researchers upon application (https://www.ukbiobank.ac.uk/enable-your-research/apply-for-access). The imputed fluid intelligence phenotype is not publicly available due to participant privacy and UK Biobank’s data sharing policies but can be regenerated by approved researchers using the code provided on GitHub (https://github.com/dmvandenberg/UKB-FI-Imputation). Summary statistics from population and within-family based GWAS of combined FIS, measured FIS, Imputed FIS, and the meta-analysis of combined FIS and COGENT will be made available via the GWAS Catalog upon peer reviewed publication. Full results of rare variant analyses, including gene-level association statistics are available in the Supplements.

## Extended Data Figures

**Extended Data Figure 1.**
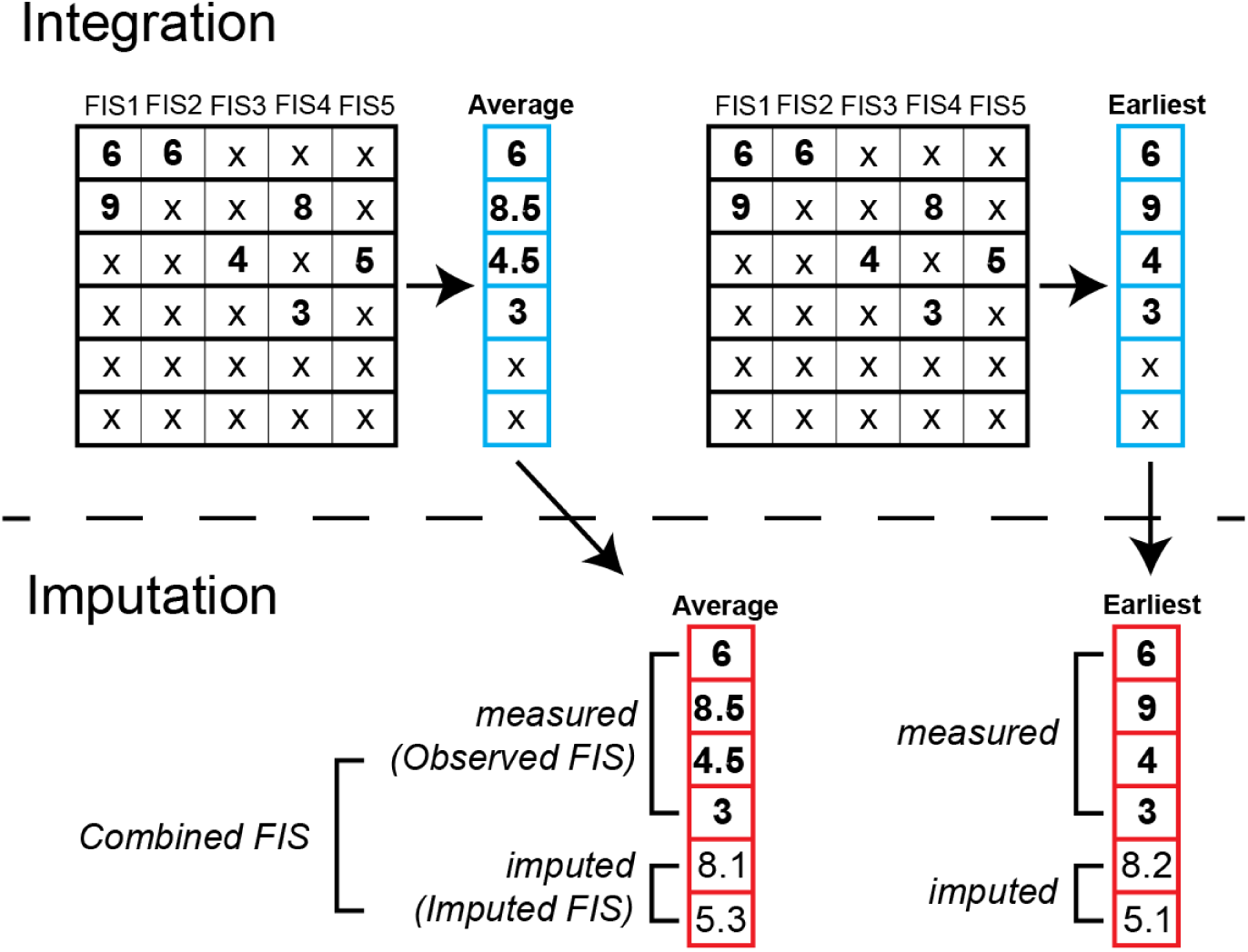
Schematic illustrating the strategies for integration and imputation of fluid intelligence described in the main text. See also Supplementary Figure 5 which illustrates additional strategies that were evaluated as described in Supplementary Note 2. The top panel shows how the summary measures are derived from the five different FI tests (“integration”) and what is included in the imputation (blue grid) along with the selected imputation variables. Although this figure shows raw FIS scores for clarity, the actual FIS scores were transformed prior to imputation (See Methods). In the average FIS approach, scores for an individual (row in grid) are combined into one average per individual, then individuals without any FI scores are imputed. In the earliest FIS approach, the first score in time is selected, and individuals without any scores are imputed. The bottom panel shows what is analyzed in Figure 1 (in red).

**Extended Data Figure 2.**
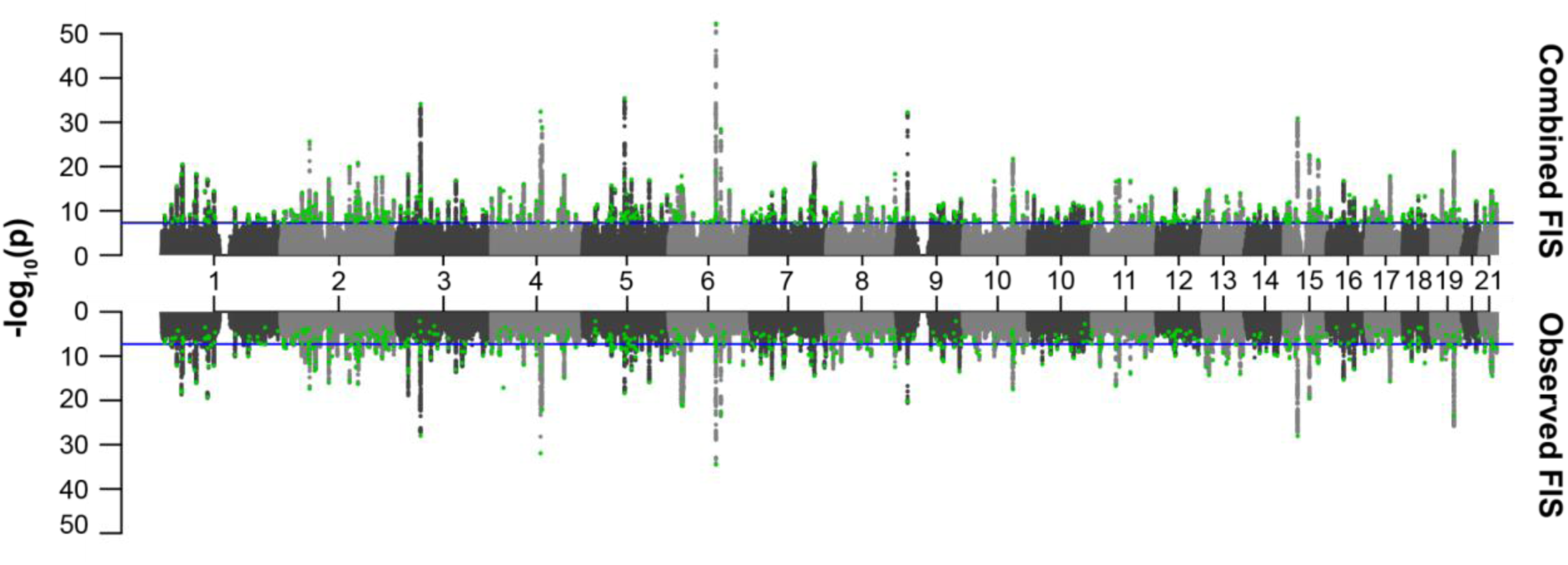
Combined FIS vs. observed FIS Miami plot: Manhattan plots showing results from the population-based GWAS of combined FIS (top) versus observed FIS (bottom). SNPs highlighted in green are lead SNPs identified in combined FIS using FUMA. The x-axis shows the chromosomes and the y-axis the -log10(p) for the SNP associations. The blue line in both plots indicates the genome wide significance threshold p<5×10^-8^. For observed FIS, highlighted SNPs below the blue line (non-significant) indicate SNPs that were identified as lead SNPs as a result of adding imputed values in combined FIS.

**Extended Data Figure 3.**
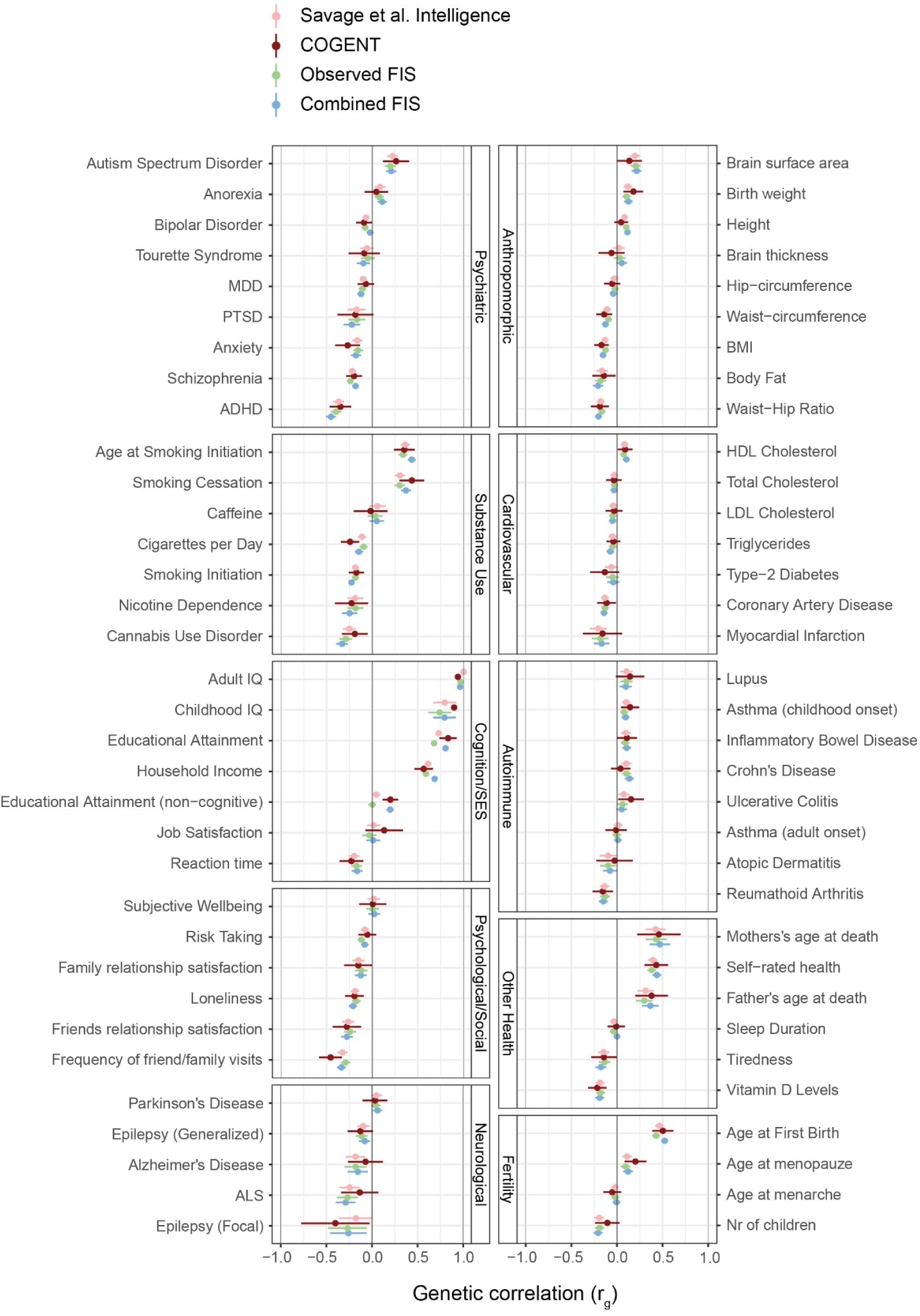
Genetic correlations of 68 traits with Savage et al. Intelligence, COGENT, Observed FIS, and Combined FIS. Error bars show the 95% confidence interval. See **Supplementary Table 3** for numeric values and information about the source GWAS.

**Extended Data Figure 4.**
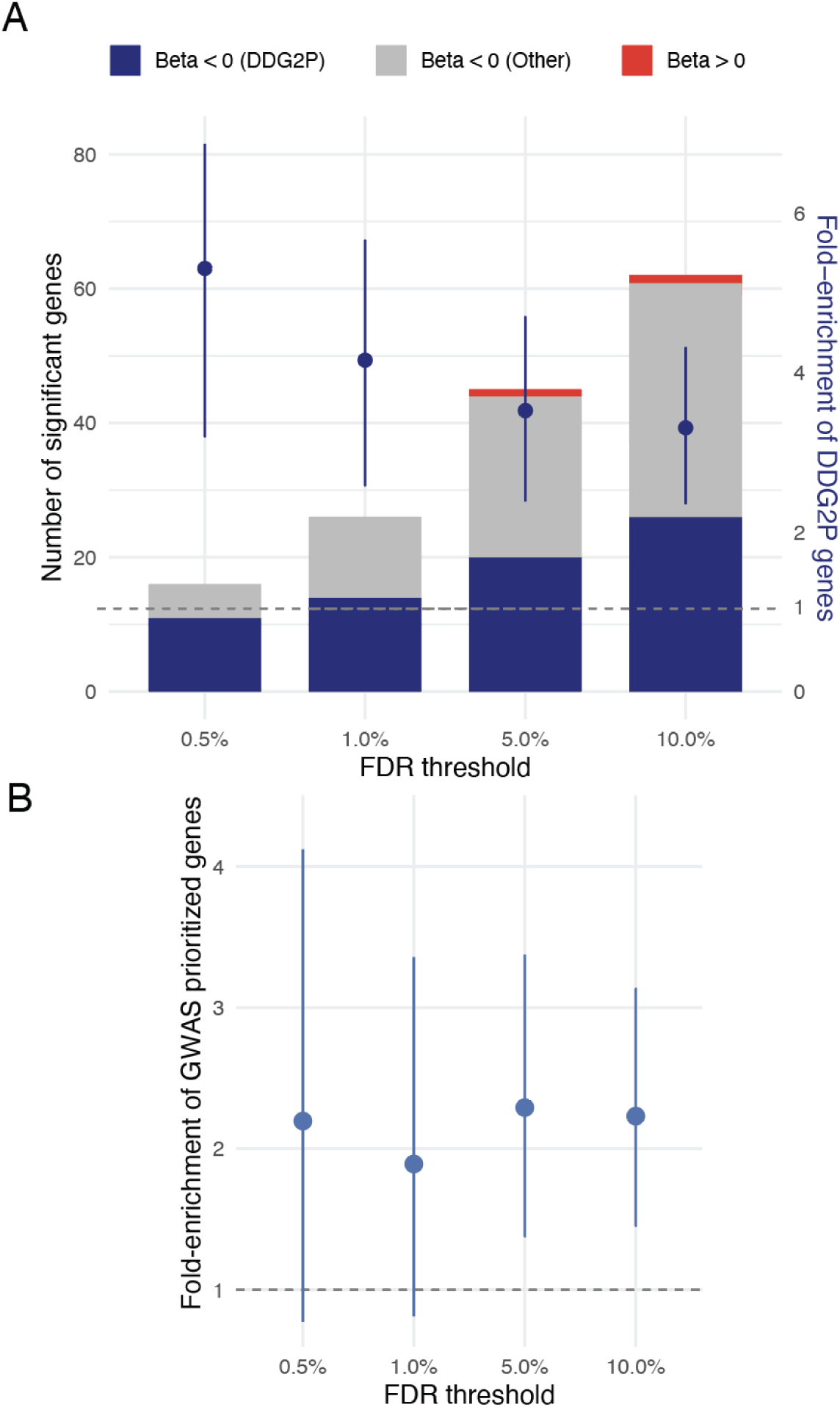
Assessing overlap between developmental disorder-associated genes and those prioritized by Combined FIS + COGENT GWAS versus genes identified by rare variant burden tests. **A)** Stacked bar plots showing the number of genes significant at each rare variant FDR threshold. Dark blue segments are genes with PTV and PTV + MIS burden beta < 0 that are in the DDG2P list; gray segments are genes with PTV and PTV + MIS burden beta < 0 that are not in DDG2P; red segments are all other significant genes. Overlaid are point estimates and 95% CI from a hypergeometric test for the fold-enrichment of DDG2P membership in each FDR-thresholded set relative to the genome-wide background, calculated as (|FDR gene set & DDG2P| / |FDR gene set|) / (|DDG2P| / |all genes|). **B)** Fold-enrichment of genes implicated by the rare variant association tests at the indicated FDR threshold amongst MAGMA-prioritized genes passing the 1% FDR threshold from the Combined FIS+COGENT GWAS (as in Figure 2A).

**Extended Data Figure 5.**
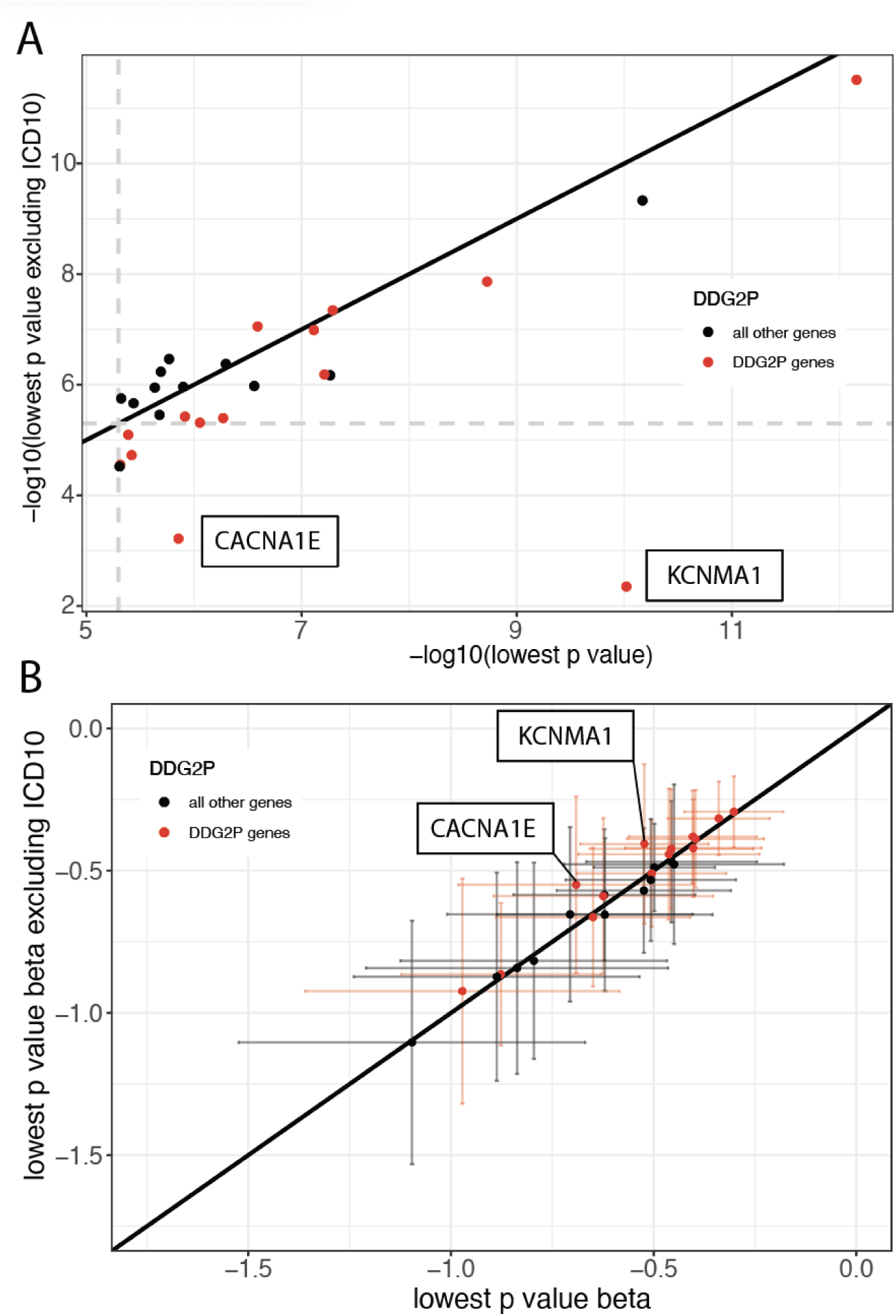
Results from gene-based rare variant burden tests before and after removing individuals with ICD-10 code indications for intellectual disability, autism, or epilepsy. **A)** On the x-axis is the -log10(p) for the most significant p-value of a given gene amongst the 26 genes passing (FDR<1%) across the twelve gene-based rare variant tests; on the y axis is the -log10(p) for the same test after removing individuals with ICD-10 indicating intellectual disability, autism, or epilepsy. **B)** On the x-axis is the beta from the gene burden test using the variant consequence class leading to the lowest p-value per gene (95%CI)); on the y-axis is the beta from the same test after removing individuals with ICD-10 indicating intellectual disability, autism, or epilepsy. Solid black lines indicate the y=x line. Gray dashed lines indicate the empirical p-value cutoff for an FDR <1% in our gene association tests (p<5.03×10^-6^).

**Extended Data Figure 6.**
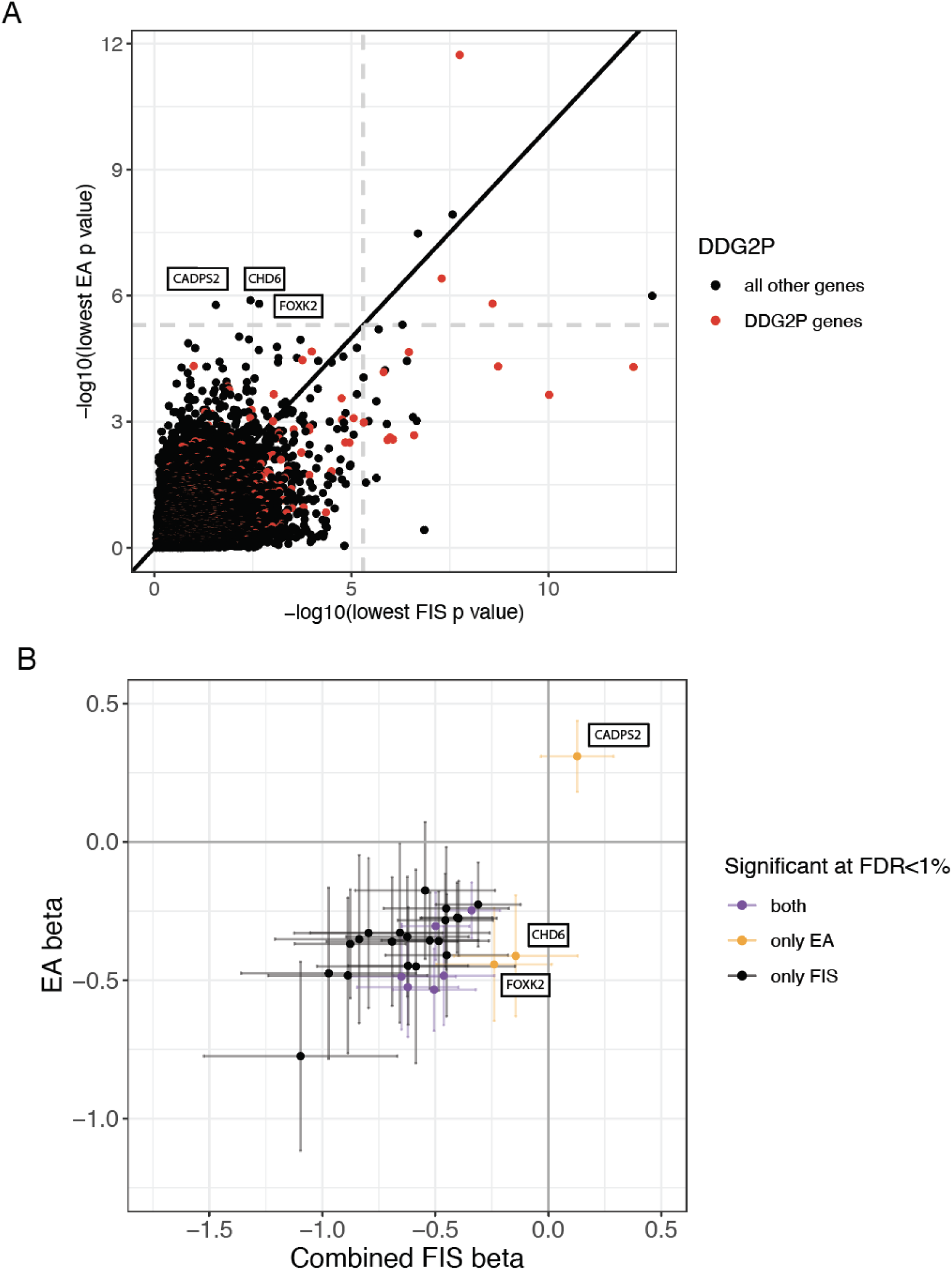
Results from gene-based rare variant burden and SKATO tests using FIS phenotypes versus educational attainment. **A)** On the x-axis is the -log10(p) for the lowest p-value attained in any test for any of the FIS phenotypes and on the y-axis is the -log10(p) for the lowest p-value attained in any test for educational attainment. Gray dashed lines indicate the empirical p-value cutoff for an FDR <1% in our gene association tests (p<5.03×10^-6^). **B)** On the x-axis is the beta for the pLoF burden test on combined FIS with 95% CIs and on the y-axis is the pLoF burden test on EA, with points colored based off whether the gene was significant at 1% FDR in either FIS only, EA only, or both.

## Methods

### Sample selection

We conducted all our analyses only in individuals who genetically cluster with those of European ancestries (N=455,943). This was inferred by first projecting PCs from 1000 Genomes onto the UKB participants and excluding participants who did not cluster with the European populations from 1000 Genomes. After this, another PCA was conducted to capture ancestry differences within the genetically more homogeneous individuals with European/British ancestries^84^.

### FIS score transformations

To correct for age decline in FI over time and differences in scales between FI tests, we transformed the FIS measures. For each FIS separately we ran a regression model: *FIS* ∼ *age* + *age*^2^, extracted the residuals and added the intercept to allow for differences in means between measures. The age used was the participant’s age at the time of the respective measurement, which was either obtained from UKB variable ‘age attended assessment center’ (Data-Field 21003) or approximated from ‘When fluid intelligence test completed’ (Data-Field 20135) using the participants’ birth year and month.

For the online measures (FIS4 & FIS5) we did an additional transformation, setting all scores of 14 to 13, prior to running the regression model. This broadly aligns the scales of the online measures (14 questions) with those of the in-person test (13 questions) (Supplementary Note 1).

### Imputation

The R package SoftImpute^85^ was used to impute FIS. SoftImpute is a matrix completion algorithm that approximates missing values by identifying and leveraging patterns in the available data. It starts with an initial guess for the missing values, then iteratively refines this guess by applying a soft-thresholded Singular Value Decomposition (SVD) on the complete matrix. The main parameters specified in SoftImpute are rank and lambda. Rank determines the maximum number of factors the method uses to represent the data, and should always be set to at most *N_variables_* − 1. Lambda controls the amount of smoothing applied during imputation. A high lambda aims at lower model complexity, making it less sensitive to noise and more prone to overlook nuances. Ideally, lambda is set to be slightly less than the rank set.

#### Variable selection

##### FIS vA

We first selected 152 UKB phenotypes amongst those collected at the initial assessment centre visit that showed at least nominally significant correlation (p<0.05) with observed fluid intelligence, focusing on FIS1 since it had the largest sample available at the time. For continuous phenotypes, we chose those with an absolute phenotype correlation (|r_pheno_|) with FIS1 greater than 0.05 at a significance threshold of p<0.05. Categorical phenotypes were first classified into ordinal and nominal types and then further evaluated. For ordinal phenotypes, we verified whether the values could be interpreted as a quantitative variable, reordered them if necessary, and then retained those with |r_pheno_| greater than 0.05. We converted nominal phenotypes with N categories into N-1 binary variables, after which we calculated the r_pheno_ between each of those binary variables and FIS1. We only included variables with two or more categories having |r_pheno_| greater than 0.1. We excluded phenotypes that are components of any FIS measure. In total, we selected 90 continuous phenotypes, 59 ordinal phenotypes, and 3 nominal phenotypes (Supplementary Table 4). For the three nominal phenotypes, we selected the categories exhibiting a strong correlation with FIS1 (|r_pheno_| > 0.1), coded them as additional binary phenotypes, and removed the original phenotypes, ultimately leading to a set of 154 variables used as our initial set. Finally, we coded all missing values (e.g., “Preferred not to answer” or “unknown”) as NA. Based on the 154 resulting phenotypes, we ran imputation FIS vA with SoftImpute parameters: rank=150, lambda=120.

##### FIS vB and FIS vC

In imputation vB and vC we narrowed the initial selection of variables down to phenotypes that correlate more specifically with cognitive signal. To examine which phenotypes fall within this criterion we derived a GWAS of the ‘non-cognitive component of imputed FIS’ (NonCog-iFIS) from FIS-vA. To do this we applied a GenomicSEM model (Supplementary Figure 7) for GWAS-by-subtraction as applied in Demange et al.^31^. GenomicSEM is a R-package that allows fitting structural equation models on summary statistics from GWAS. In GWAS-by-subtraction models, a model is fitted that includes two phenotypes (GWAS) and two latent factors. The first latent factor represents the commonalities between phenotypes by regressing both phenotypes on this latent factor. The second latent factor comprises the genetic variance unique to one of the phenotypes. This is achieved by regressing the remaining genetic variance for the phenotypes on the latent factor after regressing out the variance captured in the first latent factor. Subsequently both latent factors can be regressed on individual SNPs to obtain GWAS. In our analysis we ran a GWAS-by-subtraction model using Savage et al. Intelligence^14^ and imputed values from FIS vA as phenotypes. The model allows us to capture the genetic variance unique to imputed FISvA in the ‘NonCog’ latent factor and subsequently regress individual SNPs on this factor to obtain the NonCog-iFIS GWAS.

We then computed genetic correlations between each of the 154 imputation variables and Savage et al. Intelligence, and the derived NonCog-iFIS GWAS (Supplementary Table 4). We retained phenotypes to use for imputation vB if the absolute genetic correlation was stronger with Intelligence than with NonCog-iFIS (Supplementary Figure 8), resulting in 82 phenotypes being selected.

For imputation vB and vC, we used the same imputation method as in vA, but we adjusted our SoftImpute parameters (rank=80, lambda=70) due to the reduced number of selected phenotypes. We also explored the effects of different imputation parameters on imputation accuracy, but found that rank=80 and lambda=70 achieved the highest imputation accuracy (Supplementary Figure 9, left).

#### Removing outliers

After each imputation, we identified then removed outliers using the criterion:

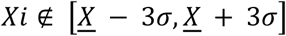

Where:

- *Xi*represents an imputed score
- <u>*X*</u> represents the mean of the measured score
- *σ* represents the standard deviation of the measured score

In other words, we removed individuals whose imputed score was more than three standard deviations from the mean of the observed scores. This results in slightly differing sample sizes across the imputation approaches (Supplementary Table 6).

#### Evaluating accuracy

To evaluate the imputation accuracy, we randomly selected 50,000 participants as the evaluation set. We introduced ‘synthetic missingness’ by setting FIS measures for these participants to be missing. After imputation, we examined the Pearson correlation between imputed FIS values and the original measures for our evaluation set and defined that correlation as the imputation accuracy. Only participants who have the targeted FI measure were included in calculation of accuracy for each FI measure (see Supplementary Table 5).

We also examined whether the imputation was more accurate in different parts of the phenotype distribution. We specifically focused on imputation vC and classified observed values of the evaluation set into low, medium and high terciles. We then examined the imputation accuracy within each tercile using the same strategy as above. We found the imputation accuracy to be higher in the low and high terciles than in the medium tercile (Supplementary Figure 9, right).

### Combining measured and imputed FI scores

Combining imputed and measured FIS was done by mega-analysis in all approaches. Prior to mega-analysis, imputed and measured values were scaled separately to have mean 0 and a standard deviation of 1. We also evaluated combining imputed and measured values through meta-analysis (Supplementary Note 3, Supplementary Table 6).

### GWAS

For the Genome-Wide Association Studies (GWAS), we used Haplotype Reference Consortium^86^ (HRC) imputed SNP genotypes, and ran ‘*--fastGWA-lmm*’ from Genome-wide Complex Trait Analysis (GCTA, version 1.92.3). Quality control parameters were kept at default values filtering MAF < 0.0001 and missingness rate > 0.1. We set the ‘*--est-vg*’ parameter to ‘*HE*’ to estimate the genetic variance component using Haseman-Elston regression, and specified covariates to account for using the ‘-*-qcovar*’ and ‘*--covar*’ parameters. The quantitative covariates (‘-*-qcovar*’) included were 25 genetic principal components that capture ancestry differences within European individuals^84^, age (at time of respective FIS measurement), age^2^, age*sex, and age^2^*sex. The other covariates (‘-*-covar*’) included the array used to measure each individuals’ genotype and sex as a binary variable. For the average FIS approach we computed the average age across measures and used that as the age covariate. For individuals who had their average FIS imputed, their age was set to the UKB variable ‘age initial assessment visit’ (Data-Field 21003, Instance 0).

### Meta-analysis

For meta-analyses we used METAL^87^ with the ‘STDERR’ approach. This weights effect size estimates by the inverse of corresponding standard errors. We only include SNPs with N > 10,000.

### Genetic correlations and SNP h^2^

Linkage disequilibrium score regression (LDSC)^35^ was used to compute genetic correlations and SNP heritabilities. This tool requires munged (parsed) sumstats. In munging we aligned SNPs with those in the HapMap3^88^ set using the ‘--merge-alleles’ flag. Heritabilities and genetic correlations were computed using the ‘--h2’ and ‘--rg’ flag respectively, with default parameters. To estimate the genome-wide correlation between the direct effects and NTCs, we used the SNIPAR package correlate.py script^36^.

### Identifying lead SNPs

The online platform for Functional Mapping and Annotation of Genome-Wide Association Studies (FUMA)^34^ SNP2GENE was used to identify lead SNPs. First, independent significant SNPs are identified (P < 5e^-8^, r^2^ < 0.6). Subsequently, identified significant SNPs are designated lead SNPs if they are independent from each other at a second threshold of r^2^ < 0.1.

### Gene prioritization and tissue expression

MAGMA^89^ (as implemented in FUMAs SNP2GENE process) was used for gene prioritization and tissue expression analyses using the results from the population-based GWAS. MAGMA aggregates SNP-level data into gene-level data and performs a gene-based association test to identify significantly associated genes. The gene window was kept at 0kb, restricting analysis to SNPs located within a gene. Resulting gene-based p-values were downloaded and FDR corrected using the Benjamini-Hochberg procedure. To obtain our set of prioritized genes, we selected protein coding genes with an FDR < 1% that were also MANE select transcripts^90^. We then applied the SNP2GENE process in FUMA to test whether the significant genes were enriched in particular tissues; this takes the gene-level p-values from MAGMA as input. We used expression data from 54 tissues from GTEx v8^91^ as reference data.

### Within-family GWAS

We conducted within-family GWAS in UKB using the SNIPAR package^36^ in individuals with European ancestries. SNIPAR leverages the presence of genotyped first-degree relatives to impute missing parental genotypes, allowing for their downstream use to conduct within-family GWAS. We estimated pairwise kinship coefficients using KING^92^ and used default parameters to conduct the within-family GWAS using scripts provided in SNIPAR. We controlled for the same covariates as in the population GWAS SNP heritability estimates were estimated using LDSC as described above.

### Whole-exome sequencing data in UKB and quality control

Whole exome sequencing data was generated at the Regeneron Genetics Center (RGC), and the sequencing procedure has been described in previous studies. We used custom applets to perform quality control for the whole exome sequencing data of 469,836 participants within the UKB research analysis platform. First, we used bcftools norm to split and left-align multi-allelic variants in the population-level Variant Call Format (VCF) files into separate alleles. Next, we performed genotype-level filtering using bcftools filter separately for Single Nucleotide Variants (SNVs) and Insertions/Deletions (InDels). Specifically, SNV genotypes with a depth below 7 and genotype quality below 20, or InDel genotypes with a depth below 10 and genotype quality below 20, were set to missing. We also applied a binomial test to check for an expected alternate allele contribution of 50% for heterozygous SNVs, and SNV genotypes with a binomial test p-value ≤ 0.0001 were set to missing. Finally, we recalculated the proportion of missing genotypes for each variant and excluded all variants with more than 50% missingness.

Next, we annotated the variants using the ENSEMBL Variant Effect Predictor (VEP) v104 with the --everything flag. For each variant, we prioritized a single ENSEMBL transcript based on whether the transcript was protein-coding, MANE Select v0.97, or the VEP canonical transcript. The variant consequence was prioritized based on severity as defined by VEP. Following annotation, we grouped stop-gained, frameshift, splice acceptor, and splice donor variants into a single Protein Truncating Variant (PTV) category. Missense and synonymous variant consequences were defined according to VEP criteria, and only autosomal variants within ENSEMBL protein-coding transcripts were retained for further analysis.

We further filtered the variants by minor allele frequency (MAF), retaining only those with a MAF lower than 0.001%. These variants were annotated with LOFTEE, REVEL, Alphamissense, and MPC for further filtering. For PTVs, only high-confidence PTVs defined by LOFTEE were retained. For missense variants, a damaging missense variant set was created by including variants with Alphamissense scores above 0.56, REVEL scores above 0.5, and MPC scores greater than 2.

### Exome-wide burden tests of rare coding variants

To examine the association between FIS and the burden of rare coding variants, we counted the number of rare PTVs, missense variants, damaging missense variants (as described above), and synonymous variants both exome-wide and in high loss-of-function-intolerant genes (pLI > 0.9). The variant burden was then used as the predictor variable in linear regression models, with FIS (observed, imputed, and combined) as the response variables. The models were run in unrelated participants with European ancestries (N=328,795). We controlled for the top 25 principal components (PCs), as well as age, sex, age², and the interactions between age and sex, and age² and sex^84^.

### Gene-based burden test of rare coding variants

We performed gene-based burden tests on observed and combined FIS and EA using a two-step regression analysis in Regenie. As Regenie accounts for relatedness and population structure, the gene-based tests were performed in all participants with European ancestries (N=438,285; note this is lower than the sample size used for common variant analyses as not all participants had exome data). In the first step, Regenie fits a stacked block ridge regression to produce a leave-one-chromosome-out (LOCO) genetic prediction of the focal phenotype. The association test is then carried out in the second step by fitting regression models conditioned on the LOCO predictions. For both steps, we adjusted for sex, age, age², sex-by-age interaction, sex-by-age² interaction, the top 25 principal components (PCs), and recruitment centers (as categorical variables) to control for population structure and age. The FIS scores were rank-based and inverse-normal transformed, as recommended by Regenie. EA was coded as in Okbay et al. by using individual reports of highest attained qualification, with “College or University degree” counting as 20 years, “other professional qualifications” as 15, “A levels/AS levels or equivalent” as 13, “O levels/GCSEs or equivalent” and “CSEs or equivalent” as 10, and “None of the above” as 7^93^. We ran both burden tests and SKAT-O tests on three consequence classes: PTV, damaging missense, and PTV+damaging missense. Thus, for FIS we conducted twelve tests per gene (two phenotypes, two statistical methods, three consequence classes). To account for multiple testing, we calculated the false-discovery rate (FDR) using the Benjamini-Hochberg method across the vector of all p-values, and considered genes passing FDR<1% as ‘significant’. For EA, we conducted six tests per gene as we only considered a single phenotype, and similarly accounted for multiple testing by calculating the FDR across a vector of all p-values, and considering genes passing FDR<1% as ‘significant’. We used the DDG2P gene list downloaded 5 February 2025 for annotating genes as ‘well-established’ developmental condition genes.

### Replication analyses in external cohorts

Information about the replication cohorts (ALSPAC, MCS and INTERVAL) is given in the Supplementary Methods. Quality control and imputation of genotype data in ALSPAC and MCS was conducted in Malawsky et al.^94^ and is summarised in the Supplementary Methods, as is the preparation of the exome data which was described in Koko et al.^74^. Quality control in INTERVAL was conducted in Sun et al.^95^ and Astle et al.^96^ and is summarised in the Supplementary Methods.

#### Cognitive performance measures

In ALSPAC, we considered IQ measured at age 8 using the Wechsler Intelligence Scale for Children test^97^. We included 5,283 unrelated children with genetically-inferred European ancestries and at least one genotyped parent in our analyses. In MCS, we derived a cognitive performance measure as previously described in Malawsky et al.^94^ by fitting a one-factor model and calculating scores using *factanal* and Bartlett scoring in R using the following measures: Bracken School Readiness at age 3, reading vocabulary at ages 3 and 5, pattern construction at ages 5 and 7, and word reading and progress in math at age 7. The summarized measure explained 39% of the variance and we included a total of 5,621 unrelated children of genetically-inferred European ancestries with at least one genotyped parent. In INTERVAL, subjects completed a FI test identical in nature to the one completed by UKB participants in the first in-person wave at two time points 12 months apart (test-retest correlation: 0.65, p<10^-15^). We took the average of the fluid intelligence score for individuals who had two measurements. We included 20,328 unrelated individuals of genetically-inferred European ancestries. All variables were standardized to have a mean of 0 and variance of 1.

#### Calculating polygenic indices

We used LDpred2-auto^98^ to calculate PGIs in ALSPAC, MCS, and INTERVAL using our the GWAS for Observed FIS (i.e., measured average FIS), Combined FIS, and Combined FIS+COGENT GWAS meta-analysis. We used unrelated individuals with European ancestries (ascertained as described in the Supplementary Methods) and generated LD reference panels restricting to HapMap3+ SNPs.^88^ We used default parameters to calculate SNP weights for the polygenic indices. We then imputed missing parental genotypes in both cohorts (i.e. if one but not both parents were genotyped) using SNIPAR^36^, as previously described^94^ and calculated PGIs with the SNP weights generated above using the pgs.py script.

#### Estimating direct and population effects of PGIs on cognitive performance measures

To estimate the population effects of the PGIs, we standardized the PGIs as above and regressed the phenotypes on the individual’s PGI, 20 genetic PCs and sex, and additionally age and age^2^ in INTERVAL, using the *lm* function in R. To estimate the direct effects in ALSPAC and MCS, we added the two parental PGIs and additional covariates to the previous regression. The coefficient estimated for the child’s PGI is the partial correlation between the PGI and the phenotype and represents the direct genetic effect.

In MCS, we accounted for ascertainment biases due the cohort’s nonrandom sampling scheme and attrition by incorporating sampling weights as previously described^8^. Briefly, we generated nonresponse weights using inverse probability weighting and multiplied these weights with the full UK sampling weights generated by the study. We then used these weights in the regression analyses conducted in MCS using the weights argument in the *lm* function in R.

#### Multilevel Mixed-Effects Regression

We employed a multilevel mixed-effects regression to evaluate the association between discovery GWAS sample size and standardized PGI effect sizes using the *metafor* package in R^99^. We regressed the PGI betas, weighted by their inverse variance to account for measurement error, on the logarithm of the GWAS discovery sample size.

Random intercepts were included for the cohorts (ALSPAC, MCS, and INTERVAL) and for effect type (population and direct) nested within each cohort to account for heterogeneity in effect estimates across these groups. The model was specified as:

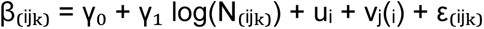

where γ_0_ is the overall intercept, γ_1_ represents the fixed effect of log(N), u_i_ is the random effect for the i-th cohort, v_j_(_i_) is the random effect for effect type nested within the i-th cohort, and ε_ijk_ is the residual error. We also ran the model using direct effects estimates alone, dropping the v_j_(_i_) term.

#### Enrichment of *de novo* damaging variants in NDC probands

We used previously published data from Kaplanis et al.^22^ on three cohorts totaling over 31,000 exome-sequenced probands with developmental conditions and their parents to assess enrichment of damaging *de novo* mutations in the set of 12 genes with rare variant associations with FIS (FDF<1%) that are not DDG2P genes.

We evaluated enrichment of *de novo* synonymous and nonsynonymous variants in the probands as follows. We used the null mutational model^75^ to compute, for each consequence class *i*, the cumulative per-site mutation rate *μ_i,g_* across all callable variants of that class in a given gene *g*. The expected burden of *de novo* mutations of class *i* for a given gene set *G* was then calculated as:

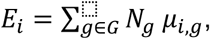

where *N_g_* is the total number of probands. We compared the observed count *O_i_* of d*e novo* mutations in class *i* to *E_i_* by performing a one-sided Poisson test with rate parameter *λ* = *E_i_*, calculating *P*(*X* ≥ *Pois*(*λ*) )_⬚_as the significance of any excess *de novo* mutations. This framework allows assessment of whether damaging protein-truncating or missense variants occur more often than expected by chance, with synonymous variants serving as an internal negative control.

#### Replication of gene-based burden test results in ALSPAC and MCS

To replicate the results of our gene-based tests in UKB, we performed gene-set burden tests in ALSPAC and MCS. For both cohorts, we included only variants with a minor allele frequency (MAF) lower than 0.1% in participants with European ancestries and had a gnomAD V3 ^21^ allele frequency of <3×10^-5^ (corresponding to allele count <5). As in UKB, we only retain high-confidence PTVs defined by LOFTEE^21^, and missense variants with an MPC score greater than 2. We then calculated the burden of PTVs for three gene sets: (i) all 26 genes discovered in UKB at FIS<1%; ii) the FIS<1% genes excluding the eight already identified by Chen et al., and iii) the twenty-one FIS<1% genes that we only discovered when using combined FIS. We used linear regression models to examine the association between cognitive measures and the gene-set burden of PTVs in unrelated children with European ancestries in both the Avon Longitudinal Study of Parents and Children (ALSPAC; N=5,283) and the Millennium Cohort Study (MCS; N=5,621), controlling for sex and population structure with the top 10 principal components (PCs).

## Notes

### Competing Interest Statement

The authors have declared no competing interest.

### Funding Statement

This research was conducted using the UK Biobank Resource under application numbers 40310 and 44165.
This work used the Dutch national e infrastructure with the support of the SURF Cooperative using grant no. EINF 7294.
This research was funded in part by Wellcome (grant no. 220540/Z/20/A, Wellcome Sanger Institute Quinquennial Review 2021 to 2026).
K.J.H.V. is supported by the Foundation Volksbond Rotterdam.
A.A. is supported by the Amsterdam UMC Fellowship.
For the purpose of open access, the authors have applied a CC-BY public copyright license to any author accepted manuscript version arising from this submission.

### Author Declarations

UK Biobank has received ethical approval from the National Health Service North West Centre for Research Ethics Committee (reference: 11/NW/0382) and gave approval for this work. Ethics committee of ALSPAC gave ethical approval for this work Ethics committee of MCS name gave ethical approval for this work

## References

1. Strenze, T. Intelligence and socioeconomic success: A meta-analytic review of longitudinal research. Intelligence 35, 401–426 (2007).

2. Schmidt, F. L. & Hunter, J. E. The validity and utility of selection methods in personnel psychology: Practical and theoretical implications of 85 years of research findings. Psychol. Bull. 124, 262–274 (1998).

3. Keuschnigg, M., van de Rijt, A. & Bol, T. The plateauing of cognitive ability among top earners. Eur. Sociol. Rev. 39, 820–833 (2023).

4. Keyes, K. M., Platt, J., Kaufman, A. S. & McLaughlin, K. A. Association of fluid intelligence and psychiatric disorders in a population-representative sample of US adolescents. JAMA Psychiatry 74, 179–188 (2017).

5. Koenen, K. C. et al. Childhood IQ and adult mental disorders: a test of the cognitive reserve hypothesis. Am. J. Psychiatry 166, 50–57 (2009).

6. Batty, G. D., Deary, I. J. & Gottfredson, L. S. Premorbid (early life) IQ and later mortality risk: systematic review. Ann. Epidemiol. 17, 278–288 (2007).

7. Calvin, C. M. et al. Childhood intelligence in relation to major causes of death in 68 year follow-up: prospective population study. BMJ 357, j2708 (2017).

8. Huang, Q. Q. et al. Examining the role of common variants in rare neurodevelopmental conditions. Nature 636, 404–411 (2024).

9. Polderman, T. J. C. et al. Meta-analysis of the heritability of human traits based on fifty years of twin studies. Nat. Genet. 47, 702–709 (2015).

10. van Leeuwen, M., van den Berg, S. M. & Boomsma, D. I. A twin-family study of general IQ. Learn. Individ. Differ. 18, 76–88 (2008).

11. Briley, D. A. & Tucker-Drob, E. M. Explaining the increasing heritability of cognitive ability across development: a meta-analysis of longitudinal twin and adoption studies: A meta-analysis of longitudinal twin and adoption studies. Psychol. Sci. 24, 1704–1713 (2013).

12. Plomin, R. & Deary, I. J. Genetics and intelligence differences: five special findings. Mol. Psychiatry 20, 98–108 (2015).

13. Markel, G. et al. Nature, nurture, and socioeconomic outcomes: New evidence from sib pairs and molecular genetic data. Social Science Research Network (2025) doi:10.2139/ssrn.5225447.

14. Savage, J. E. et al. Genome-wide association meta-analysis in 269,867 individuals identifies new genetic and functional links to intelligence. Nat. Genet. 50, 912–919 (2018).

15. Trampush, J. W. et al. GWAS meta-analysis reveals novel loci and genetic correlates for general cognitive function: a report from the COGENT consortium. Mol. Psychiatry 22, 336–345 (2017).

16. Sniekers, S. et al. Genome-wide association meta-analysis of 78,308 individuals identifies new loci and genes influencing human intelligence. Nat. Genet. 49, 1107– 1112 (2017).

17. Tan, T. et al. Family-GWAS reveals effects of environment and mating on genetic associations. Genetic and Genomic Medicine (2024).

18. Bulik-Sullivan, B. K. et al. LD Score regression distinguishes confounding from polygenicity in genome-wide association studies. Nat. Genet. 47, 291–295 (2015).

19. Gardner, E. J. et al. Reduced reproductive success is associated with selective constraint on human genes. Nature 603, 858–863 (2022).

20. Chen, C.-Y. et al. The impact of rare protein coding genetic variation on adult cognitive function. Nat. Genet. 55, 927–938 (2023).

21. Karczewski, K. J. et al. The mutational constraint spectrum quantified from variation in 141,456 humans. Nature 581, 434–443 (2020).

22. Kaplanis, J. et al. Evidence for 28 genetic disorders discovered by combining healthcare and research data. Nature 586, 757–762 (2020).

23. Samocha, K. E. et al. Substantial role of rare inherited variation in individuals with developmental disorders. medRxiv (2024) doi:10.1101/2024.08.28.24312746.

24. Young, A. S. & Martin, H. C. Discovering genes that affect cognitive ability. Trends Genet. 39, 810–812 (2023).

25. Cattell, R. B. Theory of fluid and crystallized intelligence: A critical experiment. J. Educ. Psychol. 54, 1–22 (1963).

26. Fawns-Ritchie, C. & Deary, I. J. Reliability and validity of the UK Biobank cognitive tests. PLoS One 15, e0231627 (2020).

27. Davis, K. A. S. et al. Mental health in UK Biobank - development, implementation and results from an online questionnaire completed by 157 366 participants: a reanalysis. BJPsych Open 6, e18 (2020).

28. Stamatakis, E. et al. Is cohort representativeness passé? Poststratified associations of lifestyle risk factors with mortality in the UK biobank. Epidemiology 32, 179–188 (2021).

29. Schoeler, T. et al. Participation bias in the UK Biobank distorts genetic associations and downstream analyses. Nat. Hum. Behav. 7, 1216–1227 (2023).

30. Dahl, A. et al. Phenotype integration improves power and preserves specificity in biobank-based genetic studies of major depressive disorder. Nat. Genet. 55, 2082–2093 (2023).

31. Demange, P. A. et al. Investigating the genetic architecture of noncognitive skills using GWAS-by-subtraction. Nat. Genet. 53, 35–44 (2021).

32. Wood, A. R. et al. Defining the role of common variation in the genomic and biological architecture of adult human height. Nat. Genet. 46, 1173–1186 (2014).

33. Lee, J. J. et al. Gene discovery and polygenic prediction from a genome-wide association study of educational attainment in 1.1 million individuals. Nat. Genet. 50, 1112–1121 (2018).

34. Watanabe, K., Taskesen, E., van Bochoven, A. & Posthuma, D. Functional mapping and annotation of genetic associations with FUMA. Nat. Commun. 8, 1826 (2017).

35. Bulik-Sullivan, B. et al. An atlas of genetic correlations across human diseases and traits. Nat. Genet. 47, 1236–1241 (2015).

36. Young, A. I. et al. Mendelian imputation of parental genotypes improves estimates of direct genetic effects. Nat. Genet. 54, 897–905 (2022).

37. Fraser, A. et al. Cohort Profile: the Avon Longitudinal Study of Parents and Children: ALSPAC mothers cohort. Int. J. Epidemiol. 42, 97–110 (2013).

38. Boyd, A. et al. Cohort Profile: The ‘Children of the 90s’; the index offspring of The Avon Longitudinal Study of Parents and Children (ALSPAC). International Journal of Epidemiology 42, 111–127 (2013).

39. Connelly, R. & Platt, L. Cohort profile: UK Millennium Cohort Study (MCS). Int. J. Epidemiol. 43, 1719–1725 (2014).

40. Di Angelantonio, E. et al. Efficiency and safety of varying the frequency of whole blood donation (INTERVAL): a randomised trial of 45 000 donors. Lancet 390, 2360–2371 (2017).

41. Mbatchou, J. et al. Computationally efficient whole-genome regression for quantitative and binary traits. Nat. Genet. 53, 1097–1103 (2021).

42. Schlingmann, K. P. et al. Germline DE Novo mutations in ATP1A1 cause renal hypomagnesemia, refractory seizures, and intellectual disability. Am. J. Hum. Genet. 103, 808–816 (2018).

43. Yang, L. et al. IGF1R variants in patients with growth impairment: Four novel variants and genotype-phenotype correlations. J. Clin. Endocrinol. Metab. 103, 3939–3944 (2018).

44. Du, W. et al. Calcium-sensitive potassium channelopathy in human epilepsy and paroxysmal movement disorder. Nat. Genet. 37, 733–738 (2005).

45. Tabarki, B., AlMajhad, N., AlHashem, A., Shaheen, R. & Alkuraya, F. S. Homozygous KCNMA1 mutation as a cause of cerebellar atrophy, developmental delay and seizures. Hum. Genet. 135, 1295–1298 (2016).

46. Epi4K Consortium. De Novo mutations in SLC1A2 and CACNA1A are important causes of epileptic encephalopathies. Am. J. Hum. Genet. 99, 287–298 (2016).

47. Helbig, K. L. et al. De Novo pathogenic variants in CACNA1E cause developmental and epileptic encephalopathy with contractures, macrocephaly, and dyskinesias. Am. J. Hum. Genet. 104, 562 (2019).

48. Granadillo, J. L. et al. Pathogenic variants in TNRC6B cause a genetic disorder characterised by developmental delay/intellectual disability and a spectrum of neurobehavioural phenotypes including autism and ADHD. J. Med. Genet. 57, 717–724 (2020).

49. Martin, H. C. et al. Quantifying the contribution of recessive coding variation to developmental disorders. Science 362, 1161–1164 (2018).

50. Chen, G. et al. GIGYF1 disruption associates with autism and impaired IGF-1R signaling. J. Clin. Invest. 132, (2022).

51. Lassuthova, P. et al. Mutations in ATP1A1 cause dominant Charcot-Marie-tooth type 2. Am. J. Hum. Genet. 102, 505–514 (2018).

52. Villamor-Payà, M. et al. De novo TLK1 and MDM1 mutations in a patient with a neurodevelopmental disorder and immunodeficiency. iScience 27, 109984 (2024).

53. Lise, S. et al. Recessive mutations in SPTBN2 implicate β-III spectrin in both cognitive and motor development. PLoS Genet. 8, e1003074 (2012).

54. Elsayed, S. M. et al. Autosomal dominant SCA5 and autosomal recessive infantile SCA are allelic conditions resulting from SPTBN2 mutations. Eur. J. Hum. Genet. 22, 286– 288 (2014).

55. Paul, M. S. et al. A syndromic neurodevelopmental disorder caused by rare variants in PPFIA3. Am. J. Hum. Genet. 111, 1239 (2024).

56. AlAbdi, L. et al. Loss-of-function variants in MYCBP2 cause neurobehavioural phenotypes and corpus callosum defects. Brain 146, 1373–1387 (2023).

57. Iqbal, Z. et al. Homozygous and heterozygous disruptions of ANK3: at the crossroads of neurodevelopmental and psychiatric disorders. Hum. Mol. Genet. 22, 1960–1970 (2013).

58. Kloth, K. et al. First de novo ANK3 nonsense mutation in a boy with intellectual disability, speech impairment and autistic features. Eur. J. Med. Genet. 60, 494–498 (2017).

59. Ferreira, M. A. R. et al. Collaborative genome-wide association analysis supports a role for ANK3 and CACNA1C in bipolar disorder. Nat. Genet. 40, 1056–1058 (2008).

60. Degenhardt, F. et al. Duplications in RB1CC1 are associated with schizophrenia; identification in large European sample sets. Transl. Psychiatry 3, e326 (2013).

61. Iossifov, I. et al. The contribution of de novo coding mutations to autism spectrum disorder. Nature 515, 216–221 (2014).

62. Lim, E. T. et al. Rates, distribution and implications of postzygotic mosaic mutations in autism spectrum disorder. Nat. Neurosci. 20, 1217–1224 (2017).

63. Satterstrom, F. K. et al. Large-Scale Exome Sequencing Study Implicates Both Developmental and Functional Changes in the Neurobiology of Autism. Cell 180, 568– 584.e23 (2020).

64. Vissers, L. E. L. M. et al. Mutations in a new member of the chromodomain gene family cause CHARGE syndrome. Nat. Genet. 36, 955–957 (2004).

65. Sugathan, A. et al. CHD8 regulates neurodevelopmental pathways associated with autism spectrum disorder in neural progenitors. Proc. Natl. Acad. Sci. U. S. A. 111, E4468–77 (2014).

66. Hengel, H. et al. Bi-allelic loss-of-function variants in BCAS3 cause a syndromic neurodevelopmental disorder. Am. J. Hum. Genet. 108, 1069–1082 (2021).

67. A major segment of the neurofibromatosis type 1 gene: cDNA sequence, genomic structure, and point mutations. Cell 62, 609 (1990).

68. Prasad, A. et al. A discovery resource of rare copy number variations in individuals with autism spectrum disorder. G3 (Bethesda) 2, 1665–1685 (2012).

69. St Pourcain, B., et al. Common variation near ROBO2 is associated with expressive vocabulary in infancy. Nat. Commun. 5, 4831 (2014).

70. Butler, K. M. et al. De novo variants in GABRA2 and GABRA5 alter receptor function and contribute to early-onset epilepsy. Brain 141, 2392–2405 (2018).

71. Bochdanovits, Z. et al. Joint reanalysis of 29 correlated SNPs supports the role of PCLO/Piccolo as a causal risk factor for major depressive disorder. Mol. Psychiatry 14, 650–652 (2009).

72. Nievergelt, C. M. et al. Discovery of 95 PTSD loci provides insight into genetic architecture and neurobiology of trauma and stress-related disorders. medRxiv (2023) doi:10.1101/2023.08.31.23294915.

73. Crow, A. J. D. et al. A systematic review and meta-analysis of intellectual, neuropsychological, and psychoeducational functioning in neurofibromatosis type 1. Am. J. Med. Genet. A 188, 2277–2292 (2022).

74. Koko, M. et al. Exome sequencing of UK birth cohorts. Wellcome Open Research (2024).

75. Samocha, K. E. et al. A framework for the interpretation of de novo mutation in human disease. Nat. Genet. 46, 944–950 (2014).

76. Ji, Z., Li, Y., Liu, S. X. & Sharrocks, A. D. The forkhead transcription factor FOXK2 premarks lineage-specific genes in human embryonic stem cells for activation during differentiation. Nucleic Acids Res. 49, 1345–1363 (2021).

77. van der Heide, L. P. et al. FoxK2 is required for cellular proliferation and survival: FoxK2 REGULATES PROLIFERATION AND CELLULAR SURVIVAL. J. Cell. Physiol. 230, 1013–1023 (2015).

78. Kingdom, R., Beaumont, R. N., Wood, A. R., Weedon, M. N. & Wright, C. F. Genetic modifiers of rare variants in monogenic developmental disorder loci. Nat. Genet. 56, 861–868 (2024).

79. Fridman, H., Khazeeva, G., Levy-Lahad, E., Gilissen, C. & Brunner, H. Reproductive and cognitive effects in carriers of recessive pathogenic variants. bioRxiv 2024.09.30.615774 (2024) doi:10.1101/2024.09.30.615774.

80. Heyne, H. O. et al. Mono- and biallelic variant effects on disease at biobank scale. Nature 613, 519–525 (2023).

81. International Mouse Phenotyping Consortium. Mouse gene Dpp8.

82. International Mouse Phenotyping Consortium. Mouse gene Ipo9.

83. Sadakata, T. et al. The secretory granule-associated protein CAPS2 regulates neurotrophin release and cell survival. J. Neurosci. 24, 43–52 (2004).

84. Abdellaoui, A., Dolan, C. V., Verweij, K. J. H. & Nivard, M. G. Gene-environment correlations across geographic regions affect genome-wide association studies. Nat. Genet. 54, 1345–1354 (2022).

85. Mazumder, R., Hastie, T. & Tibshirani, R. Spectral regularization algorithms for learning large incomplete matrices. J. Mach. Learn. Res. 11, 2287–2322 (2010).

86. McCarthy, S. et al. A reference panel of 64,976 haplotypes for genotype imputation. Nat. Genet. 48, 1279–1283 (2016).

87. Willer, C. J., Li, Y. & Abecasis, G. R. METAL: fast and efficient meta-analysis of genomewide association scans. Bioinformatics 26, 2190–2191 (2010).

88. International HapMap 3 Consortium et al. Integrating common and rare genetic variation in diverse human populations. Nature 467, 52–58 (2010).

89. de Leeuw, C. A., Mooij, J. M., Heskes, T. & Posthuma, D. MAGMA: generalized gene-set analysis of GWAS data. PLoS Comput. Biol. 11, e1004219 (2015).

90. Morales, J. et al. A joint NCBI and EMBL-EBI transcript set for clinical genomics and research. Nature 604, 310–315 (2022).

91. Aguet, F., et al. The GTEx Consortium atlas of genetic regulatory effects across human tissues. bioRxiv 787903 (2019) doi:10.1101/787903.

92. Manichaikul, A. et al. Robust relationship inference in genome-wide association studies. Bioinformatics 26, 2867–2873 (2010).

93. Okbay, A. et al. Polygenic prediction of educational attainment within and between families from genome-wide association analyses in 3 million individuals. Nat. Genet. 54, 437–449 (2022).

94. Malawsky, D. S. et al. The differential effects of common and rare genetic variants on cognitive performance across development. medRxiv (2024) doi:10.1101/2024.09.04.24313061.

95. Sun, B. B. et al. Genomic atlas of the human plasma proteome. Nature 558, 73–79 (2018).

96. Astle, W. J. et al. The Allelic landscape of human blood cell trait variation and links to common complex disease. Cell 167, 1415–1429.e19 (2016).

97. Wechsler, D. & Kodama, H. Wechsler intelligence scale for children. 1, (1949).

98. Privé, F., Albiñana, C., Arbel, J., Pasaniuc, B. & Vilhjálmsson, B. J. Inferring disease architecture and predictive ability with LDpred2-auto. Am. J. Hum. Genet. 110, 2042– 2055 (2023).

99. Viechtbauer, W. Conducting Meta-Analyses in R with the metafor Package. J. Stat. Softw. 36, (2010).

