## Supplementary Notes and Figures for "Imputation of fluid intelligence scores reduces ascertainment bias and increases power for analyses of common and rare variants"

### Supplementary Material

#### Index

### General Lay Summary

Research on the genetics of intelligence offers both scientific opportunities and ethical responsibilities. We caully considered the goal of this research, which was, broadly, to better understand the genetics of measures of intelligence in the general population in order to eventually inform our understanding of the causes of clinically recognised intellectual disability and developmental delay. Our research was approved by UK Biobank. In addition, we have prepared this lay summary and FAQ to help broader audiences engage with the motivation for this research and the results.

This study explores how we can improve genetic research on cognitive ability when a large part of the data is missing. Due to its large sample size, most of the statistical power in genetic research on cognitive ability so far has come from UK Biobank. About 40 percent of UK Biobank participants, however, never completed the fluid intelligence test, which limits what previous studies could discover. The missing data also introduces bias, since people who took the test tend to be more educated.

To address this, we estimated the missing fluid intelligence test scores using information from a wide range of related traits, including physical and mental health, behavior, and socio-economic outcomes. We then combined these estimated scores with the actual test scores to conduct the largest genetic analyses of cognitive ability to date.

This approach led to the discovery of many more genetic variants associated with cognition, including rare variants in genes linked to neurodevelopment. These rare variant associations would not have been detectable without the increased sample size provided by imputation, and many are located in genes known to affect brain development. Importantly, the estimated fluid intelligence scores closely matched the genetic patterns of the real test scores and helped reduce bias caused by missing data.

### Frequently Asked Questions (FAQ)

***Why study the genetics of intelligence?***

Firstly, it is important to note that we can only study intelligence through measurable and imperfect proxies, such as specific cognitive tests, and not as some idealized or fixed concept. When we talk about studying the “genetics of intelligence/cognitive ability”, we really mean we are studying the genetics of *specific measures of* intelligence/cognitive ability.

Understanding the genetic and environmental factors that shape cognitive differences can help us better understand certain health conditions, as well as how cognition relates to social and health inequalities. Many of the same genetic variants that influence cognitive ability also affect the risk for neurodevelopmental conditions. Socio-economic outcomes like educational attainment (how long do people spend in education) share roughly half of their genetic influences with intelligence. Because cognitive ability plays such a central role in many modern societies, its relationship with genetics is sometimes controversial and vulnerable to misinterpretation. We believe this is important research that should be carried out carefully, transparently, and ethically.

***What did the study find?***

- We increased the number of people in our genetic analyses from about 270,000 to over 430,000.
- This led to the discovery of more common genetic variants linked to cognitive ability, rising from 385 to 608.
- We detected 26 genes where rare damaging variants are linked to lower cognitive ability, including 14 that were already known to play a role in neurodevelopmental conditions.
- Most people carrying these rare variants in UK Biobank do not have a diagnosis of a neurodevelopmental disorder. This suggests these rare genetic effects can influence cognition even without causing people to reach a clinical threshold for diagnosis.
- The improved data led to more powerful genetic predictors of cognitive ability in other studies.

***What does “imputing intelligence” mean?***

We used a statistical model that learns how a wide range of physical, psychological, and socio-economic outcomes relate to intelligence test scores in people who completed the test. We then used that model to estimate likely scores for individuals who did not take it. This approach is commonly used in science to handle missing data. It helps increase statistical power and reduce bias, especially when participation in the test is not random.

***Are there sensitivities surrounding the imputation of cognitive scores for participants who chose not to complete the test?***

We acknowledge that imputing cognitive scores for missing participants touches on sensitive ground. In UK Biobank, approximately 60% of participants completed the fluid intelligence test at least once, and non-participation arose from various factors, including study design (e.g.

the test was only introduced into the ‘baseline assessment’ part way through recruitment), optional participation, and time constraints. Importantly, participants who formally withdraw their consent are excluded from the dataset, and we regularly updated our data to reflect these exclusions. Imputation is a standard scientific method used to handle missing data and increase statistical power. This approach not only boosts power but also helps reduce biases that arise when analyses include only participants with complete data, who often differ systematically from those with missing data. Other approaches are routinely applied to infer characteristics not directly reported by participants, such as genetic ancestry or environmental exposures. Importantly, our imputed fluid intelligence scores are used solely for research purposes at the group level and are not intended to make predictions about any individual participant.

***Does this study mean intelligence is genetic?***

It was well-established before this study that genes influence cognitive ability, but also that they are only part of the story. The environment, including education, parenting, societal context and health-related factors, also plays a major role. Genetic effects can interact with and correlate with environmental factors. This study helps identify some of the genes that may be involved, but it does not mean that intelligence is fixed or determined at birth.

***How generalizable is this research?***

This study was conducted with individuals of genetically-inferred European ancestries only. That is because most of the available genetic data comes from this group. Results from one group of people are difficult to apply directly to others, not only because of cultural and environmental differences, but also for technical reasons. The patterns of correlation between genetic variants can differ across ancestries, which can affect statistical results even when the underlying genetic effects are similar. This is a serious limitation in many genetic studies, and the field needs to become more inclusive by collecting and analyzing data from more diverse populations.

***Can this research be used to study population differences in measures of intelligence?***

While this question lies well beyond the scope of our study, we address it here because it sits at the heart of some of the public controversy around genetic research on cognitive traits. We are aware that fringe researchers have attempted to (mis)use genetic results and summary statistics in this area, and we believe it is important to clarify why such comparisons are scientifically problematic and should be approached with caution.

Firstly, human ancestries are complex, shaped by overlapping migrations, histories, and admixture. Categories like “European”, “South Asian”, or “Black African” are socially and historically constructed and do not always map cleanly onto underlying genetic patterns. This study only includes individuals of European ancestries and cannot be used to make claims about differences between ancestry groups. Applying results across populations is limited by both social and technical factors.

Even if average genetic scores differ between populations, that does not mean there are true genetic differences in the traits themselves. One reason these scores might differ is due to differences in the pattern of correlations between genetic variants across ancestries; a difference in genetic scores between ancestries does not necessarily imply systematic differences in the frequency of genetic variants that have causal effects on the trait in question (i.e. the difference in genetic scores can exist even if the underlying biology of the trait is the same between ancestries). Furthermore, there is evidence that genetic effects on cognitive traits are sensitive to social and economic conditions, which differ between groups and are shaped by history, inequality, and discrimination. Finally, natural selection can complicate comparisons. For traits that are kept close to an optimal value by natural selection (e.g., through stabilizing selection), different combinations of genetic variants can evolve differently in different populations to maintain the same average trait level. This means that polygenic scores based on one population might suggest differences that are not actually there or miss real differences that do exist.

Because of these issues, genetic comparisons of cognitive traits between populations are both very complex to execute robustly and highly sensitive to misinterpretation.

***Are these findings relevant for clinical or educational use?***

These findings are meant to improve our scientific understanding of cognition, not to make individual predictions. Although it is technically possible to make predictions of measures of intelligence using measures like polygenic scores, we do not use polygenic scores to assess individuals in this paper and do not recommend doing so. Polygenic scores based on this research are noisy and reflect subtle differences that are only useful when studying large groups. They remain inferior to direct measurements of cognition-related traits, such as standardized exam results in school systems. They are neither accurate nor reliable enough for use in personal decision-making in health, education, or employment.

The same caution applies to the rare genetic variants identified in this study. While some of the genes we found that show rare variant associations with fluid intelligence are known to play a role in neurodevelopmental conditions, many people who carry rare variants in these genes do not have a clinical diagnosis. Our findings highlight potential difficulties in interpreting rare variants in these genes found in patients with a rare neurodevelopmental condition, particularly if those variants have been inherited from clinically unaffected family members. This problem of interpreting variants like this with “incomplete penetrance” is likely to become an increasing challenge for clinical genetics over the coming years.

***Can this research be used for embryo selection?***

This research was not intended for embryo selection, and applying it in that context raises important scientific and ethical concerns. Polygenic scores have limited predictive power within families (i.e. the scenario in which they would be used when selecting one of a set of embryos produced from the same couple), especially for traits like intelligence. More importantly, these scores reflect a mix of biological and social influences, many of which are shaped by the current environment and may change over time.

Many countries, including the UK, have strict regulations on the use of polygenic scores in embryo selection. However, some countries allow their use under certain circumstances. We do not recommend using polygenic scores of measures of intelligence in any such contexts. There are many reasons for this, including technical challenges in applying selection, the limited variance explained even under theoretically optimal conditions, the small number of embryos available, and uncertainties about what the scores actually capture.

Selecting embryos based on polygenic scores also involves trade-offs that are hard to foresee. For example, the genetic variants that are associated with higher intelligence also tend to increase the risk for autism, meaning that selecting for one trait could unintentionally affect another. Because complex traits are influenced by many overlapping biological pathways, the long-term effects of selecting embryos in this way are unpredictable.

***What is the next step?***

We hope other researchers will use our methods to improve the quality of genetic research in large datasets. We also intend to use our results to see if they help us discover more genes associated with neurodevelopmental conditions in clinical cohorts. Future studies should apply these approaches in more diverse populations, and continue to explore how genes and environments interact to shape cognitive outcomes and neurodevelopmental conditions.

### Supplementary Glossary

#### Supplementary Box: Glossary of Abbreviations and Terms

| Abbreviation | Main text abbreviation | Description |
|---|---|---|
| **FIS** | | Fluid intelligence score |
| ***Observed FIS1** | FIS1 | UKB baseline measured FIS (20016-0.0) |
| ***Earliest observed FIS** | Earliest FIS | Earliest measured FIS across 5 measured tests |
| ***Average observed FIS** | Average FIS/Observed FIS | Average measured FIS across 5 measured tests |
| ****ImpFIS-vA** | | Imputed FIS only, from imputation vA |
| ****ImpFIS-vB** | | Imputed FIS only, from imputation vB |
| ****ImpFIS-average** | Imputed FIS | Imputed FIS only, from imputation using the average approach |
| ****ImpFIS-earliest** | | Imputed FIS only, from imputation using the earliest approach |
| ****ImpFIS-all5** | | Imputed FIS only, from imputation using the all5 approach |
| ***MegaFIS-vA** | | Mega analysis of Observed FIS1 and ImpFIS-vA |
| ***MegaFIS-vB** | | Mega analysis of Observed FIS1 and ImpFIS-vB |
| ***MegaFIS-average** | Combined FIS | Mega analysis of Average observed FIS and ImpFIS-average |
| ***MegaFIS-earliest** | | Mega analysis of Earliest observed FIS and ImpFIS-earliest |
| ***MegaFIS-all5** | | Mega analysis of Observed FIS1 and ImpFIS-all5 |
| **COGENT** | COGENT | GWAS meta-analysis of intelligence across 24 cohorts in the COGENT consortium by Trampush et al.[1] No UKB participants included. |
| **Savage et al. Intelligence** | Savage et al. Intelligence | GWAS meta-analysis of intelligence in 14 cohorts, including UKB, by Savage et al.[2] |
| **NonCogIFIS** | | GWAS-by-subtraction derived GWAS of Intelligence (Savage et al.) minus ImpFIS-vA |

See *Supplementary Figure 4 and **Supplementary Table 5 for sample sizes

### Supplementary Notes

#### Supplementary Note 1: Available Fluid Intelligence data in UKB

Currently, the UK Biobank has six measurements of Fluid Intelligence Scores (FIS) (see Supplementary Table 1 and Supplementary Figure 1). Four of these measurements were taken in person during different types of visits, and the other two were taken via an online questionnaire. The in-person and online measures are on different scales; in the in-person measurements, thirteen questions had to be answered within two minutes, whereas in the online version, fourteen questions had to be answered in the same amount of time. The order of questions was the same across in-person and online measures, hence the fourteenth measure in the online questionnaire is additional. Only nine people attempted this additional question and answered it correctly within the two-minute time constraint. Thus, we decided to set FI scores of fourteen to thirteen to broadly align scales between measures, retaining these people in the top performing percentile.

We calculated the test-retest correlations among individuals with multiple FIS measurements (Supplementary Table 2). The test-retest correlations were fairly modest across all FIS measurements, in the order of r=~0.6, as previously reported[3]; thus, they are less reliable than standard IQ tests which have test-retest correlations closer to 0.9[4]. To verify whether the different FIS measurements capture similar genetic signals, we ran GWASs for each measure and computed genetic correlations between them (Supplementary Table 2). None of them had a genetic correlation with any other that was significantly lower than 1, except FIS5 which has a genetic correlation with FIS1 and FIS3 slightly below 1.

We noted that the mean and variance were largely consistent across FIS measures, with the exception of FIS5 having a slightly lower mean (Supplementary Figure 2A). This slightly lower mean is likely to be due to several factors, including participants experiencing cognitive decline as time went on as well as ascertainment biases in the characteristics of people taking the different tests. Notably, we observed that having completed the various fluid intelligence tests was associated with higher educational attainment, particularly for FIS2 to FIS5 (Supplementary Figure 2B). It has been shown that non-random ascertainment can influence heritability and genetic correlation estimates. Thus, we decided to impute fluid intelligence scores for UK Biobank participants for whom they were not measured, with two goals in mind: i) to increase power for genetic studies of fluid intelligence, and ii) to ameliorate the ascertainment bias whereby more educated individuals were more likely to complete the tests.

#### Supplementary Note 2: Comparison of imputation strategies

We went through three iterations of imputation strategies. In the first iteration of imputed FIS (ImpFIS-vA), we sought to impute individuals without a FIS1 measurement using 154 correlated variables that had lower missingness (e.g., educational attainment, time spent on a computer, and household income) (Supplementary Table 4). The accuracy of imputation was assessed based on the correlation between observed and imputed FI scores for a set of randomly chosen participants (Supplementary Table 5), whose measured values were set to missing prior to imputation. This imputation achieved an accuracy of r=0.52 (SE = .006) (Supplementary Figure 4A, Supplementary Table 5), which is slightly lower than the test-retest correlation of FI we found in UK Biobank (r ≈ 0.6). Measured FIS1 (FIS1, N=146,526) and imputed scores (ImpFIS-vA, N=309,128) were then standardized separately and mega-analysed in a single GWAS (MegaFIS-vA, N=455,654), which we found performed better than meta-analyzing separate GWASs of measured and imputed FIS1 (Supplement 3; Supplementary Figure 6; Supplementary Table 6). MegaFIS-vA had an estimated $h^2_{pop}$ of 0.19 (SE = .005), which was similar to that of the measured phenotypes (Supplementary Figure 4C) though significantly lower than that for FIS1 ($p = 3.57x10^{-4}$, z-test). Subsequently, we computed genetic correlations between 73 traits and MegaFIS-vA and compared the genetic correlation profile to that derived by correlating the COGENT GWAS with those same 73 traits (Supplementary Figure 3). Specifically, we noted that MegaFIS-vA showed higher genetic correlations with socioeconomic-related variables such as NonCogEA (~0.36 versus ~0.2; p=) and height (~0.23 versus ~0.05) (Supplementary Figure 4D). The within-family GWAS on ImpFIS-A showed that the $h^2_{direct}$ was about a third lower than the heritability obtained in the population-based GWAS (Supplementary Figure 4C). We hypothesized that these results may be due to some of the selected imputation variables only correlating with FI via confounding paths such as population stratification.

In the next iteration we used GWAS-by-subtraction[5] (Supplementary Figure 7) to subtract the signal of the Savage et al. Intelligence GWAS. from our previous GWAS on imputed values (ImpFIS-vA, N=309,128), deriving a GWAS of “non-cognitive imputed FIS” (NonCog-iFIS).

The NonCog-iFIS GWAS captured signals associated with imputed FI that are not observed in the Savage et al. Intelligence GWAS, and thus likely reflected confounding due to correlations between measured FI and e.g. socioeconomic variables. We then calculated genetic correlations between the 154 imputation variables used to produce ImpFIS-vA and both NonCog-iFIS and the Savage et al. intelligence GWAS (Supplementary Figure 8, Supplementary Table 4). Based on these results, we refined the set of imputation variables to a subset of 82 variables that showed a stronger genetic correlation with intelligence than with NonCog-iFIS. These variables were then used for a new imputation of FIS1 (ImpFIS-vB), achieving an accuracy of r = 0.49 (SE=.006), which was comparable to (albeit slightly lower) than that obtained in ImpFIS-vA (Supplementary Table 5, Supplementary Figure 4A). We again combined ImpFIS-vB with measured FI scores (FIS1) in a GWAS via mega-analysis (MegaFIS-vB). The resulting GWAS had an estimated $h^2_{pop}$ = 0.18 (SE=.005) (Supplementary Figure 4C), similar to that obtained with MegaFIS-vA. Additionally, MegaFIS-vB had a similar pattern of $r_g$ with external trait GWASs as MegaFIS-vA , except that it had a substantially lower $r_g$ with height (Supplementary Figure 4D). However, the elevated genetic correlation with NonCogEA remained ($r_g$=0.36, SE=.02) and after conducting a within-family GWAS of MegaFIS-vB, we estimated $h^2_{direct}$ to be about half that of population-based GWAS (Supplementary Figure 4C), suggesting the imputed phenotype still contained substantial confounding signal.

In the final iteration, we tried three different strategies to further refine the imputation. In the first two, we input either the average or the earliest FIS measures into the imputation as single variables alongside the 82 variables used above. Imputed missing values (ImpFIS-average, ImpFIS-earliest) are then taken for downstream analysis. In the third, we put all five FIS measures into the imputation as separate variables alongside the aforementioned 82 variables, imputed missing values, then used the imputed values for FIS1 in downstream analyses ("ImpFIS-all5") (Supplementary Figure 5). These three strategies produced accuracies of ~0.45-0.55, similar to the first two imputation attempts (Supplementary Figure 4A), with ImpFIS-all5 being the highest. It is important to note, however, that the overall accuracy of ImpFIS-all5 is inflated because some of the individuals who were imputed for FIS1 had other FIS measures, and can thus be imputed more accurately (Supplement 4, Supplementary Table 5). In all approaches, the highest accuracy was observed for individuals in the top and bottom terciles of the measured phenotype distribution (Supplementary Figure 9).

We then conducted a GWAS mega-analysis of each of these with average observed FIS (MegaFIS-average), earliest observed FIS (MegaFIS-earliest) or observed FIS1 (MegaFIS-all5) measures respectively. Imputation increased the sample size to around 455,640 individuals with estimated $h^2_{pop}$ values of 0.16 (SE = .005) for both MegaFIS-average and MegaFIS-earliest, and 0.18 (SE = .005) for MegaFIS-all5 (Supplementary Figure 4D). We also conducted within-family GWAS mega-analyses on the three phenotypes and found minimal attenuation of point estimates of $h^2_{direct}$ relative to $h^2_{pop}$ (Supplementary Figure 4C), consistent with the results for the measured phenotypes. For all three of these imputation approaches approaches, the genetic correlation with NonCogEA attenuated to values consistent with the $r_g$ estimated using the COGENT cognition GWAS[1,2] ($r_g \approx$ 0.20, SE=.02) (Supplementary Figure 4D), and the $r_g$ with height remained in line with that using measured FIS1 ($r_g$=0.11, SE = .02). Lastly, we found that MegaFIS-average and MegaFIS-earliest yielded $r_{direct\text{-}NTC}$ not significantly different from 0, though MegaFIS-all5 did have a significant negative $r_{direct\text{-}NTC}$ (Supplementary

Figure 4G). Collectively, these results suggested that the MegaFIS-average and MegaFIS-earliest imputation approaches yielded traits that were highly genetically similar to the measured FIS while reducing ascertainment biases. We observed a slightly higher phenotypic imputation accuracy, more significant lead SNPs, and slightly higher $h^2_{pop}$ for MegaFIS-average than for MegaFIS-earliest, so decided to use that for the main downstream analyses.

#### Supplementary Note 3: Combining measured and imputed data

The goal of imputation was to combine the measured and imputed values to obtain a single GWAS with the largest effective sample size possible. We examined whether combining them should be done by meta- or mega-analysis by applying both strategies to the imputed and measured values from the last three imputation approaches i.e. ImpFIS-earliest, ImpFIS-average and ImpFIS-all5. For the mega-analyses, we scaled imputed and measured values separately before combining them and then ran a GWAS as described in the Methods. In the meta-analysis we used GWASs conducted on imputed and measured values separately and then meta-analyzed them using METAL[6] following the procedure described in the Methods. Ultimately, we ended up with six GWASs: a meta- and mega-analysis derived-GWAS for each of the three imputation approaches (ImpFIS-average, ImpFIS-earliest and ImpFIS-all5).

Generally, scaling measured and imputed values separately and combining them by mega-analysis showed better results than meta-analysis across all approaches, namely lower genetic correlations with NonCogEA and higher genetic correlations with the measured intelligence GWASs (Supplementary Figure 6). In addition, the mega-analyses contain more genetic signals, as demonstrated by higher heritability estimates (Supplementary Table 6). When combined with the observed measures via mega-analysis, the three imputation strategies performed extremely similarly in terms of their genetic correlations with other traits.

We acknowledge that this mega-analysis approach of scaling the measured and imputed values separately and then combining them is flawed in that it is unlikely that these two groups truly have the same mean FI, given that they differ in average educational attainment (Supplementary Figure 2). The imputed FI scores show considerably less variance than the measured scores, as a consequence of the imputation tending to push people towards the middle of the distribution. In theory, the meta-analysis approach gets around these differences in mean and variance between the observed and imputed values, but since in practice, we found that the mega-analysis worked slightly better, we decided to use this for downstream analyses.

#### Supplementary Note 4: Performance of the ImpFIS-all5 strategy in different subsets

Combining measurements before imputation in ImpFIS-average and ImpFIS-earliest causes a discrepancy in what is imputed between these approaches and ImpFIS-all5. In ImpFIS-average and ImpFIS-earliest, only individuals without any FIS will be imputed, whereas in the ImpFIS-all5, all individuals without a FIS1 score are imputed, including those that are scored in FIS2-5. To be able to directly compare imputed FIS across approaches, we partitioned ImpFIS-all5 individuals into two groups (Supplementary Figure 5): (i) people who *do* have a score available, but not at FIS1 (ImpFIS-all5 ever-measured); and (ii) people *without any* available score (ImpFIS-all5 never-measured). This allows the ImpFIS-all5 never-measured group to be compared fairly with ImpFIS-average and ImpFIS-earliest.

We compared these subsets based on imputation accuracies, estimated SNP $h^2$, and $r_g$ with NonCogEA and Savage et al. Intelligence (Supplementary Table 5 & 7). Although ImpFIS-all5 appeared to perform better overall than the ImpFIS-average and ImpFIS-earliest (most notably having higher imputation accuracy, lower $r_g$ with NonCogEA and higher $r_g$ with Intelligence), we saw that distinguishing between ever-measured and never-measured individuals attenuates this advantage. While ImpFIS-all5 ever-measured shows $r_g$ and estimated SNP $h^2$ values similar to measured value GWASs, ImpFIS-all5 never-measured shows $r_g$ and estimated SNP $h^2$ values in line with those of ImpFIS-average and ImpFIS-earliest. These findings indicated that all imputation approaches work similarly when evaluated within the same set of individuals (i.e. those with no FIS measure), which allowed for appropriate comparisons of corresponding mega-analyses. Consequently, we decided to proceed with MegaFIS-average in downstream analysis.

### Supplementary Note 5: Rare variant analyses

#### Exome-wide burden analyses

The burden of rare (MAF<0.001%) PTVs and damaging missense variants was negatively associated with average observed FIS and with ImpFIS-average (Supplementary Figure 10). As expected, rare PTVs and damaging missense variants in constrained genes (pLI>0.9) showed stronger associations than all such variants exome-wide (Supplementary Figure 10). However, the exome-wide burden of rare synonymous variants, which we expect to show no association, was significantly positively associated with both ImpFIS-average and average observed FIS, with a significantly larger effect size for ImpFIS-average (Supplementary Figure 10). This is likely due to fine-scale population structure that is not controlled for by the inclusion of 25 common variant-based principal components; this fine-scale structure is likely correlated with variables used in the imputation and hence amplified when using imputed rather than observed FIS. We found that additionally controlling for the number of singleton synonymous variants exome-wide (as proxy for this fine-scale structure) rendered the association between observed average FIS and rare synonymous burden non-significant; it also reduced the association with ImpFIS-average, while barely altering the negative association with rare PTV and damaging missense variants (Supplementary Figure 11), indicating that the latter is likely to be causal.

#### Sensitivity analyses controlling for the number of singleton synonymous variants

Given the concerns about subtle population stratification highlighted above in the analyses of exome-wide rare variant burden, we repeated the gene-based rare variant tests of average observed FIS and MegaFIS-average while controlling for the number of singleton synonymous per individual. We found this made very little difference to the significance of our associations (Supplementary Figure 13), suggesting that our gene discovery analyses are robust to any subtle stratification. This is presumably because we are only powered to find large effects which are most likely to be truly causal.

##### Comparison of our results with those of Chen et al.

At our FDR<1% threshold, we replicate all of the genes reported by Chen et al. as associated with FI (which they call 'verbal-numerical reasoning', VNR) at Bonferroni significance (*KDM5B*, *ANKRD12*, *RC3H2, CACNA1A*); of these, all passed this same p-value threshold ($p<4.2 \times 10^{-6}$) in our analysis using the PTV burden test on observed FIS - the closest comparison with Chen et al. - except for *CACNA1A* ($p=1.8 \times 10^{-3}$ on this test, although much more significant when considering combined FIS ($p=6.4 \times 10^{-6}$ on the PTV burden test; Supplementary Table 12). The additional five genes they reported at FDR<5% did not pass our FDR<1% threshold (*NDUFA6*, *ARHGEF7*, *C11orf94*, *KIF26A* and *MAP1A*).

Differences between our results and those of Chen et al. can likely be explained by several factors. They integrated the various FI measures in a different way to us: specifically, they considered the initial assessment visit, first repeat assessment visit and first imaging visit (FIS1, 2 and 3 in Supplementary Table 1) and took the highest measure of these. They also only considered unrelated individuals. Hence, their sample size for VNR was only N=128,812 unrelated Europeans, whereas ours was N=273,091 for observed FIS. Furthermore, they considered only burden tests on PTVs, whereas we also conducted tests incorporating missense variants, as well as SKATO tests. Finally, they used slightly different variant and genotype QC to us, and also presumably calculated their minor allele frequency for filtering on a different set of individuals. However, the most important difference between our studies is our incorporation of imputed FI; this explains most of the increase in the number of significant genes.

#### Supplementary Note 6: Novel genes showing rare variant associations with FIS

Here we discuss the eight genes passing FDR<1% in the rare variant association tests with FI which are neither DDG2P genes nor already reported by Chen *et al.*[7] (Table 1). Both mono- and biallelic disruption of *ANK3* has been reported to cause non-syndromic ID, with the biallelic form being apparently more severe[8]. Recurrent deletions in *ROBO2*, encoding an axon-binding receptor, have been reported in autism[9], and this gene is also the most likely causal gene near a GWAS hit for expressive vocabulary in infancy[10] which is associated with later cognitive ability[11]. *TLK1* encodes a serine/threonine kinase in the same family as *TLK2*, a gene known to cause a neurodevelopmental condition (NDC)[12]; *de novo* missense mutations in *TLK1* have been reported in two NDC patients[13]. *CHD9* is in the same family of chromatin remodelers as *CHD7* and *CHD8*, both well-known NDC-associated genes[14,15]; all three of these genes are in gene modules that are enriched for expression in the prenatal human brain[16]. Some *de novo* missense mutations in *CHD9* have been reported in autism[17–19]. Duplications in *RB1CC1* are associated with schizophrenia[20]. *PCLO* encodes a protein involved in synaptic transmission and is a GWAS hit for major depression[21] and post-traumatic stress disorder[22]. There is no prior genetic evidence directly linking *DPP8* and *IPO9* to neurodevelopment in humans, although *Dpp8*-deficient mice showed some behavioural abnormalities[23] and brain development was severely disrupted in *Ipo9*-deficient mice[24].

### Supplementary Note 7: Association between *CADPS2* and educational attainment

We found a significant positive association between rare PTVs in *CADPS2* and educational attainment (burden test beta = 0.31, p value = $2.5 \times 10^{-6}$), but found no effect on FIS ($p>0.05$ on all tests). This finding is consistent with results attained by Chen et al., which also showed a suggestive positive association between PTVs in *CADPS2* and EA (beta = 0.25, p value = $4.9 \times 10^{-4}$). The absence of impact on FIS provides molecular evidence that EA is genetically distinct from FIS, suggesting *CADPS2* may influence traits specifically related to educational persistence or achievement rather than cognitive ability per se. Consistent with these rare variant findings, *CADPS2* has been implicated in a MAGMA gene prioritization analysis of EA using a common variants GWAS[25] (FDR-adjusted p-value=$7.55 \times 10^{-14}$), but we find no evidence for gene prioritization using MAGMA using any of our FIS GWASs (FDR adjusted p-value>0.05). Additionally, a conditional and joint (COJO) multi-SNP analysis of the largest GWAS of EA[26] identified two lead SNPs at this locus (rs6466819 and rs2430048, COJO p-value < $10^{-11}$ for both), both of which had $p > 0.01$ in our FIS GWASs as well as $p > 0.6$ in the GWAS for the cognitive component of EA[5]. Interestingly, rs6466819 showed a suggestive association ($p=1.5 \times 10^{-4}$) with the non-cognitive component of EA [5], and other SNPs at this locus have been associated with risk taking[27] (rs10252114; $p=5 \times 10^{-11}$) as well as with brain imaging traits[28] .

The rs6466819 GWAS hit for EA has been colocalized with an eQTL for *CADPS2* in cerebellum by OpenTargets[29]. This analysis found that variants associated with decreased expression of *CADPS2* were associated with increased EA. For example, the A allele at the lead eQTL at the locus, rs10241928, is associated with decreased *CADPS2* expression (eQTL beta = -0.28, p value = $1.7 \times 10^{-6}$) and with increased EA in Okbay et al. (GWAS beta = 0.00904, p value = $3.1 \times 10^{-7}$). The same conclusion was reached when using either the Okbay et al. or Lee et al. GWAS results. Coupled with our observation, this suggests we have identified a directionally concordant allelic series at *CADPS2* for EA.

*CADPS2* is strongly expressed in various brain regions[30,31] has been implicated in autistic-like behavioural phenotypes in mice with heterozygous loss-of-function mutations [32]. Additionally, the gene's LOEUF score of 0.7 (gnomAD v4.1.0) indicates mild depletion of heterozygous pLoF carriers, consistent with subtle but not strong constraint against loss-of-function variation.

In summary, both rare and common variant evidence points at a potential role for *CADPS2* in EA via traits other than cognition. The rare variant association first needs to be confirmed in an independent dataset. If confirmed, it is notable for being the first gene (to our knowledge) in which rare loss-of-function variants are associated with increased rather than decreased EA.

### Supplementary Tables

#### Supplementary Table 1

Overview of available FIS measures in UKB. Datafield 20016 is taken at the assessment centre via touch screen whereas 20191 is an online web-based questionaire. We excluded the data collected at the first repeat imaging visit from our analyses due to the small sample size.

#### Supplementary Table 2

In both tables, the diagonal shows the correlation between raw values and transformed values (in which FIS was regressed on age and age2 then the intercept was added to the residual (see Methods) for each measure. Correlations for raw FIS measures are shown above the diagonal, and below the diagonal are the correlations for transformed FIS values. Standard errors are given in brackets. Note that the phenotypic correlations are effectively the test-retest reliability, and are based on the subset of individuals who did both of the relevant tests (shown in Supplementary Figure 1). The genetic correlations are based on GWASs of each respective measure and therefore contain only partially overlapping sets of individuals.

#### Supplementary Table 3

Table showing LDSC calculated genetic correlations for Observed FIS1; Integrated GWASs Earliest FIS, Average FIS and all5 FIS; Mega analyzed GWASs MegaFIS-vA, MegaFIS-vB, MegaFIS-earliest, MegaFIS-average, MegaFIS-all5; and intelligence reference GWASs by Savage et al. and the COGENT consortium.

#### Supplementary Table 4

Overview of UKB variables used for imputation in this study. Per variable the phenotypic correlation with FIS1 is shown, along with the genetic correlation with NonCogIFIS and Savage et al. Intelligence. The final selection of imputation variables covers a subset of all variables listed. Inclusion is shown in the last column.

#### Supplementary Table 5

Accuracy for the different imputation runs, and number of individuals used to assess this. Accuracy is computed as correlation between observed and imputed values for N individuals set to missing prior to imputation. Note that N differs per imputation run due to differing numbers of individuals amongst our static evaluation set of 50,000 randomly-chosen individuals having a measured value.

#### Supplementary Table 6

Common variant based comparison of combining imputed and observed measures. The top shows statistics of combining GWAS summary statistics via meta-analysis using METAL. The bottom shows statistics of combining via mega-analysis after first scaling imputed and measured values separately. Mega-analysis shows both higher SNP heritability and more identified lead SNPs using FUMA. Note that the sample sizes differ slightly between the various imputation runs due to how the outlier removal was done (Methods).

#### Supplementary Table 7

Here we compare downstream GWAS analyses of imputed and measured individuals separately within each imputation approach. ImpFIS-average and ImpFIS-earliest perform very similarly across all metrics. Although at first glance ImpFIS-all5 appears to achieve the best imputed set, splitting this up (Supplementary Figure 5) shows the results are inflated by measured individuals

#### Supplementary Table 8

MAGMA results produced by the SNP2GENE tool in FUMA. The type column indicates the GWAS used as input. Per GWAS only FDR<1% significant protein-coding genes are shown.

#### Supplementary Table 9

Statistics of PGIs constructed with GWAS summary statistics from the various GWAS, tested in three independent cohorts. In ALSPAC we used full-scale IQ measured at age 8. In MCS we used a cognitive performance measure derived from a factor analysis using a combination of various cognitive tests administered across childhood. In INTERVAL we used the average of the two fluid intelligence tests administered 24 and 48 months after recruitment. The population effects indicate results from a regression of the PGIs on the cognitive ability measure, controlling for sex and 20 PCs. The direct effects were estimated in the two birth cohorts by additionally controlling for parental PGIs in the regressions.

#### Supplementary Table 10

Results from MAGMA gene-level tissue enrichment implemented in FUMA SNP2GENE. Per Intelligence measure -log10(P) values are shown for each GTEx v8 Tissue.

#### Supplementary Table 11

Regenie results for observed FIS (i.e. average measured FIS) and combined FIS (i.e. megaFIS-average), for all tested genes. The column headers indicate the phenotype tested in the first row, then the variant consequence included in the second row (PTV, damaging missense (MIS) or PTV+MISS). N_carrier: number of rare variant carriers; BETA, SE, p are the effect size, standard error and p-value from the burden test; P_SKATO is the p-value from the SKATO test.

#### Supplementary Table 12

Regenie results for observed FIS (i.e. average measured FIS) and combined FIS (i.e. megaFIS-average), for genes passing FDR<1%. (This is a subset of Supplementary Table 11, provided for convenience). The column headers indicate the phenotype tested in the first row, then the variant consequence included in the second row (PTV, damaging missense (MIS) or PTV+MISS). N_carrier: number of rare variant carriers; BETA, SE, p are the effect size, standard error and p-value from the burden test; P_SKATO is the p-value from the SKATO test.

#### Supplementary Table 13

Regenie results for observed FIS (i.e. average measured FIS) and combined FIS (i.e. megaFIS-average), for genes passing FDR<1% after excluding individuals with ICD10 codes for intellectual disability, epilepsy or autism. The column headers indicate the phenotype tested in the first row, then the variant consequence included in the second row (PTV, damaging missense (MIS) or PTV+MISS). N_carrier: number of rare variant carriers; BETA, SE, p are the effect size, standard error and p-value from the burden test; P_SKATO is the p-value from the SKATO test.

#### Supplementary Table 14

Regenie results for educational attainment. The column headers in row 4 indicate the variant consequence included (PTV, damaging missense (MIS) or PTV+MISS). BETA, SE, p are the effect size, standard error and p-value from the burden test; P_SKATO is the p-value from the SKATO test.

### Supplementary Figures

#### Supplementary Figure 1

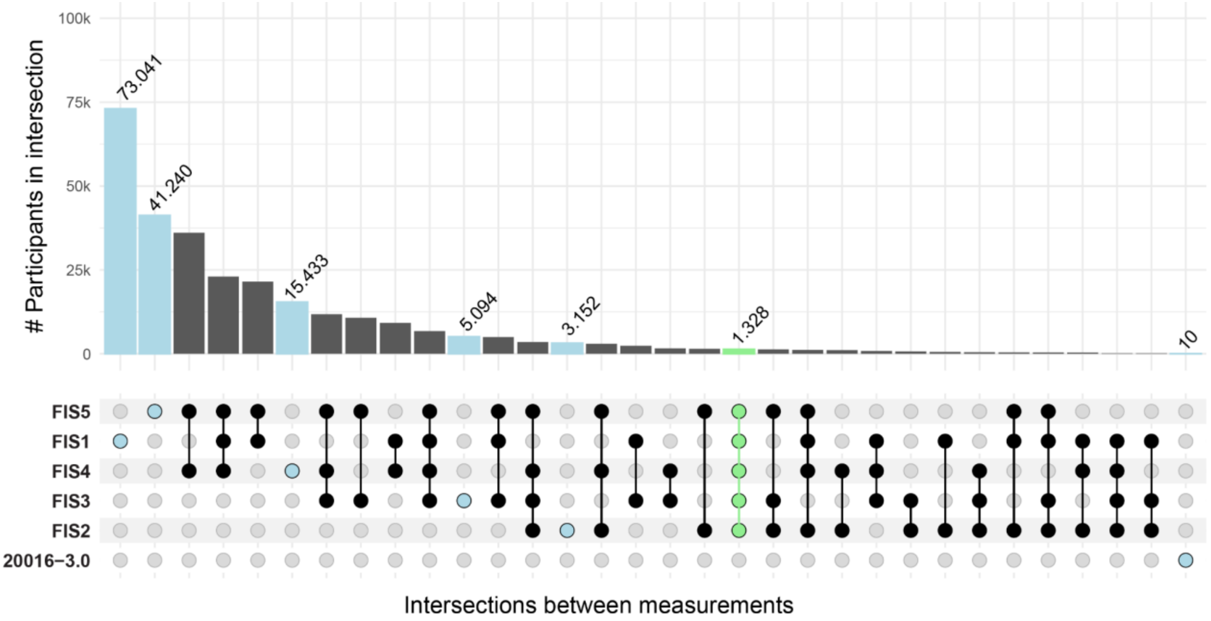

***Intersections between available FIS measurements*.** Highlighted in blue: Set of participants who only have the indicated measure; Highlighted in green: Intersection of participants measured at all FIS measures. Most participants have a FIS measure either at 20016-0.0 (FIS1) or one of the online FIS measures 20191-0.0 (FIS4), 20191-1.0 (FIS5), or a combination of those. 10 people were measured only during measurement 20016-3.0, which is why we omitted this measure in downstream analyses.

### Supplementary Figure 2

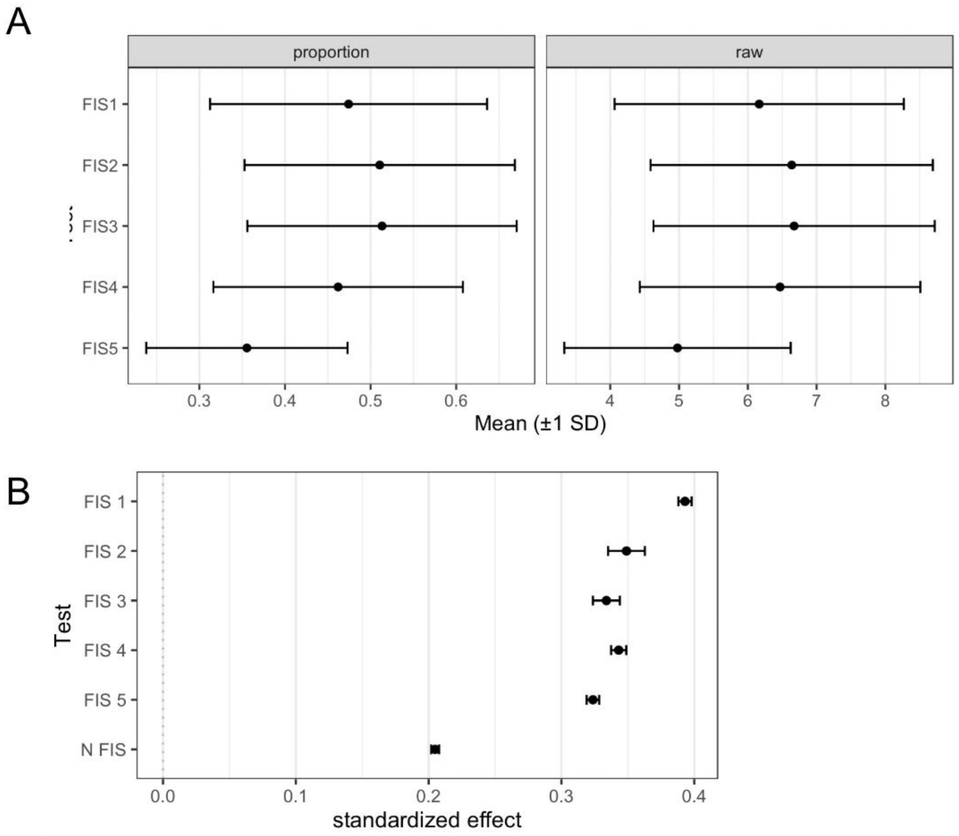

***Average scores on the various FIS tests and association between FI test completion and number of FIS measures taken with EA.*** **A)** Mean and 1 standard deviation bands of FIS measures divided by total number of questions asked (proportion) and of the unscaled scores (raw). **B)** Associations between educational attainment (EA) and having a measured value for a given FIS measure and the total number of FI measures recorded using logistic and linear regressions, respectively.

### Supplementary Figure 3

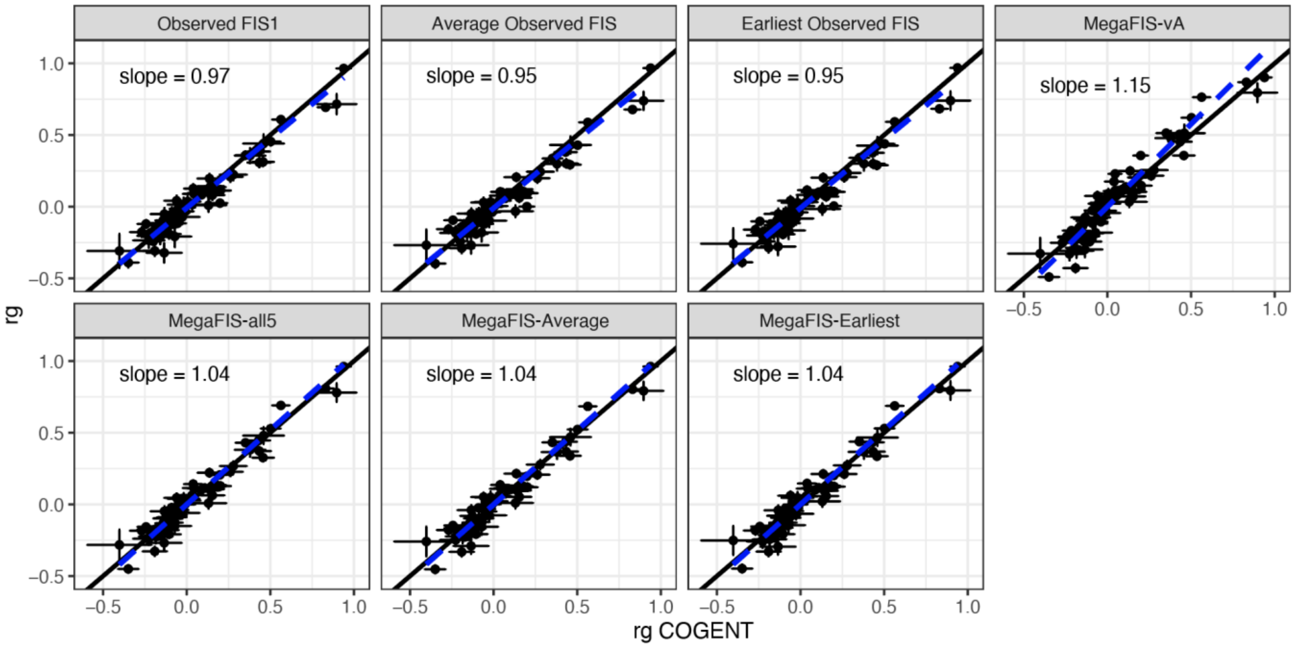

***Genetic correlation profile comparison between derived FIS GWAS and COGENT.*** Genetic correlations are estimated between derived FIS GWASs and 68 UKB phenotypes and plotted against genetic correlations estimated between COGENT and the same 68 UKB phenotypes. The black line indicates the y=x line and the dashed blue line indicates the Deming regression line of best fit, which takes into account the fact that genetic correlations are estimated with error.

### Supplementary Figure 4

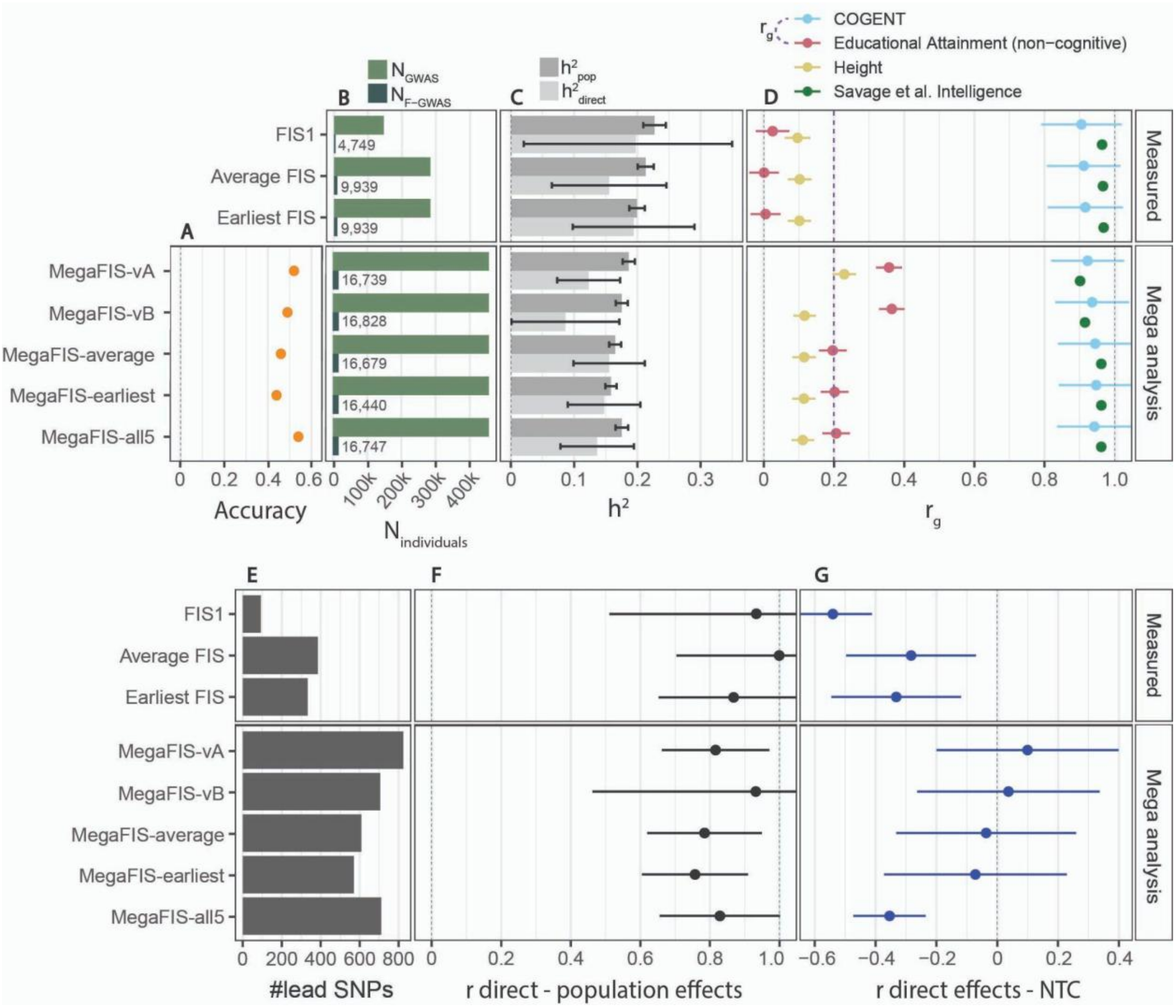

***Extended version of Figure 1, adding statistics on MegaFIS-earliest and MegaFIS-all5.*** All panels give various summary statistics from either population-based or within-family GWASs of measured FI phenotypes (top three results) and our via mega-analyses combined GWAS (bottom result). **A)** Sample sizes of population GWASs ($N_{GWAS}$) and within-family GWASs ($N_{F\text{-}GWAS}$); **B)** Population and direct effect heritabilities estimated using LDSC[33]; **C)** Population effect genetic correlations estimated using LDSC[34] with traits from external studies of intelligence (blue and green), the non-cognitive component of educational attainment (NonCogEA) (red), and height (yellow). The purple dashed line indicates the genetic correlation between the COGENT GWAS and NonCogEA; **D)** Number of independent lead SNPs from the population-based GWASs computed using FUMA[35]; reaching genome-wide significance at $p<5x10^{-8}$. **E)** Genetic correlations estimated using LDSC between the population effects and the direct effects from a within-family GWAS conducted on the corresponding trait; **F)** Genome-wide correlation between direct effects and non-transmitted coefficients (NTC) estimated using snipar[36] for each corresponding trait. Error bars indicate 95% confidence interval.

### Supplementary Figure 5

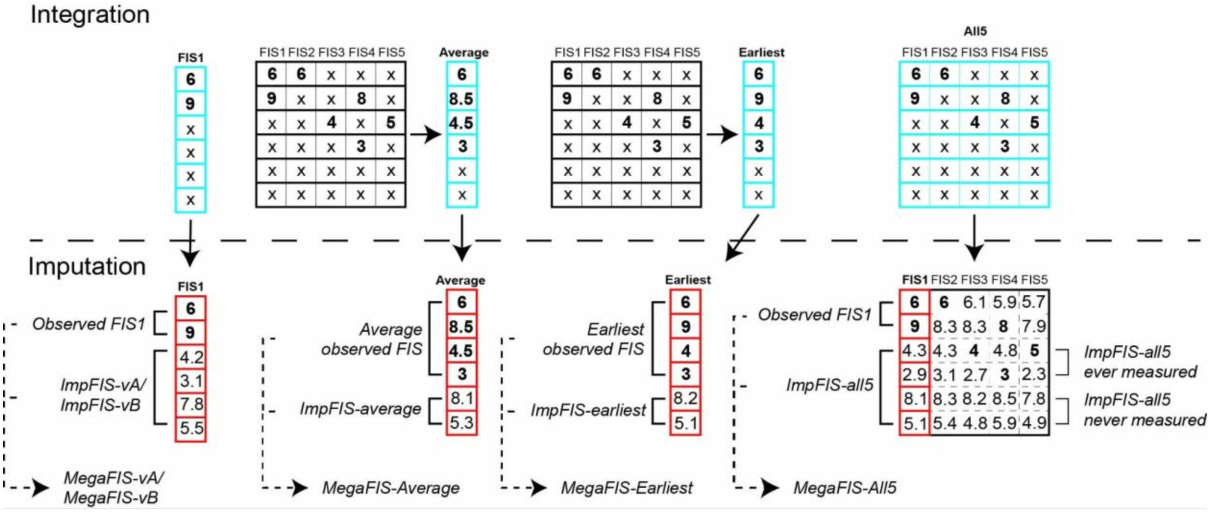

***Schematic illustrating the various strategies for integration and imputation of fluid intelligence.*** (This is an expanded version of Extended Data Figure 1.) The top panel shows how the summary measures are derived from the five different FI tests ("integration") and what is included in the imputation (blue grid) along with the selected imputation variables. Although this figure shows raw FIS scores for clarity, the actual FIS scores were transformed prior to imputation (See Methods). In the average FIS approach, scores for an individual (row in grid) are combined into one average per individual, then individuals without any FI scores are imputed. In the earliest FIS approach, the first score in time is selected, and individuals without any scores are imputed. The bottom panel illustrates how missing values are filled in through imputation following various strategies, and the observed and imputed values are then combined via mega-analyses, as described in Supplementary Notes 2 and 3 respectively.

#### Supplementary Figure 6

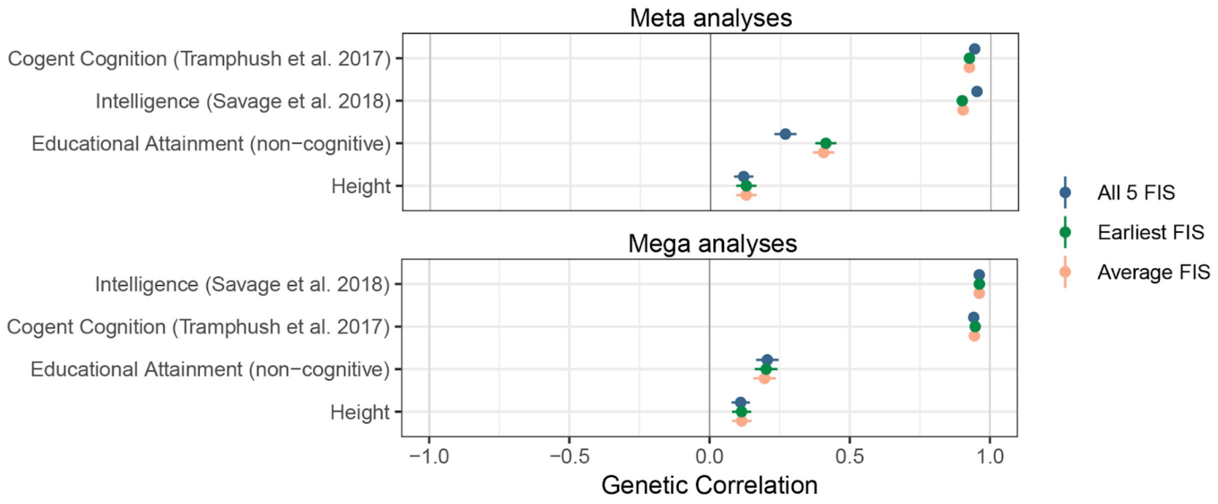

***Genetic correlation for combined GWASs.*** Top panel: imputed and measured values were combined by meta-analysis for each of the three imputation approaches. Bottom panel: Imputed and measured values were scaled separately and combined in mega-analyses for each of the three imputation approaches.

#### Supplementary Figure 7

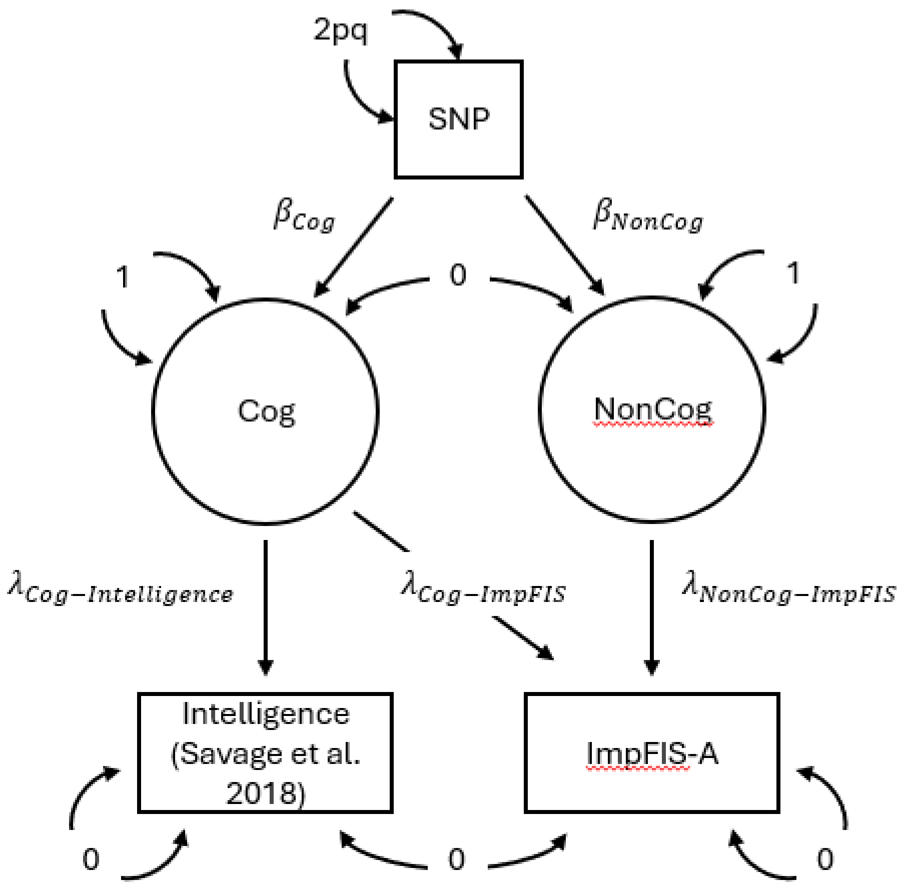

***GWAS-by-subtraction model used to derive non-cognitive imputed FIS (NonCogIFIS).*** Boxes indicate observed variables from GWAS, circles represent latent factors for cognitive (Cog) and non-cognitive (NonCog) signals. Bidirectional arrows represent variances for the respective variables. The variance of SNP is defined as 2pq, representing genetic variance under Hardy-Weinberg equilibrium. Latent factor variances are fixed to 1 and observed variable variances and covariances are fixed to 0. This is to ensure that all variance of the model is explained by the defined latent factors. β parameters indicate effect sizes of a SNP on respective latent factors, $\lambda$ parameters indicate path loadings obtained from regressing observed variables on the latent variables. Genetic signals that are shared between GWASs of Intelligence and ImpFIS-A are assigned to latent factor Cog, the residual signal in ImpFIS-A is assigned to NonCog.

### Supplementary Figure 8

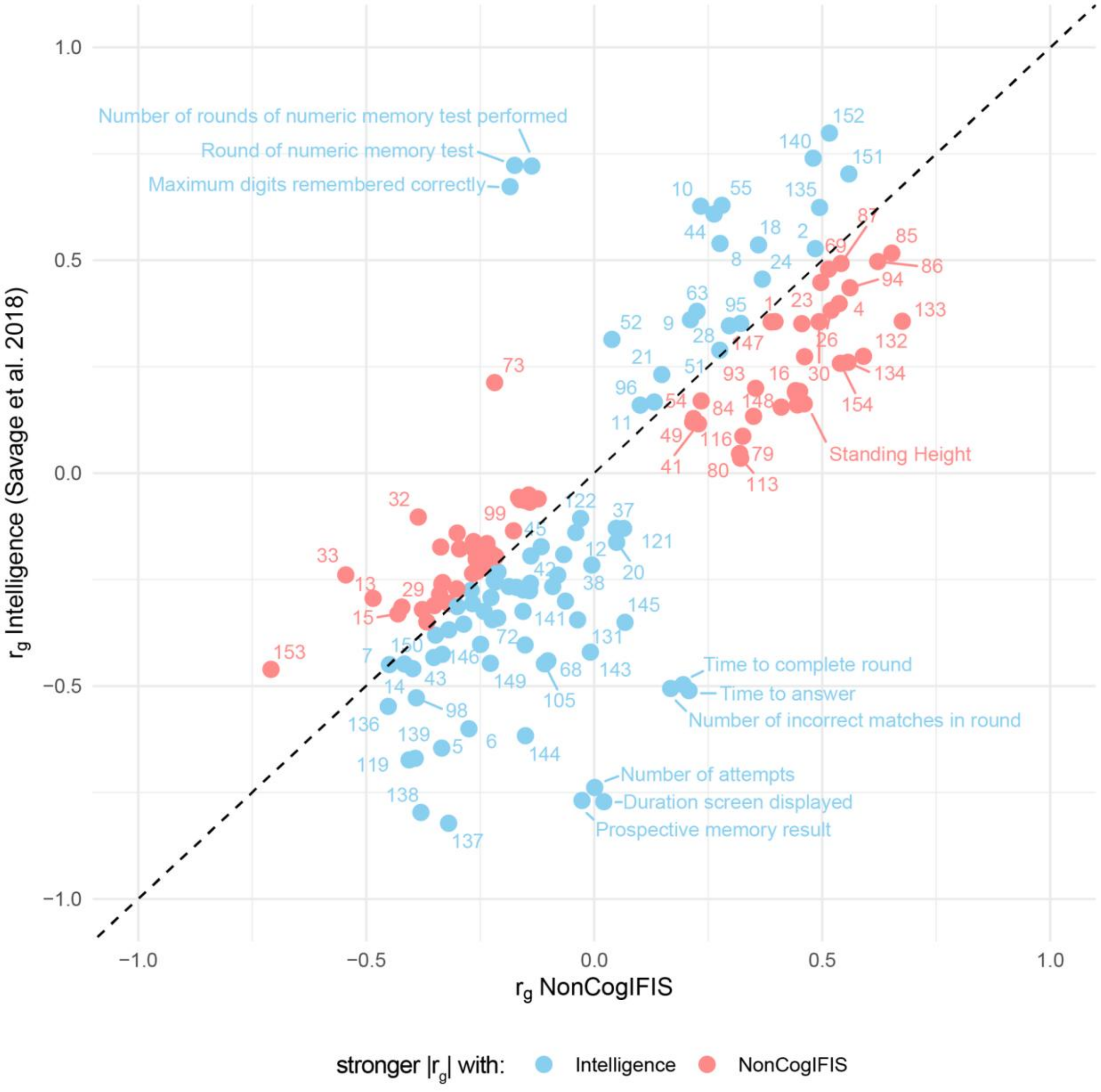

***Correlation based imputation phenotype selection.*** This table shows the genetic correlation of each trait used in imputation with measured intelligence (Savage et al.) and NonCogIFIS. NonCogIFIS contains the genetic signal of the ImpFIS-A GWAS that is not captured by the measured intelligence GWAS by Savage et al. *red dots |rg(noncogFIS)| > |rg(Intelligence)|; blue dots |rg(Intelligence)| > |rg(noncogFIS)|. Labels correspond with the labels in Supplementary Table 2.*

#### Supplementary Figure 9

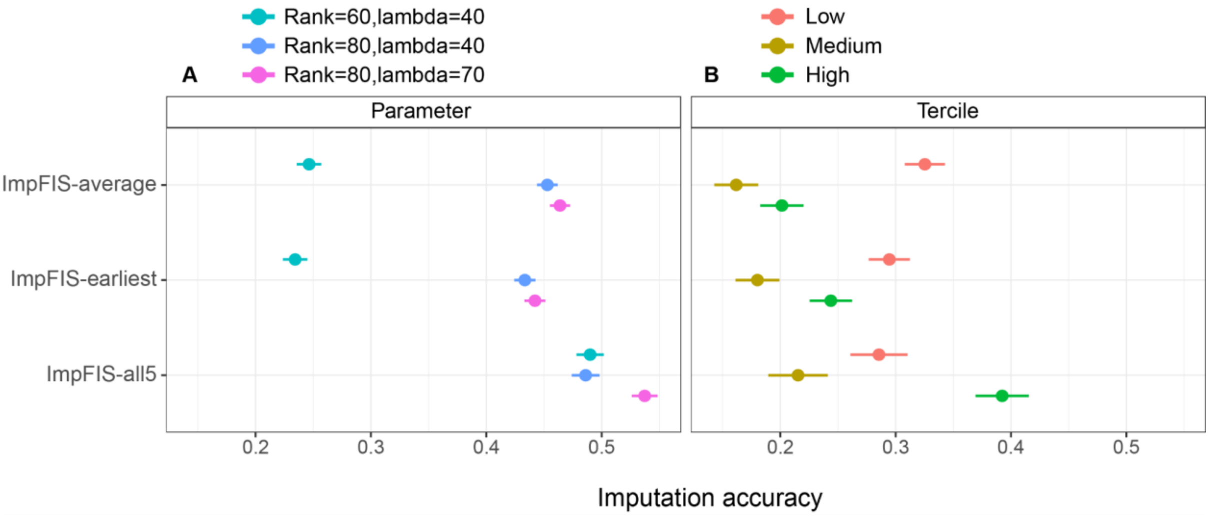

***Imputation accuracies across different FIS scores.*** A) Shows the overall accuracy when using different imputation parameters. B) Shows the accuracy obtained with rank=80 and lambda=70 for three tranches of people divided by their observed FIS value (FIS1, observed average FIS, observed earliest FIS).

### Supplementary Figure 10

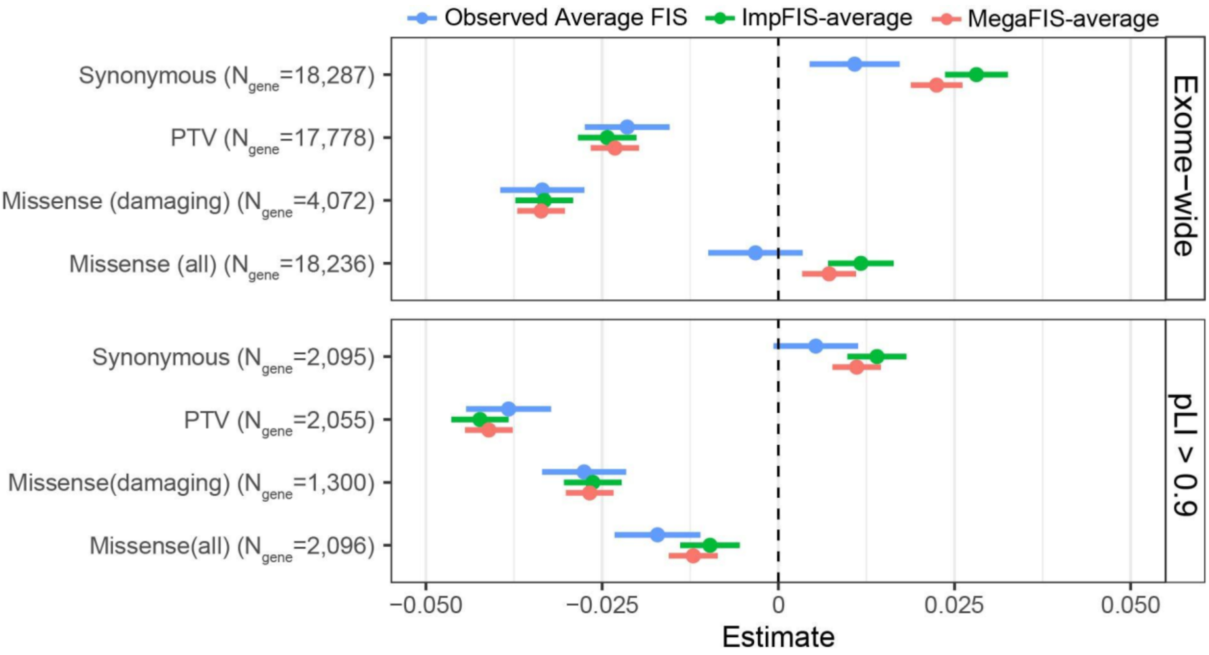

***Impact of exome-wide burden of rare protein-coding variants for FIS in EUR samples in UK Biobank.*** The effects of protein-truncating, all missense, damaging missense and synonymous variant burden on observed average FIS (blue), ImpFIS-average (green), and MegaFIS-average (red) across the exome (top) and in genes with high LOF intolerance (pLI ≥ 0.9) (bottom). Note that these three subsets are referred to as “observed FIS”, “imputed FIS” and “combined FIS” respectively in the main text. Unrelated UKB EUR samples were included in this analysis (n = 328,795). pLI is the probability of being LOF-intolerant as recorded in the gnomAD database. The number of genes included in each burden was labelled. Data are presented as effect size estimates (β) with 95% confidence intervals (CIs).

### Supplementary Figure 11

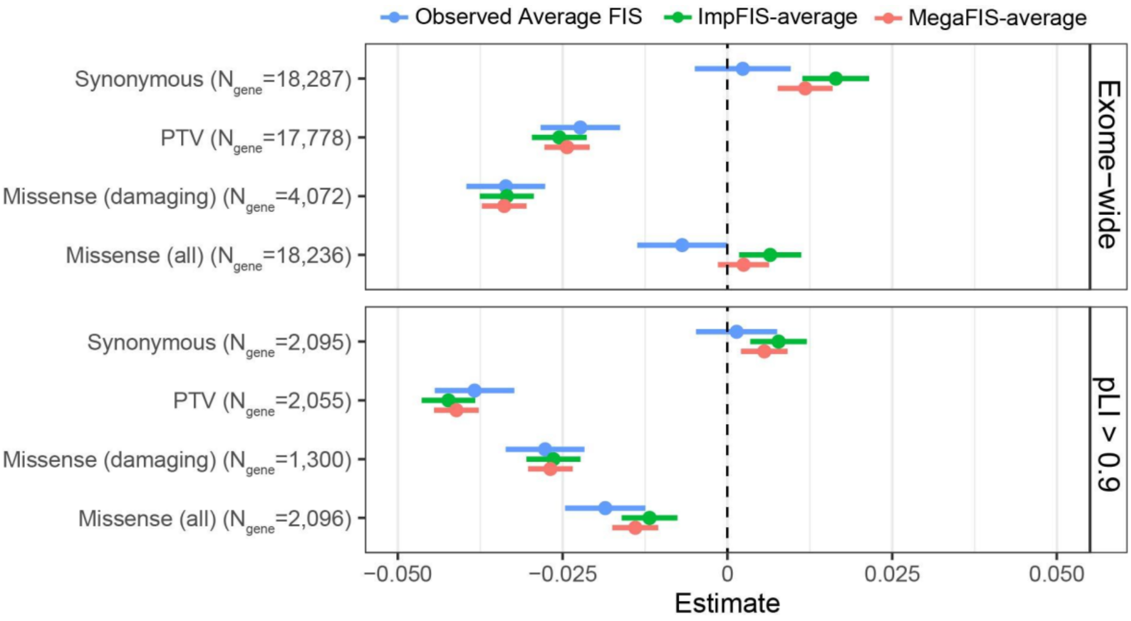

***Exome-wide burden of rare protein-coding variants for FIS in EUR samples in the UKB controlling for the number of singleton synonymous variants per individual.*** The effects of protein-truncating, all missense, damaging missense and synonymous variant burden on observed average FIS (blue), ImpFIS-average (green), and MegaFIS-average (red) across the exome (top) and in genes with high LOF intolerance (pLI ≥ 0.9) (bottom). Note that these three subsets are referred to as "observed FIS", "imputed FIS" and "combined FIS" respectively in the main text. Unrelated UKB EUR samples were included in this analysis (n = 328795). pLI is the probability of being LOF-intolerant as recorded in the gnomAD database. The number of genes included in each burden was labeled. Data are presented as effect size estimates (β) with 95% confidence intervals (CIs). Observed average FIS, ImpFIS-average, and MegaFIS-average are marked with blue, green and red.

#### Supplementary Figure 12

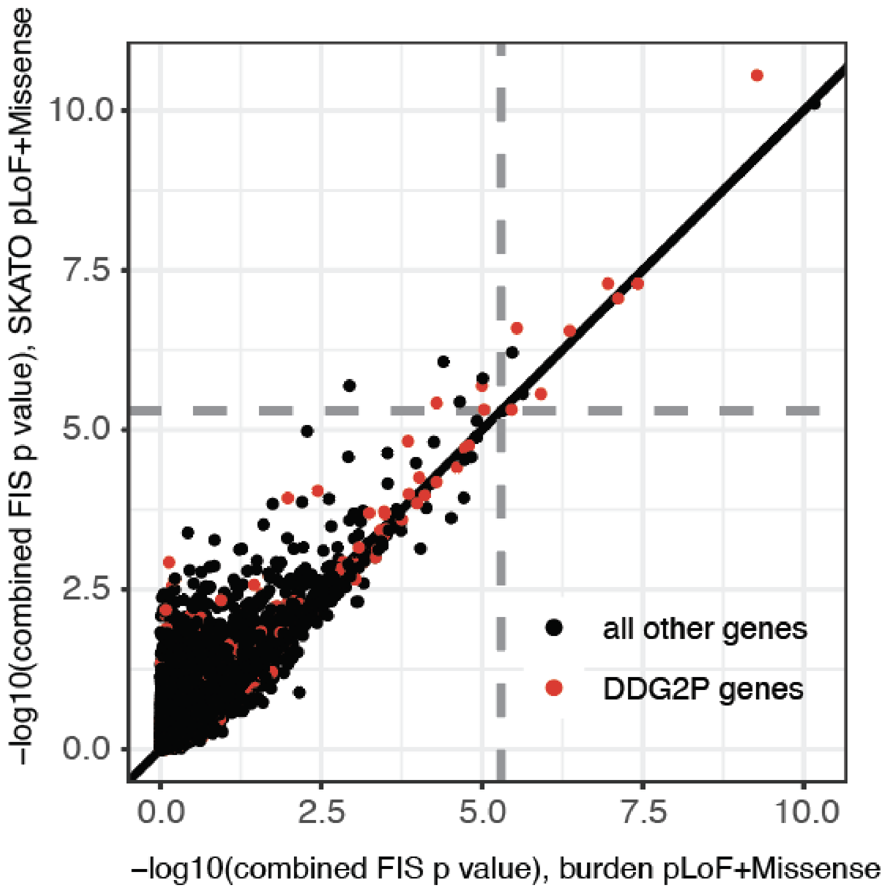

***Results from gene burden test versus SKAT-O***. -log10(p-values) obtained with SKAT-O versus burden tests using PTV+damaging missense variants, for combined FIS. Gray dashed lines indicate the empirical p value cutoff for an FDR <1% in our gene association tests ($p<5.03 \times 10^{-6}$).

#### Supplementary Figure 13

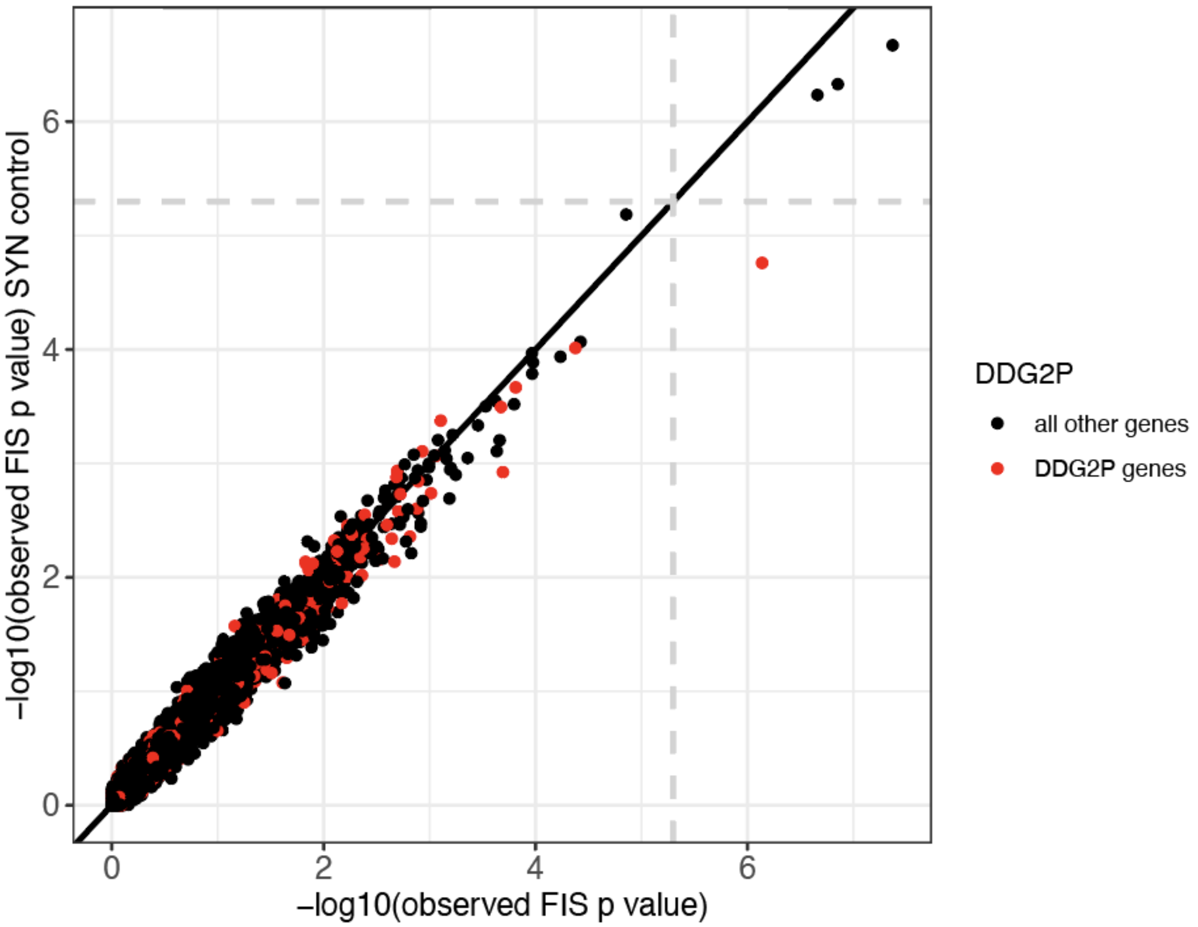

***Results from gene-based rare variant burden tests before and after conditioning on the number of singleton synonymous variants.*** On the x-axis is the -log10(p) for the PTV burden test and on the y-axis is the -log10(p) after conditioning on the number of singleton synonymous variants an individual carries using observed FIS. Gray dashed lines indicate the empirical p value cutoff for an FDR <1% in our gene association tests ($p<5.03x10^{-6}$).

### Supplementary Methods

#### Introduction to the replication cohorts

The Avon Longitudinal Study of Parents and Children (ALSPAC) [37,38], and the Millennium Cohort Study (MCS) are two British birth cohorts that followed a group of participants from their birth. The studies collected information on education and employment, family and parenting, physical and mental health, and social attitudes, as well as applying cognitive tests at various ages. ALSPAC recruited 14,833 women based in Avon, UK who were pregnant during 1991 and 1992 and eventually included a total of 14,901 children of predominantly white ethnicity (>95%). In addition, 12,113 partners of pregnant women also participated in the study and 3,807 are still enrolled. Please note that the study website contains details of all the data that is available through a fully searchable data dictionary and variable search tool.[39] Ethical approval for the study was obtained from the ALSPAC Ethics and Law Committee and the Local Research Ethics Committees. MCS is a UK-wide nationally representative cohort that recruited 18,818 children born between 2000 and 2002. It intentionally over-sampled areas with high child poverty, large ethnic minority populations, and smaller UK nations (Wales, Scotland and Northern Ireland). INTERVAL is a prospective cohort study of about 50,000 participants nested within a pragmatic randomized trial of blood donors [40]. Between 2012 and 2014, blood donors aged 18 years and older were recruited from 25 National Health Service Blood and Transplant (NHSBT) donor centers across England. All participants gave informed consent before joining the study and the National Research Ethics Service approved this study (11/EE/0538). Participants were predominantly healthy individuals since people with major disease (myocardial infarction, stroke, cancer etc.) are excluded from blood donation, as well as those with recent illness or infection. Participants completed online questionnaires about basic lifestyle and health-related information, including self-reported height and weight, ethnicity, current smoking status, alcohol consumption, doctor-diagnosed anemia, use of medications (hormone replacement therapy, iron supplements) and menopausal status.

#### Quality control and imputation of ALSPAC and MCS genotype data

Quality control procedures included excluding individuals with high missing genotype rates (>3%) and filtering autosomal SNPs to retain those with MAF >0.5%, missingness <3%, and Hardy–Weinberg equilibrium $p > 1\times10^{-5}$. Relatedness was inferred using KING [76], and a set of unrelated individuals was derived by iteratively removing participants with excessive inferred kinship. European ancestry was confirmed by projecting samples onto 1,000 Genomes Phase 3 data using the smartpca function from EIGENSOFT, with variants pruned for linkage disequilibrium (pairwise $r^2 < 0.2$ in sliding windows) and exclusion of high/long-range LD regions (including the HLA). Principal component analysis was then conducted on the unrelated samples, and all individuals were projected into this space. Prior to imputation, SNPs that were palindromic, absent from the reference panel, or exhibited allele mismatches were removed. Genotypes were subsequently imputed to the TOPMed r2 reference panel via the TOPMed imputation server[41], and only well-imputed common variants (Minimac4 $R^2 > 0.8$; MAF >1%) were retained for analysis.

#### Quality control of ALSPAC and MCS whole-exome sequencing data

For both ALSPAC and MCS cohorts, the quality control of Whole exome sequencing data was described in the previous paper. First, we used GATK v4.2 to call short variants (SNVs and indels) in 11,994 samples from ALSPAC and 11,916 samples from MCS. We then applied sample quality control (QC) measures to remove outliers on several metrics (e.g., heterozygosity, variant counts) and likely sample mismatches. To identify low-quality variants (variant QC), we trained a random forest on pre-defined truth sets in each cohort separately and then used the random forest filtering in combination with genotype-level and missingness filters to balance precision, recall, true and false positive rates, and synonymous transmission ratios. Specifically, we filtered SNVs (genotypes set to missing) if they an allele depth (DP) < 5, a heterozygous allele balance ratio (AB) < 0.2, or a genotype quality (GQ) < 20 (ALSPAC) or < 15 (MCS) and Indels using these thresholds: DP < 10, AB < 0.3, GQ <10 (ALSPAC) or GQ < 20 (MCS). We finally excluded the variants that failed the random forest filtering or if the fraction of missing genotypes (missingness) exceeded 0.5. In summary, we retained 8,436 children and 3,215 parents in ALSPAC and 7,667 children and 6,925 parents in MCS.

#### Quality control and imputation of INTERVAL genotype data

The genotyping and imputation protocol and QC for the INTERVAL samples (n ≈ 50,000) have been described previously in detail [42]. INTERVAL samples were genotyped in ten batches using DNA extracted from buffy coat on the Affymetrix Axiom UK Biobank array, which assayed approximately 830,000 variants. High-quality autosomal variants (MAF >0.05, HWE $p > 1\times10^{-6}$, and low inter-variant LD with $r^2 \leq 0.2$) were employed to identify duplicate samples (using a PLINK Method-of-Moments IBD threshold of 0.9) and to exclude non-European individuals based on principal component analysis (PCA), where subjects with PC1 or PC2 scores <0 relative to 1000 Genomes major ancestry populations were removed.

Subsequent variant-level filtering within each batch excluded variants that deviated strongly from Hardy–Weinberg equilibrium ($p < 5\times10^{-6}$) and those with a call rate <97%. Variants failing in multiple batches were dropped from the merged dataset. Sample contamination was assessed using the relationship between allele frequency and probe intensity, and samples with >10% contamination, or 3–10% contamination coupled with high relatedness, were removed. An additional set of ~100,000 high-quality variants was used for global IBD analysis (to remove across-batch duplicates and overlapping participants with UK Biobank/UK BiLEVE) and for PCA using flashpca. Outlier individuals were excluded based on lower principal components, and the final PCA was rerun to generate covariates for downstream analyses, resulting in 43,059 participants.

Prior to imputation, further QC steps established a high-quality imputation scaffold by retaining autosomal, bi-allelic, non-monomorphic variants with a global HWE filter ($p > 5\times10^{-6}$), a within-batch call rate >99%, and a global call rate >75%. Non-autosomal, multi-allelic, and inconsistent variants were removed. Phasing was performed with SHAPEIT3 using chunks of 5,000 variants (with a 250-variant overlap), and imputation was conducted on the Sanger Imputation Server using the PBWT algorithm with a combined 1000 Genomes Phase 3–UK10K reference panel. This process yielded approximately 87.7 million imputed variants.

Imputation accuracy was subsequently validated against whole-exome sequencing data from 3,976 participants, confirming high levels of non-reference concordance and precision across common, low-frequency, and rare variants.

#### Calculating PRS in INTERVAL

After removing 1,557 individuals who also participated in UKB and after removing one of each related pair (up to 3$^{rd}$ degree), 40,813 individuals remained. PRS was calculated from the imputed genotype data in PLINK v1.9 using 967,141 variants, after filtering for imputation quality (INFO) score <0.7, minor allele count <8 and HWE p-value $< 5 \times 10^{-6}$, similar to Sun et al.[43].

### References

1. Trampush, J. W. *et al.* GWAS meta-analysis reveals novel loci and genetic correlates for general cognitive function: a report from the COGENT consortium. *Mol. Psychiatry* **22**, 336–345 (2017).
2. Savage, J. E. *et al.* Genome-wide association meta-analysis in 269,867 individuals identifies new genetic and functional links to intelligence. *Nat. Genet.* **50**, 912–919 (2018).
3. Fawns-Ritchie, C. & Deary, I. J. Reliability and validity of the UK Biobank cognitive tests. *PLoS One* **15**, e0231627 (2020).
4. Brown, H. S. & May, A. E. A test–retest reliability study of the Wechsler Adult Intelligence Scale. *J. Consult. Clin. Psychol.* **47**, 601–602 (1979).
5. Demange, P. A. *et al.* Investigating the genetic architecture of noncognitive skills using GWAS-by-subtraction. *Nat. Genet.* **53**, 35–44 (2021).
6. Willer, C. J., Li, Y. & Abecasis, G. R. METAL: fast and efficient meta-analysis of genomewide association scans. *Bioinformatics* **26**, 2190–2191 (2010).
7. Chen, C.-Y. *et al.* The impact of rare protein coding genetic variation on adult cognitive function. *Nat. Genet.* **55**, 927–938 (2023).
8. Furia, F. *et al.* The phenotypic and genotypic spectrum of individuals with mono- or biallelic ANK3 variants. *Clin. Genet.* **106**, 574–584 (2024).
9. Prasad, A. *et al.* A discovery resource of rare copy number variations in individuals with autism spectrum disorder. *G3 (Bethesda)* **2**, 1665–1685 (2012).
10. St Pourcain, B. *et al.* Common variation near ROBO2 is associated with expressive vocabulary in infancy. *Nat. Commun.* **5**, 4831 (2014).
11. Marchman, V. A. & Fernald, A. Speed of word recognition and vocabulary knowledge in infancy predict cognitive and language outcomes in later childhood. *Dev. Sci.* **11**, F9–16 (2008).
12. Reijnders, M. R. F. *et al.* De Novo and inherited loss-of-function variants in TLK2: Clinical and genotype-phenotype evaluation of a distinct neurodevelopmental disorder. *Am. J. Hum. Genet.* **102**, 1195–1203 (2018).
13. Villamor-Payà, M. *et al.* Identification of a de novo mutation in TLK1 associated with a neurodevelopmental disorder and immunodeficiency. *medRxiv* (2023) doi:10.1101/2023.08.22.23294267.

14. Vissers, L. E. L. M. *et al.* Mutations in a new member of the chromodomain gene family cause CHARGE syndrome. *Nat. Genet.* **36**, 955–957 (2004).
15. Sugathan, A. *et al.* CHD8 regulates neurodevelopmental pathways associated with autism spectrum disorder in neural progenitors. *Proc. Natl. Acad. Sci. U. S. A.* **111**, E4468–77 (2014).
16. Li, M. *et al.* Integrative functional genomic analysis of human brain development and neuropsychiatric risks. *Science* **362**, eaat7615 (2018).
17. Iossifov, I. *et al.* The contribution of de novo coding mutations to autism spectrum disorder. *Nature* **515**, 216–221 (2014).
18. Satterstrom, F. K. *et al.* Large-Scale Exome Sequencing Study Implicates Both Developmental and Functional Changes in the Neurobiology of Autism. *Cell* **180**, 568–584.e23 (2020).
19. Lim, E. T. *et al.* Rates, distribution and implications of postzygotic mosaic mutations in autism spectrum disorder. *Nat. Neurosci.* **20**, 1217–1224 (2017).
20. Degenhardt, F. *et al.* Duplications in RB1CC1 are associated with schizophrenia; identification in large European sample sets. *Transl. Psychiatry* **3**, e326 (2013).
21. Bochdanovits, Z. *et al.* Joint reanalysis of 29 correlated SNPs supports the role of PCLO/Piccolo as a causal risk factor for major depressive disorder. *Mol. Psychiatry* **14**, 650–652 (2009).
22. Nievergelt, C. M. *et al.* Discovery of 95 PTSD loci provides insight into genetic architecture and neurobiology of trauma and stress-related disorders. *medRxiv* (2023) doi:10.1101/2023.08.31.23294915.
23. International Mouse Phenotyping Consortium. Mouse gene Dpp8.
24. International Mouse Phenotyping Consortium. Mouse gene Ipo9.
25. Lee, J. J. *et al.* Gene discovery and polygenic prediction from a 1.1-million-person GWAS of educational attainment. *Nat. Genet.* **50**, 1112 (2018).
26. Okbay, A. *et al.* Polygenic prediction of educational attainment within and between families from genome-wide association analyses in 3 million individuals. *Nat. Genet.* **54**, 437–449 (2022).
27. Karlsson Linnér, R. *et al.* Genome-wide association analyses of risk tolerance and risky behaviors in over 1 million individuals identify hundreds of loci and shared genetic influences. *Nat. Genet.* **51**, 245–257 (2019).
28. Baselmans, B. *et al.* The genetic and neural substrates of externalizing behavior. *Biol. Psychiatry Glob. Open Sci.* **2**, 389–399 (2022).

29. Open Targets Platform. https://platform.opentargets.org/credible-set/651a7e1654d0a7f9a9dde0f4850c0365.

30. Speidel, D. *et al.* A family of Ca2+-dependent activator proteins for secretion. *J. Biol. Chem.* **278**, 52802–52809 (2003).

31. Lee, J. J. *et al.* Gene discovery and polygenic prediction from a genome-wide association study of educational attainment in 1.1 million individuals. *Nat. Genet.* **50**, 1112–1121 (2018).

32. Sadakata, T. *et al.* Autistic-like phenotypes in Cadps2-knockout mice and aberrant CADPS2 splicing in autistic patients. *J. Clin. Invest.* **117**, 931–943 (2007).

33. Bulik-Sullivan, B. K. *et al.* LD Score regression distinguishes confounding from polygenicity in genome-wide association studies. *Nat. Genet.* **47**, 291–295 (2015).

34. Bulik-Sullivan, B. *et al.* An atlas of genetic correlations across human diseases and traits. *Nat. Genet.* **47**, 1236–1241 (2015).

35. Watanabe, K., Taskesen, E., van Bochoven, A. & Posthuma, D. Functional mapping and annotation of genetic associations with FUMA. *Nat. Commun.* **8**, 1826 (2017).

36. Young, A. I. *et al.* Mendelian imputation of parental genotypes improves estimates of direct genetic effects. *Nat. Genet.* **54**, 897–905 (2022).

37. Boyd, A. *et al. The 'Children of the 90s'; the Index Offspring of The Avon Longitudinal Study of Parents and Children*. (ALSPAC).

38. Fraser, A. *et al.* Cohort Profile: the Avon Longitudinal Study of Parents and Children: ALSPAC mothers cohort. *Int. J. Epidemiol.* **42**, 97–110 (2013).

39. Explore data and samples. https://www.bristol.ac.uk/alspac/researchers/our-data/.

40. Moore, C. *et al.* The INTERVAL trial to determine whether intervals between blood donations can be safely and acceptably decreased to optimise blood supply: study protocol for a randomised controlled trial. *Trials* **15**, 363 (2014).

41. McCarthy, S. *et al.* A reference panel of 64,976 haplotypes for genotype imputation. *Nat. Genet.* **48**, 1279–1283 (2016).

42. Astle, W. J. *et al.* The Allelic landscape of human blood cell trait variation and links to common complex disease. *Cell* **167**, 1415–1429.e19 (2016).

43. Sun, B. B. *et al.* Genomic atlas of the human plasma proteome. *Nature* **558**, 73–79 (2018).
